# Comparison of methods for assessing *Chlamydia trachomatis* transmission intensity: a systematic review

**DOI:** 10.1101/2025.07.16.25331656

**Authors:** Kristen K Renneker, Che-Chi Lin, Jennifer L Hsieh, PJ Hooper, Robert Butcher, T Déirdre Hollingsworth, Diana L Martin, Anthony W Solomon, Emma M Harding-Esch

## Abstract

**Topic:** To assess the relationship between field-graded trachomatous inflammation—follicular (TF) and other methodologies for evaluating *Chlamydia trachomatis* (*Ct)* transmission intensity.

**Clinical relevance:** TF has limitations as the sole indicator of *Ct* transmission for trachoma programmatic decision-making. The relationships between TF and other indicators, including other clinical signs, photography, infection, and serology have yet to be synthesized.

**Methods:** A systematic review was conducted. Five databases (MEDLINE, EMBASE, Global Health database, Scopus, and Global Index Medicus) were searched on October 19, 2022, and the e-Theses Online Service was searched on April 12, 2023. Studies from 1987 onward that reported primary data collection of field-graded TF in children and at least one other indicator were included. To compare field-graded vs. photo-graded TF, the mean difference in prevalence and 95% confidence intervals (CIs) were calculated. To compare field-graded TF against the other indicators, weighted correlation coefficients and p-values were calculated in pre-vs. post-antibiotic mass drug administration (MDA) settings. The review protocol was prospectively registered with PROSPERO (CRD42022356013).

**Results:** A total of 35,764 studies were screened, yielding 235 included studies from 49 countries, spanning the years (of data collection) 1991–2021. For field-graded vs. photo-graded TF (n=10), the mean difference in prevalence was 0.7 percentage points, 95% CI (−15.2–16.7). The relationship between TF and other indicators was stronger pre-vs. post-MDA: TF vs infection (R^2^: 0.43, p=0.003 vs. R^2^: 0.002, p=0.788); TF vs seroprevalence (R^2^: 0.56, p<0.001 vs. R^2^: 0.03, p=0.353); and TF vs seroconversion rate (SCR) (R^2^: 0.52, p=0.012 vs. R^2^: 0.26, p=0.061). Post-MDA, infection and SCR were highly correlated (R^2^: 0.71, p=0.001). In studies reporting data used for programmatic decision-making, 48% of the areas warranting MDA based solely on TF had at least one other indicator categorized as indicating “low” or “medium” *Ct* transmission intensity.

**Conclusions:** The correlation between TF and measures of infection and serology weakens post-initiation of MDA, which is strongly suggestive of poor performance of TF prevalence for guiding programmatic decision-making post-MDA. Measures of infection and SCR are strongly correlated with each other post-MDA. Infection and/or serology should be considered to help guide programmatic decision-making post-MDA.

**Target Journal:** Ophthalmology

## Introduction

Trachoma is a neglected tropical disease which can cause blindness. In the infectious phase of the disease, repeated conjunctival infection with ocular strains of the bacterium *Chlamydia trachomatis* (*Ct*) results in inflammation of the conjunctiva, known as active trachoma.^1^ Trachoma has a number of clinical features, including signs such as Herbert’s pits and pannus at the corneal limbus, which were captured in the 22-sign 1966 World Health Organization (WHO) grading scheme.^2^ However, the complexity of that system precluded its reliable use at scale. As such, WHO condensed this into the Follicles, Papillae, Cicatrices (FPC) system in 1981^3^ and then further into the simplified trachoma grading scheme in 1987. The simplified system was designed to be simple enough to be reliably graded by non-specialists as part of community-level field surveys.^4^ Since then, the WHO simplified system has received wide acceptance and is now the system used for programmatic decision-making. It defines active trachoma as the presence in either eye of trachomatous inflammation—follicular (TF) and/or trachomatous inflammation—intense (TI)^4^ (Table 1).

**Table 1:** Summary of indicators of ocular *Chlamydia trachomatis* (*Ct*) transmission included in this review

| Indicator | Description | Collection method | Strengths and weaknesses as a programmatic marker |
| --- | --- | --- | --- |
| Trachomatous inflammation—follicular (TF) | The presence of five or more follicles of at least 0.5 mm in diameter in the central upper tarsal conjunctiva<sup>4</sup> | Clinical exam, measured as part of standard trachoma prevalence surveys<sup>5, 6</sup> | Strengths: Easy to measure reproducibly;<sup>4</sup> high correlation with Ct infection pre-MDA;<sup>7, 8</sup> currently recommended for programmatic decision-making by World Health Organization (WHO). Weaknesses: Poor correlation with Ct infection after MDA;<sup>7</sup> feasibility of standardizing measurement<sup>5, 6</sup> in low-prevalence settings. |
| Trachomatous inflammation—intense (TI) | Pronounced inflammatory thickening of the upper tarsal conjunctiva that obscures more | Clinical exam, measured as part of standard trachoma | Strength: Evidence that TI is highly correlated with infection,<sup>9</sup> even in areas with a history of MDA (at implementation-unit levels).<sup>10</sup> Weaknesses: Low specificity for ocular Ct infection;<sup>11-13</sup> lack of evidence of a post-MDA correlation between TI and infection at the community level;<sup>7</sup> harder to |
|  | than half of the deep tarsal vessels<sup>4</sup> | prevalence surveys<sup>5, 6</sup> | standardize grading of TI at-scale compared to TF; population-level prevalence of TI not currently recommended by WHO for programmatic decision-making. |
| Pannus | Invasion of blood vessels and fibrous tissue into the cornea, measured as the distance of such ingrowth from the limbus<sup>2</sup> | Clinical exam | Strength: Some evidence that presence of upper pole pannus correlated with later severe conjunctival scarring with eyelid deformity.<sup>14</sup> Weaknesses: Lack of evidence for interpretation; a challenge to measure reproducibly without a slit lamp.<sup>2</sup> |
| Herbert's pits | Areas of the limbus where previous inflammation has resolved with scarring<sup>2</sup> | Clinical exam | Strength: Evidence that Herbert's pits may be associated with having had inflammatory trachoma.<sup>14, 15</sup> Weaknesses: Lack of evidence for interpretation; a challenge to measure reproducibly without a slit lamp.<sup>2</sup> |
| Conjunctival photography | Photograph of the tarsal conjunctiva | Photographs of the conjunctiva are taken and then later graded for signs of trachoma | Strengths: May mitigate the challenge of training field graders, provide an opportunity for validation of field grades, and reduce biases compared to field-grading.<sup>16</sup> Weaknesses: Lack of standards in the literature on camera type and quality control methods;<sup>16</sup> high incidence of ungradable images; implementation considerations such as ethics, case management, and data systems.<sup>17</sup> |
| *Ct* infection | Identification of the presence of *Ct*, most commonly through a nucleic acid amplification test | Swab of the upper tarsal conjunctiva | Strength: Direct measurement of target of antibiotics. Weakness: Concerns around potential for increased cost and infrastructure to measure (compared to clinical signs).<sup>18</sup> |
| Serology | Identification of the presence of antibodies that indicate prior exposure to *Ct* | Blood collection via finger-prick | Strengths: Direct measurement of exposure; capacity for integration with other serosurveillance programs;<sup>19</sup> ability to measure cumulative exposure to *Ct* infection over time.<sup>20</sup> Weaknesses: Antigens in conjunctival *Ct* strains are shared by urogenital *Ct* strains.<sup>20</sup> |

WHO recommends the SAFE strategy (surgery, antibiotics, facial cleanliness and environmental improvement) for the elimination of trachoma as a public health problem.^21^ The purpose of the A, F and E components of SAFE is to reduce the prevalence and transmission of ocular *Ct*. However, at the time the simplified grading system was developed, measuring ocular *Ct* infection directly (for example, with molecular analysis of conjunctival swabs) at scale was not feasible in most trachoma-endemic settings. As a consequence, the district-level prevalence of TF in children aged 1–9 years (TF_1–9_), as measured by trained field graders, is used to guide the need for and prioritization of implementation of A, F and E. Indeed, one of the three criteria for elimination of trachoma as a public health problem is that TF_1–9_ is <5% in all formerly endemic districts.^22^

As the global prevalence of trachoma has declined, several limitations of field-graded TF have emerged (Table 1). These limitations are likely to be exacerbated as we approach and achieve elimination of trachoma, with TF also being unreliable as a tool for post-elimination surveillance. There are a number of indicators for *Ct* transmission that have been described as “alternative” to (i.e., collected in place of) or “complementary” to (i.e., collected alongside) field-graded TF (Table 1). There is currently no WHO guidance for the routine use of serological or infection testing for population-based prevalence surveys; therefore, data on TF will continue to be collected as routine for the foreseeable future, with the other indicators complementing the TF data and decisions being made on the whole body of evidence.

Therefore, for the purposes of this review, these other indicators will be referred to as complementary. Two complementary indicators of particular interest have emerged as having the potential to help guide programs in decision-making: prevalence of *Ct* infection as measured by PCR and seroconversion rate (SCR); the latter preferable to seroprevalence due to the implicit adjustment for age.^23^

Separate reviews looking at the association between field-graded TF and *Ct* infection^7^, photo-graded TF^16^, and serology^20^ have been published in the past decade; however, these results have yet to be collated or assessed as a single body of evidence. In addition, since the search dates of those previous reviews, considerable amounts of new data have been published. The research question for this review was: “What is the relationship between field-graded TF and other methodologies for evaluating ocular *Ct* transmission intensity?” Our objectives were to identify studies collecting data on trachoma assessed via field clinical grading of TF and at least one complementary indictor, to explore the implications of the use of complementary indicators on programmatic decision-making in populations with measures of TF, infection prevalence, and SCR, and to identify gaps in understanding of complementary indicators, including their use for decision-making, especially in settings post-initiation of MDA. A full PICOST (population, intervention, comparators, outcomes, situation, and study type) framework was developed^24^ (Supplemental File S1).

## Methods

### Protocol

This review protocol was prospectively registered on PROSPERO (CRD42022356013) in March 2022. Prior to commencing the search, the search strategy was reviewed using PRESS guidelines^25^ by a reference librarian at the London School of Hygiene & Tropical Medicine (LSHTM). As all data in the review were already in the public domain, no formal ethical review was required.

### Eligibility criteria

We included studies that met the following inclusion criteria: a) on trachoma; b) publication from 1987 (the publication year of the WHO simplified grading system^4^ for trachoma programmatic decision-making); c) publication in English, French, Spanish, or Portuguese; d) reporting measurement of TF or F2/F3^a^ from the FPC system by clinical field-grading and at least one other indicator; e) reporting primary data collection results; and f) on humans.

### Search methods

We conducted the systematic search in six databases: Medline (US Medical Library/Ovid), Embase (Elsevier/Ovid), Global Health (CABI/Ovid), Global Index Medicus (WHO), Scopus (Elsevier), and e-Theses Online Service (EThoS, University of London). The search terms were split into two main components joined with an AND term:

- Component 1: trachoma
- Component 2: complementary indicator terms joined with an OR term:
  - Non-TF (or non-F) clinical field-graded measures of current or previous infection (e.g., TI, pannus, and Herbert’s pits)
  - Infection
  - Serology
  - Conjunctival photography

EThoS did not allow Boolean searches, therefore the term “trachoma” was searched and the result set checked manually for inclusion. The full search term lists for each database are included as Supplemental File S2. All databases except for EThoS were searched on October 19, 2022. EThoS was searched on April 12, 2023.

### Study selection

Two-stage screening was conducted in Covidence.^26^ Two authors independently reviewed titles and abstracts of search results against the inclusion criteria for potentially eligible publications. Once a consensus was reached on which titles/abstracts to include, the same two authors independently reviewed the full text of references that passed the title and abstract screen. Where the reviewers agreed, the search result was included; where they did not, search results were discussed and a consensus was reached.

Where full texts were excluded, the reason for exclusion was recorded. Searches for full texts were completed through the online libraries of Emory University and LSHTM, using the available search tools provided by relevant journal websites, and by contacting corresponding authors where contact information could be found. The following exclusion criteria were used: a) studies with missing abstract, full text, or other key information that could not be found; b) studies reporting only on genomic data, cellular data, or non-clinical science; c) opinion pieces; d) studies only reporting on conditions not related to ocular *Ct* (including sexually transmitted infections, genital chlamydia, or other infections); e) studies only reporting trachoma measures that are not active trachoma (e.g., trachomatous trichiasis [TT]); f) case studies; g) systematic reviews, modelling, economic evaluations, or other secondary analyses; h) studies duplicating results in a less-comprehensive way (e.g., conference proceedings that were later published as full peer-reviewed articles).

### Data extraction

One author extracted the primary data from each included study. The extracted variables included information on year of publication, year of data collection, country of data collection, MDA setting (pre-initiation of MDA or post-initiation of MDA), and type of study (cross-sectional, case-control, etc.). In addition, for each indicator (field-graded TF and at least one of: another clinical indicator, photography, infection, or serology), population tested size, population tested age range, population tested gender, outcome type, and outcome (prevalence by indicator, including 95% confidence intervals (CI) where reported) were extracted. Any reported measures of association between indicators were also extracted. No assumptions about missing data were made; if information was not reported, this was recorded as “unknown” and excluded from relevant analyses. A random 20% sample of included studies was independently extracted by a second reviewer, and the results of these double-extracted studies were compared to check for accuracy and completeness of the primary extraction. When information about history of MDA was not included in a publication, this information was abstracted from the Trachoma Atlas,^27^ which includes global binary (treated/not treated) treatment history information by year. Regions were defined according to WHO classifications.^28^

### Risk of bias assessment

Individual studies were assessed for quality and risk of bias using the Joanna Briggs Institute (JBI) critical appraisal Checklist for Prevalence Studies^29^ or Checklist for Diagnostic Test Accuracy Studies^30^ based on study type. Prevalence studies that included measures of association between indicators were assessed using both checklists. A per-study score was calculated by considering a response of “Yes” to each checklist item as a marker of high methodological quality, and these scores were averaged to produce an overall measure of reported methodological quality; “good” quality was defined as a score of 70% or higher. (Note that this measures quality pertaining to this review, rather than general study quality.) The body of evidence for each outcome was assessed according to GRADE guidelines.^31^

### Data synthesis and analysis

During analysis, comparisons not measuring TF_1–9_ were excluded, as comparing the performance of other indicators to this commonly measured indicator of programmatic importance was the primary objective. Studies not reporting a point prevalence from each studied indicator were excluded. Studies reporting more than one comparison by each method were de-duplicated in the following manner: if serological results using different antigens were reported, results using the Pgp3 antigen (only) were kept and others (i.e. CT-694 or a combination of CT-694 and Pgp3) discarded; if multiple serological assay types were reported, results using the ELISA assay type were kept and others (e.g. lateral flow assay results) discarded, as in papers reporting multiple assay types, the ELISA assay was considered the reference standard; if SCRs were estimated at multiple time points, those estimated for time periods aligning with measurements of TF_1–9_ were kept, while others were discarded. Measurements of complementary indicators tested only in clinically positive children were excluded. While the outcomes of interest (TF_1–9_, other clinical indicator prevalence, infection prevalence, seroprevalence, and SCR) are each continuous measures, it can be useful to divide such measures into categories, which are either currently used (in the case of TF_1–9_) or could potentially be used in the future to determine programmatic action. In order to explore the relationships of TF_1–9_ with complementary indicators, analyses were performed comparing continuous measures against each other, by comparing categorical TF_1–9_ vs. continuous measures of complementary indicators, and by comparing categorical TF_1–9_ vs. potential categories of complementary indicators. Similar methods were also used in order to compare complementary indicators against each other.

#### Field-graded vs. photo-graded TF_1–9_

The agreement between field-graded and photo-graded TF was assessed through Bland-Altman analysis.^32^ The difference between the two measures from the same population was calculated and the mean taken in order to find the mean difference. CIs for the mean difference were calculated. Mean differences and CIs were calculated based on camera type (digital single-lens reflex (DSLR) or unaided smartphone, defined as the use of a smartphone camera without any additional devices such as the head-mounted ICAPS system^33^ or Corneal CellScope attachment^34^). Data were visualized with a Bland-Altman plot.

#### TF_1–9_ vs. complementary indicators pre- and post-initiation of MDA

The relationships between TF_1–9_ and the prevalence estimates or SCR estimates obtained in the same populations via different indicators in pre- and post-initiation of MDA settings were visualized via scatterplots. Coefficients of determination (R^2^) between indicators were obtained by calculating and then squaring Pearson’s correlation coefficients. Standard errors, t-values, and p-values were calculated. For significance testing, an alpha of 0.05 was used. Correlation coefficients were weighted based on sample size. Comparisons between TI prevalence and infection prevalence were analyzed in the same manner.

#### Binary TF_1–9_ category vs. continuous measures of complementary indicators

Distributions of continuous complementary indicator results within binary categorical TF_1–9_ results (TF_1–9_ <5% vs. TF_1–9_ ≥5%) were visualized using boxplots, including medians and interquartile ranges [IQRs], and histograms. The lower and upper whiskers of the boxplot extend to 1.5 times the lower and upper IQR, respectively; points outside of this range were plotted individually as outliers.^35^

#### Categorical TF_1–9_ category vs. complementary indicator category in surveys used for programmatic decision-making

To compare measurements of TF_1–9_, infection prevalence, and SCR in studies reporting results of population-based prevalence surveys used for programmatic decision-making, outcome measure estimates were categorized into low, medium, and high categories using the following thresholds: for TF_1–9_, low: <5%,^22^ medium: 5–10%, high: ≥10%;^36^ and for *Ct* infection, low: <1%, medium: ≥1–2%, high: ≥2%.^37^ For SCR, the thresholds used depended on the context of study: in areas that either had no history of MDA or had unusual epidemiology (i.e., demonstrated persistent or recrudescent active trachoma or high reported TF_1–9_ but no or low reported TT) the thresholds used were: low: ≤ 1.6; medium: 1.6–3.8; high: ≥3.8 seroconversions per 100 person-years. In areas post-initiation of MDA (including post-validation) but no reported unusual epidemiology, the thresholds used were: low ≤2.2; medium: 2.2–4.5; high: ≥4.5 seroconversions per 100 person-years. SCR thresholds were based on a recent global analysis.^23^ Two indicators were “concordant” if they fell into the same category in the same population, using the categories defined above.

#### Populations with three indicators (TF_1–9_, infection prevalence, and SCR): continuous and categorical comparisons

To explore implications of decision-making for populations with all three metrics (TF_1–9_, infection prevalence, SCR) reported, the two-way relationships between the continuous indicator values (TF_1–9_ vs. infection prevalence, TF_1–9_ vs. SCR, and infection prevalence vs. SCR) were visualized using scatterplots; coefficients of determination between indicators were obtained as above. Indicators were categorized using the thresholds defined above, and categorical measures were compared; two indicators were “concordant” if they fell into the same category in the same population.

Data cleaning and analyses were conducted in R,^38^ using the tidyverse,^39^ cowplot,^40^ and weights^41^ packages. Due to the heterogeneous nature of the included studies and caution against comparing indicators that measure linked biological phenomena but cannot be considered directly comparable, a formal meta-analysis was not conducted. This manuscript was written in accordance with the Preferred Reporting Items for Systematic reviews and Meta-Analysis (PRISMA) 2020 statement guidelines and checklist (Supplemental file S3).^42^

## Results

### Characteristics of included studies

A total of 35,764 studies were imported for screening. Of those, 21,150 records were unique, and of those, 883 were assessed for eligibility at the full-text screening stage. 238 papers met the inclusion criteria and were included in the review.^8, 10, 33, 34, 43–276^ Comparison between the full extraction and a random 20% of papers that were extracted by a second reviewer yielded an initial match rate of 94%. After reviewing the discrepancies, in all cases the original extraction was accurate. Complete results of the screening process are included in the PRISMA diagram (Supplemental File S4). Of the 129 studies with insufficient information in the abstract and no available full text, 36 were conference abstracts that did not meet the inclusion criteria and 93 were journal articles where the full text could not be found, representing 10.5% of the 883 studies assessed at the full-text stage. A majority of these missing full text papers pre-dated online archives of their respective journals, with 54% of studies published in 1997 or earlier (Supplemental File S5). A table of included studies is included as Supplemental File S6. Studies were included from 49 countries, spanning the years of data collection 1991–2021. Of the 238 included papers, 225 were in peer-reviewed journals, 3 were PhD dissertations,^78, 107, 244^ and 10 were conference abstracts for which a subsequent peer-reviewed article could not be found.^70, 73, 87, 138, 143, 158, 161, 164, 202, 206^ Three pairs of the 238 papers reported on different indicators as part of the same data collection activity, yielding a total of 235 unique studies. The proportion of studies by reported region of data collection was 75% in the African Region, 8% in the Western Pacific Region, 6% in each of the Americas Region and Eastern Mediterranean Region, 3% in the South-East Asia Region, and 1% did not report the country of data collection (Supplemental File S7). A total of 183 studies reported a clinical indicator other than TF, 9 reported photo-graded TF, 106 reported *Ct* infection, and 27 reported using serology, of which 13 reported at least one SCR analysis (Supplemental File S6). Three studies measured field-graded F2/F3,^8, 172, 203^ with one of these^8^ reporting results as TF.

#### Overview of included comparisons

For the purposes of this review, a “comparison” is defined as two indicators measured in the same population. Unless otherwise noted, one of these measurements is TF_1–9_. Note that studies sometimes reported either multiple non-TF indicators in the same population and/or multiple populations in the same study, hence there is not a one-to-one relationship between study and comparison. Table 2 includes the number of studies and comparisons of field-graded TF vs. another indicator. A total of 1,775 comparisons were reported, including comparisons between field-graded TF and other clinical indicators (n=1,001); photo-graded TF (n=10); *Ct* infection (n=350); and serology (n=414). No papers reported data on Herbert’s pits and only two papers reported pannus in children.^172, 277^

**Table 2:** Number of studies and comparisons between field-graded clinical signs of trachomatous inflammation—follicular (TF) and other indicators

| Complementary indicator type | Number of studies | Number of comparisons with TF |
| --- | --- | --- |
| Clinical indicators |  | 1,001 |
| follicular conjunctivitis |  |  |
| <5 follicles | 3 | 13 |
| papillary conjunctivitis |  |  |
| TI (WHO simplified grading system) | 164 | 700 |
| P2/P3 (FPC) | 1 | 4 |
| P3 (FPC) | 4 | 6 |
| follicular and/or papillary conjunctivitis |  |  |
| TF and TI | 13 | 29 |
| TF and/or TI | 53 | 246 |
| other |  |  |
| Pannus | 2 | 3 |
| Photography | 9 | 10 |
| Infection | 106 | 350 |
| Serology | 27 | 414 |
| **Total** | **235\*** | **1,775** |

*F2/F3: A grade of F2 or F3 (≥ 5 follicles) in the follicles, papillae, cicatrices (FPC) grading system; TI: trachomatous inflammation—intense; P2/P3: A grade of P2 or P3 in the FPC grading system, where P3 is equivalent to TI; WHO: World Health Organization. \*Column does not sum to total number of studies as each study may report one or more indicator types.*

### Outcomes

#### Comparison between field-graded TF and photo-graded TF in the same population

The mean difference in prevalence measured by field-graded vs. photo-graded TF was 0.7 percentage points, with a 95% CI of -15.2–16.7. The range of the absolute value of differences was (0, 17.6). Only one included study reported a population prevalence of TF by both field grading and photographs taken with an unaided smartphone,^69^ and this study reported a mean difference between the two methods of 14.8 percentage points. Six studies^8, 69, 197, 207, 217, 218^ reported a population prevalence of TF by both field grading and photographs taken with a DSLR camera, and the mean difference in these comparisons was 3.9 percentage points (95% CI:-8.1–15.9). The range of the absolute value of differences for the studies using a DSLR was (0.2, 17.6). The Bland-Altman plot visualizes these results (Supplemental File S8).

#### Comparison between TF and TI, infection, and serology pre-vs. post-initiation of MDA

The relationships between TF_1–9_ and TI prevalence, infection prevalence, seroprevalence, and SCR pre-vs. post-initiation of MDA are shown in Figures 1a and 1b. The comparison of TF_1–9_ and TI prevalence shows that the correlation increased post-initiation of MDA (from a weighted correlation of R^2^=0.65 to R^2^=0.81) and was statistically significant both pre- and post-initiation of MDA (p<0.001 in both settings). TF_1–9_ and infection were more strongly correlated pre-initiation of MDA (weighted R^2^=0.43, p=0.003) compared to post-initiation of MDA (weighted R^2^=0.002, p=0.788). TF_1–9_ and seroprevalence were more strongly correlated pre-initiation of MDA (weighted R^2^=0.56, p<0.001) compared to post-initiation of MDA (weighted R^2^=0.03, p=0.353). TF_1–9_ and SCR were more strongly correlated pre-initiation of MDA (weighted R^2^=0.52, p=0.012) compared to post-initiation of MDA (weighted R^2^=0.26, p=0.061).

**Figure 1a.**
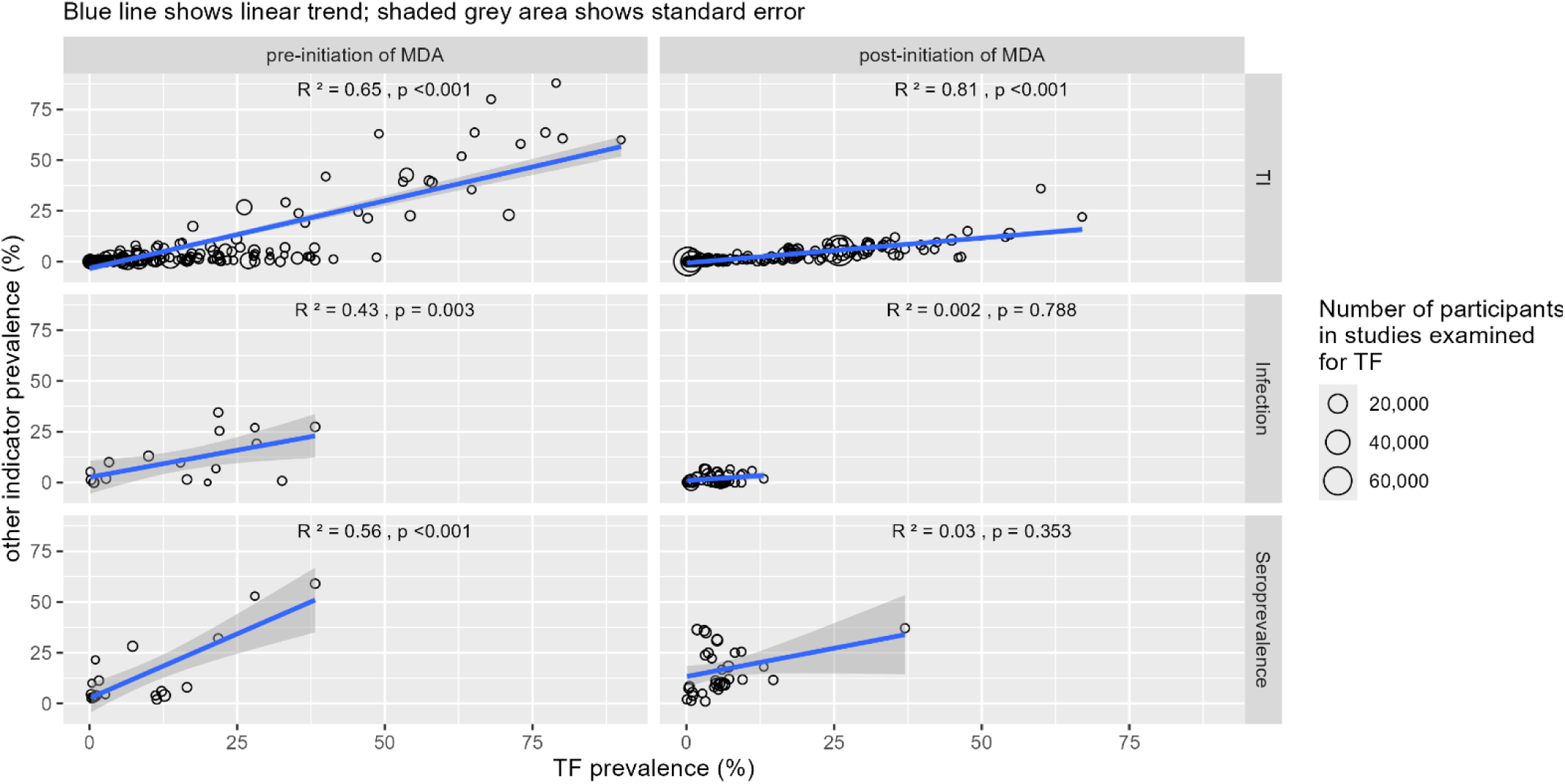
Trachomatous inflammation—follicular (TF) in 1–9-year-olds vs. other indicators, pre-vs. post-initiation of mass drug administration (MDA). Scatterplots of population-level prevalence estimates of TF vs. trachomatous inflammation—intense in 1–9-year-olds (TI), infection, and seroprevalence with weighted correlations. The blue line shows the linear trend; the shaded gray area shows standard error.

**Figure 1b.**
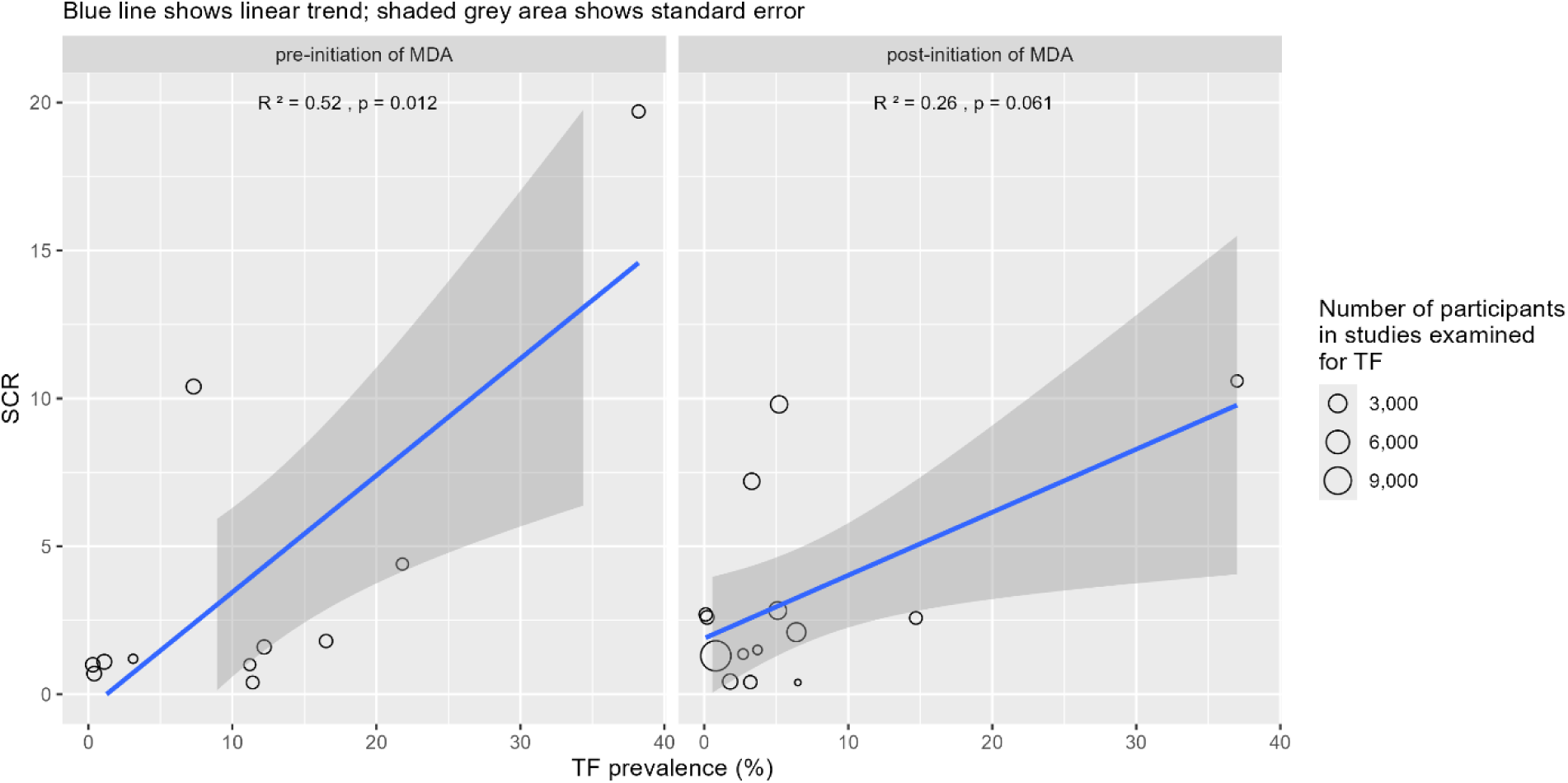
Trachomatous inflammation—follicular (TF) in 1–9-year-olds vs. Seroconversion rate (SCR), per 100 person-years, pre-vs. post-initiation of mass drug administration (MDA). Scatterplots of population-level prevalence estimates of TF vs. SCR with weighted correlations. The blue line shows the linear trend; the shaded gray area shows standard error.

#### Comparison between TI and infection pre-vs. post-initiation of MDA

The relationship between TI in 1–9-year-olds and infection pre-vs. post-initiation of MDA is shown in Supplemental File S9. While the sample size with TI and infection data is relatively small (a total of 13 comparisons), the correlation of TI prevalence vs. infection prevalence decreased post-initiation of MDA, with a weighted R^2^=0.46, p=0.063 pre-initiation of MDA and a weighted R^2^=0.37, p=0.273 post-initiation of MDA. In both MDA settings, the weighted correlation between TI and infection was higher than that between TF_1–9_ and infection.

#### Comparison between categorical TF_1–9_ and continuous measures of other indicators

Figure 2 shows the results of each continuous non-TF indicator (TI, infection, seroprevalence, and SCR, respectively) vs. categorical TF_1–9_,using the 5% programmatic threshold to categorize TF_1–9_. Within each pair of results in the figure rows, the left panel presents a boxplot and the right panel a histogram where the y-axis shows the frequency. Table 3 presents the median, IQR, minimum, and maximum of each comparison group for each non-TF indicator.

**Figure 2.**
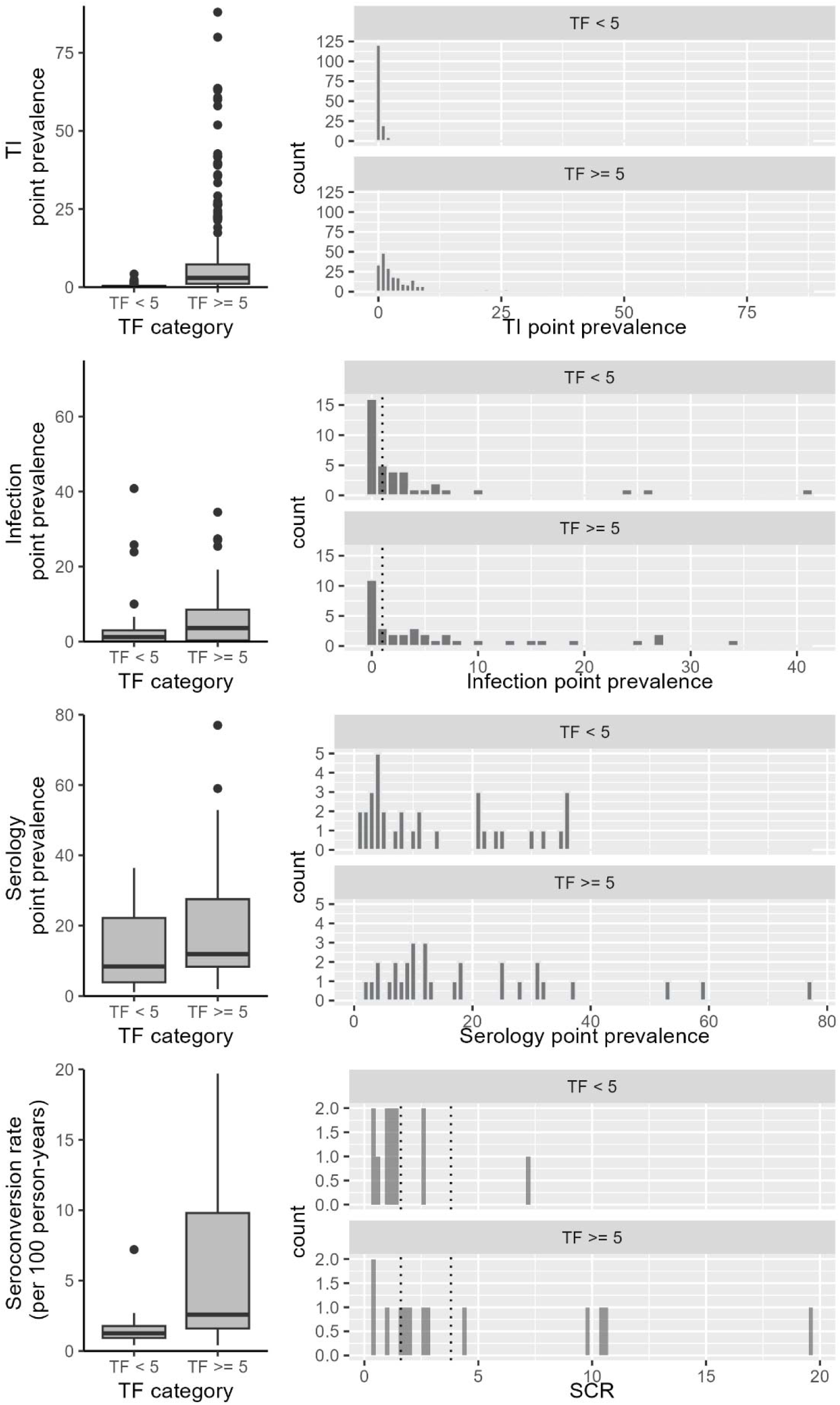
Indicator results by trachomatous inflammation—follicular (TF) in 1–9-year-olds categories. Left panels display boxplots by TF category (TF <5% vs. TF≥5%), right panels display histograms by TF category. Dotted lines represent threshold values of interest: Infection: the potential threshold of 1%, SCR: proposed thresholds of 1.6 and 3.8 per 100 person-years. TI: trachomatous inflammation— intense in 1–9-year-olds

**Table 3.** Summary statistics for continuous measures of complementary indicators with trachomatous inflammation—follicular (TF) in 1–9-year-olds above and below 5% threshold

|  | **TF category** | **minimum** | **Q1** | **median** | **Q3** | **maximum** |
| --- | --- | --- | --- | --- | --- | --- |
| **TI prevalence (%)** | TF <5% | 0.0 | 0.0 | 0.1 | 0.4 | 4.2 |
|  | TF ≥5% | 0.0 | 1.1 | 3.0 | 7.3 | 88.0 |
| **Ct infection prevalence (%)** | TF <5% | 0.0 | 0.1 | 1.2 | 3.0 | 40.8 |
|  | TF ≥5% | 0.0 | 0.3 | 3.6 | 8.5 | 34.5 |
| **Seroprevalence (%)** | TF <5% | 1.1 | 3.9 | 8.4 | 22.2 | 36.4 |
|  | TF ≥5% | 2.0 | 8.4 | 11.9 | 27.5 | 77.0 |
| **SCR (per 100 person-years)** | TF <5% | 0.4 | 0.9 | 1.3 | 1.8 | 7.2 |
|  | TF ≥5% | 0.4 | 1.6 | 2.6 | 9.8 | 19.7 |

Q1(3): first (third) quartile, TI: trachomatous inflammation—intense in 1–9-year-olds, *Ct*: *Chlamydia trachomatis*, SCR: seroconversion rate

#### TF_1–9_ category vs. complementary indicator category in surveys used for programmatic decision-making

A total of 45 discrete areas (either evaluation units or combinations of evaluation units) in 17 studies were identified as reporting infection and/or SCR data collected as part of surveys used for programmatic decision-making (Supplemental file S10). Twenty-five areas had a TF_1–9_ result warranting continued MDA (i.e., TF_1–9_ ≥5%). Of these, seven had at least one other complementary indicator that was “low”, and a further five areas had at least one other indicator that was “medium.” If the typical programmatic action (based solely upon the TF_1–9_ category) were adhered to, 10 areas may have had three unnecessary rounds of MDA and two areas may have had one additional round of MDA, resulting in a total of 32 unnecessary MDA rounds.

Table 4 presents, for the subset of surveys identified as being used for programmatic decision-making, the results of pairwise categorical comparisons between TF_1–9_, infection prevalence, and SCR in three different contexts: no history of MDA, unusual epidemiology, and post-initiation of MDA (including post-validation). In areas with no history of MDA, 100% of the TF_1–9_ vs. infection prevalence comparisons (5/5) and infection prevalence vs. SCR comparisons (2/2) were concordant and 86% of the TF_1–9_ vs. SCR comparisons (6/7) were concordant. In the areas with unusual epidemiology, 56% of the TF_1–9_ vs. infection prevalence comparisons (9/16), 38% of the TF_1–9_ vs. SCR comparisons (3/8), and 80% of the infection prevalence vs. SCR comparisons (4/5) were concordant. In the areas that were post-initiation of MDA, 67% of the TF_1–9_ vs. infection prevalence comparisons (8/12), 50% of the TF_1–9_ vs. SCR comparisons (3/6), and 67% of the infection prevalence vs. SCR comparisons (2/3) were concordant.

**Table 4:**
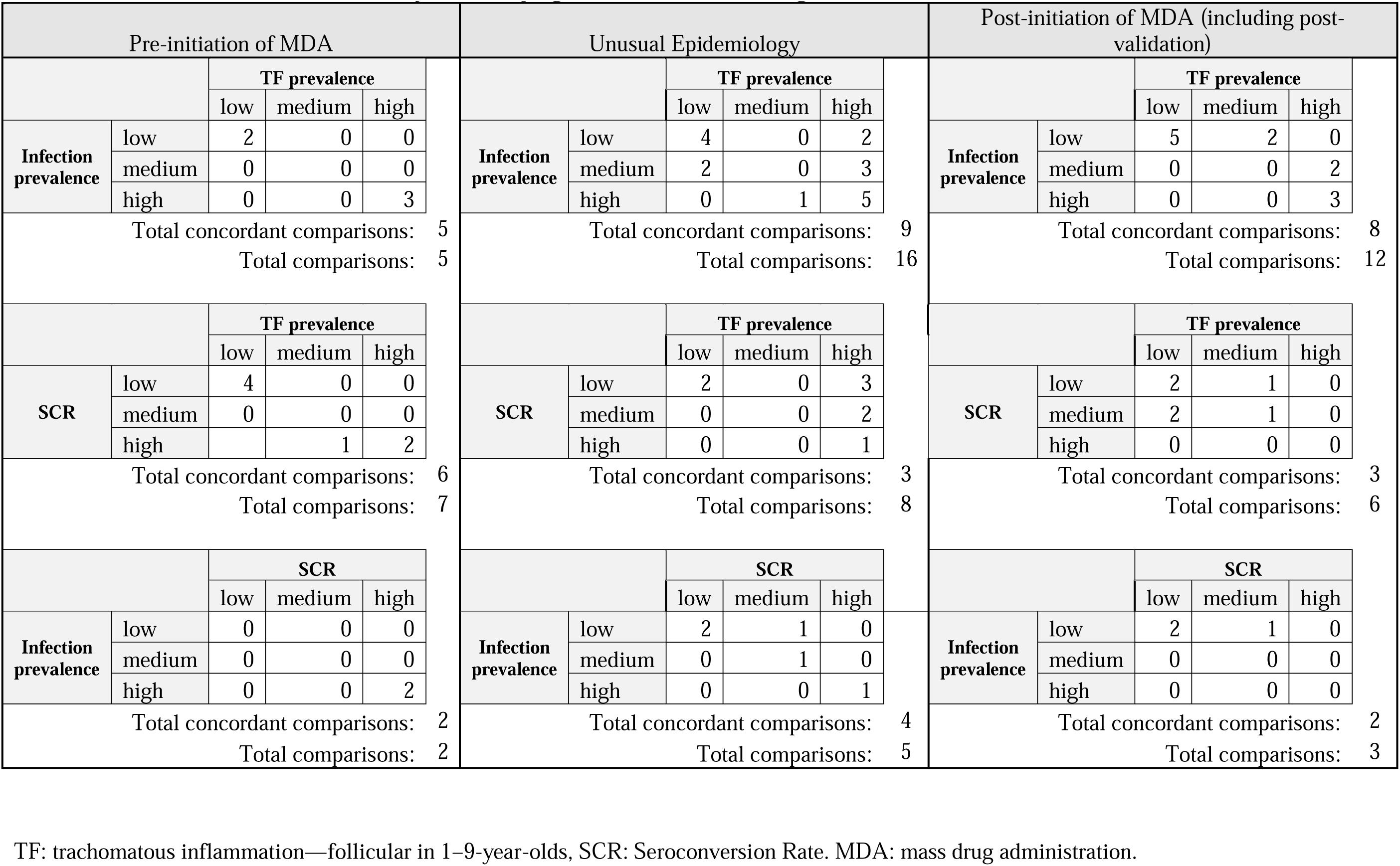
Concordance of indicators in surveys used for programmatic decision-making in different contexts

| Pre-initiation of MDA |  |  |  |  | Unusual Epidemiology |  |  |  |  | Post-initiation of MDA (including post-validation) |  |  |  |  |
| --- | --- | --- | --- | --- | --- | --- | --- | --- | --- | --- | --- | --- | --- | --- |
|  |  | TF prevalence |  |  |  |  | TF prevalence |  |  |  |  | TF prevalence |  |  |
|  |  | low | medium | high |  |  | low | medium | high |  |  | low | medium | high |
| **Infection prevalence** | low | 2 | 0 | 0 | **Infection prevalence** | low | 4 | 0 | 2 | **Infection prevalence** | low | 5 | 2 | 0 |
|  | medium | 0 | 0 | 0 |  | medium | 2 | 0 | 3 |  | medium | 0 | 0 | 2 |
|  | high | 0 | 0 | 3 |  | high | 0 | 1 | 5 |  | high | 0 | 0 | 3 |
| Total concordant comparisons: 5 |  |  |  |  | Total concordant comparisons: 9 |  |  |  |  | Total concordant comparisons: 8 |  |  |  |  |
| Total comparisons: 5 |  |  |  |  | Total comparisons: 16 |  |  |  |  | Total comparisons: 12 |  |  |  |  |
|  |  | TF prevalence |  |  |  |  | TF prevalence |  |  |  |  | TF prevalence |  |  |
|  |  | low | medium | high |  |  | low | medium | high |  |  | low | medium | high |
| **SCR** | low | 4 | 0 | 0 | **SCR** | low | 2 | 0 | 3 | **SCR** | low | 2 | 1 | 0 |
|  | medium | 0 | 0 | 0 |  | medium | 0 | 0 | 2 |  | medium | 2 | 1 | 0 |
|  | high |  | 1 | 2 |  | high | 0 | 0 | 1 |  | high | 0 | 0 | 0 |
| Total concordant comparisons: 6 |  |  |  |  | Total concordant comparisons: 3 |  |  |  |  | Total concordant comparisons: 3 |  |  |  |  |
| Total comparisons: 7 |  |  |  |  | Total comparisons: 8 |  |  |  |  | Total comparisons: 6 |  |  |  |  |
|  |  | SCR |  |  |  |  | SCR |  |  |  |  | SCR |  |  |
|  |  | low | medium | high |  |  | low | medium | high |  |  | low | medium | high |
| **Infection prevalence** | low | 0 | 0 | 0 | **Infection prevalence** | low | 2 | 1 | 0 | **Infection prevalence** | low | 2 | 1 | 0 |
|  | medium | 0 | 0 | 0 |  | medium | 0 | 1 | 0 |  | medium | 0 | 0 | 0 |
|  | high | 0 | 0 | 2 |  | high | 0 | 0 | 1 |  | high | 0 | 0 | 0 |
| Total concordant comparisons: 2 |  |  |  |  | Total concordant comparisons: 4 |  |  |  |  | Total concordant comparisons: 2 |  |  |  |  |
| Total comparisons: 2 |  |  |  |  | Total comparisons: 5 |  |  |  |  | Total comparisons: 3 |  |  |  |  |

TF: trachomatous inflammation—follicular in 1–9-year-olds, SCR: Seroconversion Rate. MDA: mass drug administration.

#### Comparison between infection and SCR pre-vs. post-initiation of MDA

Figure 3 presents comparisons of continuous measures of infection prevalence and SCR for the studies where all three measures were reported in the same populations (n=14 populations reported in 9 studies). Panel A presents the relationship between infection and SCR in all observations regardless of MDA history; in Panel B the observations are split by pre-vs. post-initiation of MDA. Panel A shows that the correlation between infection and SCR was significant (weighted R^2^ = 0.4, p=0.015); however, when split by MDA history, there was no correlation in pre-MDA settings (weighted R^2^=0.23, p=0.681), whereas the correlation in post-initiation of MDA settings was strong (weighted R^2^ = 0.71) and statistically significant (p=0.017). There were only three observations pre-MDA. All pre-MDA observations were from the Pacific Islands, while all post-MDA observations were from Africa.

**Figure 3.**
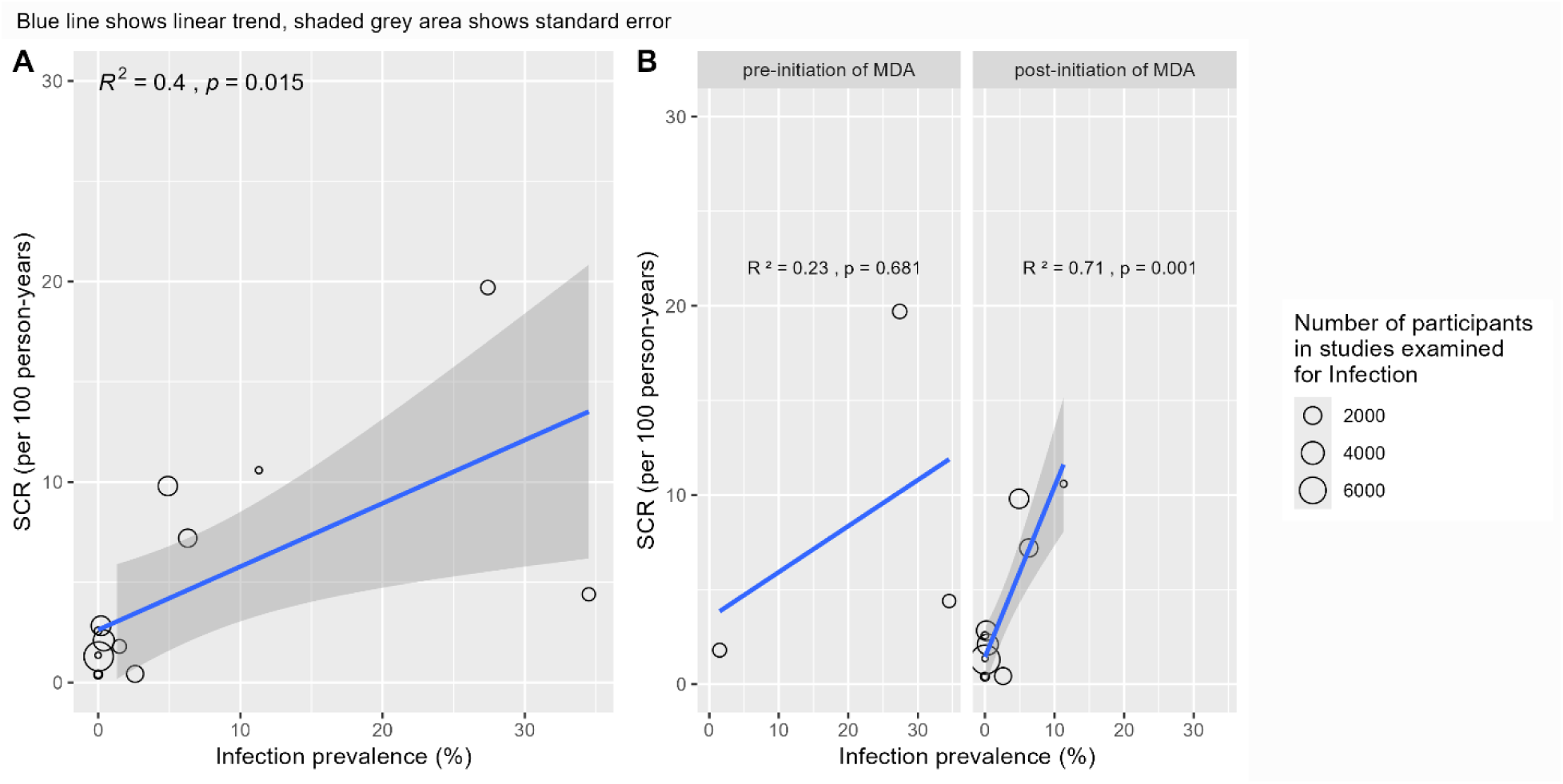
Comparisons of continuous measures of infection and seroconversion rate (SCR) for studies where both measures reported in the same population. Scatterplots of population-level infection prevalence estimates vs. SCR. The blue line shows the linear trend; the shaded gray area shows standard error. Panel A displays all observations, Panel B splits the observations according to mass drug administration (MDA) setting.

Comparisons of TF_1–9_ and infection prevalence and TF_1–9_ and SCR separately, for the populations with all three indicators reported are presented in Supplemental File S11. For each of the TF_1–9_ vs. infection prevalence and TF_1–9_ vs. SCR comparisons, the weighted correlations were moderate and significant (weighted R^2^: 0.49, p=0.006 and weighted R^2^: 0.54, p=0.003, respectively), although the sample size was small (n=14).

Table 5 presents categorical comparisons of measures of TF_1–9_, infection prevalence, and SCR for the studies in which all three measures were reported in the same populations (regardless of the context of the study or whether it was used for programmatic decision-making). Of 14 total comparisons with all three measures, 43% of comparisons of TF_1–9_ vs. infection prevalence (n=6), 57% of comparisons of TF_1–9_ and SCR (n=8), and 79% of comparisons of infection prevalence vs. SCR (n=11) were concordant.

**Table 5.** Categorical comparisons of measures of trachomatous inflammation—follicular in 1–9-year-olds (TF_1–9_), infection prevalence, and seroconversion rate (SCR)

|  |  | TF prevalence |  |  |
| --- | --- | --- | --- | --- |
|  |  | low | medium | high |
| prevalence | low | 3 | 3 | 1 |
|  | medium | 0 | 0 | 1 |
|  | high | 2 | 1 | 3 |
|  |  | low | medium | high |
| SCR | low | 4 | 2 | 0 |
|  | medium | 0 | 1 | 2 |
|  | high | 1 | 1 | 3 |

|  |  | SCR |  |  |
| --- | --- | --- | --- | --- |
|  |  | low | medium | high |
| Infection prevalence | low | 5 | 2 | 0 |
|  | medium | 0 | 1 | 0 |
|  | high | 1 | 0 | 5 |

### Risk of bias and study quality assessment

The mean quality score for studies assessed using the JBI Checklist for Prevalence Studies was 74%, and the mean quality score for studies assessed using the JBI Checklist for Diagnostic Test Accuracy was 83% (Supplemental File S6). Of the 220 studies assessed using the JBI Checklist for Prevalence Studies, 57% (125) achieved a score of 70% or higher; of the 42 studies assessed using the JBI Checklist for Diagnostic Test Accuracy, 79% (33) achieved a score of 70% or higher. When categorized by indicator, the mean quality scores were: 72% for studies reporting clinical indicators other than TF, 41% for studies reporting photo-graded TF, 62% for studies reporting infection, 79% for studies reporting seroprevalence, and 83% for studies reporting SCR. Due to the majority of the contributing studies being observational, a “low” GRADE rating for each outcome was given.

## Discussion

This review explored the relationship between TF and other indicators reported in 235 different studies over a 30-year period, including data from 49 countries and representing all trachoma-endemic WHO regions.

The comparisons of TF_1–9_ to *Ct* infection, seroprevalence, and SCR in pre-vs. post-MDA initiation settings showed a similar pattern for each indicator: in each analysis, the weighted correlation between TF and the other indicator was stronger pre-vs. post-initiation of MDA. A previous review comparing TF and infection found similar results.^7^ Given that the relationship deteriorates for each of the three pairwise comparisons with TF (TF vs. infection, TF vs. seroprevalence, and TF vs. SCR), it is likely that the indicator that has changed its performance post-initiation of MDA is TF. Thus, these results are strongly suggestive of poor performance of TF prevalence for guiding programs on decision-making after MDA has commenced. Conversely, infection and SCR were strongly and statistically significantly correlated post-initiation of MDA, although there were few studies containing data on TF_1–9_, infection, and SCR in the same population.

With a very small sample size, evidence suggests a strong correlation between TI and infection; however, given difficulties of reliably training graders, the low specificity of this clinical sign,^278^ and the reduction of the correlation evident in post-initiation of MDA settings, challenges would remain with using TI as an indicator of *Ct* transmission intensity.

When comparing results of TF_1–9_, infection prevalence, and SCR in the subset of studies reporting data used for programmatic decision-making, almost half (48%) of the areas where TF_1–9_ was above the threshold for MDA (TF_1–9_ ≥5%) had at least one other indicator with a result that was low or medium. Considering these results, populations for which programmatic decisions were based solely on TF_1–9_ may have proceeded with additional rounds of MDA (and later, trachoma impact surveys) that may not have been warranted based on infection and/or serology data. The additional cost of adding complementary indicators to standard trachoma surveys must be weighed against the potential additional costs of unnecessary rounds of MDA and subsequent surveys. Similar justification has been made on the cost-savings of trachoma impact surveys; a relatively costly activity, but if it demonstrates that MDA can be stopped it ultimately saves programs money.^279^

When comparing continuous measures of TI, infection, and serology to categorical TF_1–9_, in each comparison, the mean complementary indicator measure was higher for the higher TF_1–9_ category. Generally, when TF_1–9_ was under the 5% threshold, measures of infection and SCR tended to also be low; when TF_1–9_ was over the 5% threshold, measures of infection and SCR had a wider range of values (i.e., could be high or low). This suggests that making programmatic decisions based solely on TF_1–9_ has a propensity for “over-calling” (i.e., continuing MDA unnecessarily) rather than “under-calling” (i.e., stopping MDA prematurely) treatment decisions, resulting in unnecessary MDA rounds, unnecessary expense, and delaying validation of elimination of trachoma as a public health problem.

In the subset of populations that included TF, *Ct* infection, and SCR, the infection vs. SCR comparisons had a higher rate of concordant results compared to either of the comparisons with TF, which suggests SCR is a better marker for true new infections and a good estimate of the force of infection compared to TF. In the comparison of continuous infection and SCR data, the weighted correlation between these measures was strong and significant in post-initiation of MDA settings. And in studies reporting data used for programmatic decision-making, the concordance of infection and SCR was as high or higher than either the TF_1–9_ vs. infection or TF_1–9_ vs. SCR comparisons in all three settings. However, the sample size for these comparisons was small.

Recent attempts to produce a serological cutoff based on SCR acknowledge the existence of a gray area between proposed thresholds indicating high probability of elimination (or not) where additional measurements or context-specific consideration may be needed to determine future programmatic activity.^23^ Relatively few included studies that reported the use of serological indicators included an estimation of SCR, and even fewer included both SCR and infection data. Given the growing evidence that SCR has a potential use for programmatic decision-making,^23, 280, 281^ more collection and analysis of serological and infection data in a variety of trachoma settings would be beneficial. WHO has recommended that complementary indicator data be collected and used for programmatic decision-making in areas where trachoma is persistent or recrudescent.^282^ WHO also recently published a Target Product Profile for trachoma surveillance, which identified additional epidemiological contexts in which complementary data could be beneficial, including in newly suspected trachoma-endemic areas, after discontinuation of MDA, and in areas with high TF_1–9_ but little evidence of its sequela, TT, in adults.^283^ Work by Tropical Data to standardize the collection and analysis of infection and serology data as part of routine trachoma surveys is ongoing.^5^

Conjunctival photography is a potential solution to several problems with field-grading of signs of trachoma, including allowing review of grading decisions by outside experts, the adjudication of border cases by multiple graders,^284^ and the potential for machine learning technologies to remove human subjectivity and reduce overall error.^285^ Comparing field-graded TF to photo-graded TF, we found that the mean prevalence by each method was very close, but the 95% CIs were wide and crossed 0 (meaning that one type of grading did not consistently over- or underestimate the other). Despite different inclusion criteria and key outcome measures, these results are consistent with a systematic review published in 2021 showing high agreement between the two methods.^16^ However, only one included study in either review compared photos taken with an unaided smartphone against field grading. A recently published study showed very poor sensitivity of smartphones vs. field grading for TF and a high proportion (20%) of photos being ungradable due to poor photo quality.^286^ Thus, further work is needed to ensure accurate and reproducible smartphone photos in the field, and their grading. Key challenges remain in deploying the use of photography at-scale, including needing to develop minimum criteria for photographers, camera equipment, and image quality, as well as reducing the proportion of images that are ungradable and understanding the risk of bias posed by ungradable images.^17^ While routine use of the International Coalition for Trachoma Control’s manual for taking high quality field photos^287^ will help overcome some of these issues (such as decreasing the proportion of ungradable photos), the trachoma community has agreed that further work is needed before recommending the routine use of photos to replace field grading.^17, 285^

This review had several limitations. While mean quality scores of included studies were high, these varied by indicator, and the observational nature of the included studies conferred a “low” GRADE rating for each outcome, indicating that our confidence in the summary statistics output is limited. The wide scope yielded many studies to be screened for eligibility from a time period spanning over 30 years and there were studies where the full text was unable to be located, in part due to publication dates prior to widespread digitization; all care was taken to locate records wherever possible. A majority of these studies pre-date the use of serological detection platforms for trachoma,^288^ and did not reference data collection beyond clinical indicator types in their abstracts; therefore, the effect on the results for the non-clinical indicator types is likely minimal. The wide scope also led to a heterogeneous final dataset with different study types, purposes, and methodologies (including different grader training systems, different nucleic acid amplification tests, different serological assays, etc.) which led, in part, to the decision not to pursue a meta-analysis. In addition, we were unable to complete a full double data extraction; however, comparing full text extraction with a random sample of 20% of papers that were double extracted yielded a high degree of agreement, lending confidence to the accuracy of the full data extraction. The majority of the included studies in this review came from published literature, which introduces the risk of publication bias. In our case, there may be less of a risk of prevalence studies with results under programmatic thresholds remaining unpublished, as country programs must provide evidence that thresholds have been achieved in order to be validated by WHO as having achieved the elimination of trachoma as a public health problem.^22^ However, there is undoubtedly a risk of bias attributed to the non-standard application of complementary indicators, as it is probable that additional indicators (especially infection and serology) are more likely to be collected in areas with “unusual” epidemiology.

Furthermore, for infection data, in the absence of publications demonstrating a more rigorous analytical approach to determine proposed thresholds (as there is for SCR)^23, 281^, thresholds for this analysis were based on values proposed less formally, i.e., at an international meeting of stakeholders; the use of different thresholds could alter the results of the categorical analyses. In addition, while we extracted (and abstracted) data on whether settings had ever conducted MDA, we did not extract data on the timing of data collection post-MDA; as there is evidence that both infection and TF prevalence change over time after the cessation of MDA,^289^ the timing of the post-MDA data collection could influence the results of these indicators. Lastly, a number of comparisons had relatively small sample sizes; of note are those populations reporting both infection and SCR. There is a chance that some studies with these data did not meet the inclusion criteria if they did not also report field-graded TF. Due to the small number of studies with these data and lack of diversity in settings in the studies that were included, it is likely that outliers disproportionately affected these results.

Because categorical thresholds are particularly useful for programmatic decision-making, further work to propose meaningful thresholds for infection would be beneficial. Further studies should also look to better understand how each of the included indicators relate to longer-term sequelae of the disease: trachomatous scarring, TT, and eventual vision impairment and blindness.

## Conclusion

TF is not strongly correlated with other indicators of transmission of ocular *Ct* (i.e., infection prevalence and SCR) in post-MDA-initiation settings. With the global target of elimination of trachoma as a public health problem by 2030,^290^ and evidence that TF has a tendency to over-call transmission intensity, resulting in unnecessary costly and time-consuming additional MDA and survey rounds, the evidence supports inclusion of these indicators as part of the global strategy to achieving a world free from trachoma.

## Authors’ Contributions

KKR and EMHE conceived of the study; KKR, RB, TDH, and EMHE designed the study protocol; PJH, AWS, and DLM reviewed the study protocol; KKR performed the searches and KKR and CCL conducted both stages of screening; KKR and JLH extracted data; KKR conducted data analysis; KKR, RB, TDH, and EMHE conducted data interpretation; KKR prepared the original draft of the manuscript; all authors critically reviewed the manuscript and approved the final manuscript for publication.

## Data availability statement

All relevant data are within the manuscript, its Supporting Information files, and within the relevant references.

## Supplemental Material (list)

- S1: PICOST table
- S2: Search terms/search strategy
- S3: PRISMA checklist
- S4: PRISMA diagram
- S5: Missing full text by year of publication
- S6: Table of included studies
- S7: Analysis by region
- S8: Bland-Altman plots (field-graded vs photo-graded TF)
- S9: TI vs. infection, pre- vs. post-initiation of MDA
- S10: Table of studies reporting data used for programmatic decision-making
- S11: TF vs. infection, TF vs. SCR in subset of populations with all 3 indicators
- S12: Full dataset (long)

## Supporting information

S1 - PICOST framework

S2 - Search terms

S3 - PRISMA checklist

S4 - PRISMA diagram

S5 - Publication year of full texts

S6 - Table of included studies

S7 - Analysis by region

S8 - Bland Altman plot

S9 - TI vs. infection plot

S10 - Table of programmatic decisions

S11 - TF vs. infection, TF vs. SCR

## Abbreviations and acronyms

CI: Confidence interval
*Ct*: Chlamydia trachomatous
FPC: follicles papillae cicatrices
IQR: Interquartile range
JBI: Joanna Briggs Institute
MDA: Mass Drug Administration
SCR: seroconversion rate
TF: trachomatous inflammation—follicular
TI: trachomatous inflammation—intense
WHO: World Health Organization

## Footnotes

a TF and F2/F3 are not completely synonymous, as the F2 category denotes more than 5 follicles whereas TF denotes 5 or more follicles;^13^ however, for the purposes of this review, TF and its precursor F2 were considered interchangeable, and this marker will be referred to as “TF” throughout.

