## Supplementary material for "Comparison of methods for assessing *Chlamydia trachomatis* transmission intensity: a systematic review": S1 - PICOST framework

### Table – PICOST Framework

| | Term | Question | Response |
|---|---|---|---|
| **P** | Population or Problem | What are the characteristics of the Population or patient? What is the Problem, condition, or disease you are interested in? | Populations where TF has been diagnosed clinically and at least 1 alternative and/or complementary indicator |
| **I** | Intervention or Exposure | How do you wish to Intervene - what do you want to do with this patient - treat, diagnose, observe, etc.? | Diagnosis of trachoma via non-field-graded clinical TF or FPC (the precursor to TF) |
| **C** | Comparison | What is the Comparison or alternative to the intervention - placebo, different drug or therapy, surgery, etc.? | Diagnosis of trachoma via field-graded clinical TF or FPC (the precursor to TF) |
| **O** | Outcome | What are the possible Outcomes - morbidity, death, complications, etc.? | Prevalence of trachoma as measured by different indicators and correlations between different measures |
| **S** | Situation | Context or location | Populations in trachoma-endemic (or formerly-endemic) countries |
| **T** | Type of Study | Study type(s), such as systematic reviews, randomised control trials (RCTs), observational studies, economic evaluations, or others. | Randomized control trials, observational studies, longitudinal studies, cross-sectional studies |
