## Supplementary material for "Comparison of methods for assessing *Chlamydia trachomatis* transmission intensity: a systematic review": S2 - Search terms

1. Search in OVID (for Medline, Embase, and Global Health databases)

| | |
|---|---|
| 1 | exp Trachoma/ |
| 2 | exp Chlamydia trachomatis/ |
| 3 | trachoma.mp. [mp=title, book title, abstract, original title, name of substance word, subject heading word, floating sub-heading word, keyword heading word, organism supplementary concept word, protocol supplementary concept word, rare disease supplementary concept word, unique identifier, synonyms] |
| 4 | ocular trachoma.mp. [mp=title, book title, abstract, original title, name of substance word, subject heading word, floating sub-heading word, keyword heading word, organism supplementary concept word, protocol supplementary concept word, rare disease supplementary concept word, unique identifier, synonyms] |
| 5 | **1 or 2 or 3 or 4** |
| 6 | Polymerase Chain Reaction/ |
| 7 | Nucleic Acid Amplification Techniques/ |
| 8 | infect*.tw. |
| 9 | PCR.mp. [mp=title, book title, abstract, original title, name of substance word, subject heading word, floating sub-heading word, keyword heading word, organism supplementary concept word, protocol supplementary concept word, rare disease supplementary concept word, unique identifier, synonyms] |
| 10 | LCR.mp. [mp=title, book title, abstract, original title, name of substance word, subject heading word, floating sub-heading word, keyword heading word, organism supplementary concept word, protocol supplementary concept word, rare disease supplementary concept word, unique identifier, synonyms] |
| 11 | **6 or 7 or 8 or 9 or 10** |
| 12 | Enzyme-Linked Immunosorbent Assay/ |
| 13 | Serology/ |
| 14 | lateral flow assay.mp. |
| 15 | sero*.mp. [mp=title, book title, abstract, original title, name of substance word, subject heading word, floating sub-heading word, keyword heading word, organism supplementary concept word, protocol supplementary concept word, rare disease supplementary concept word, unique identifier, synonyms] |
| 16 | antibod*.mp. [mp=title, book title, abstract, original title, name of substance word, subject heading word, floating sub-heading word, keyword heading word, organism supplementary concept word, protocol supplementary concept word, rare disease supplementary concept word, unique identifier, synonyms] |
| 17 | **12 or 13 or 14 or 15 or 16** |
| 18 | Photography/ |
| 19 | photo*.mp. [mp=title, book title, abstract, original title, name of substance word, subject heading word, floating sub-heading word, keyword heading word, organism supplementary concept word, protocol supplementary concept word, rare disease supplementary concept word, unique identifier, synonyms] |
| 20 | image.mp. [mp=title, book title, abstract, original title, name of substance word, subject heading word, floating sub-heading word, keyword heading word, organism supplementary concept word, protocol supplementary concept word, rare disease supplementary concept word, unique identifier, synonyms] |

| | |
|---|---|
| 21 | images.mp. [mp=title, book title, abstract, original title, name of substance word, subject heading word, floating sub-heading word, keyword heading word, organism supplementary concept word, protocol supplementary concept word, rare disease supplementary concept word, unique identifier, synonyms] |
| 22 | imaging.mp. [mp=title, book title, abstract, original title, name of substance word, subject heading word, floating sub-heading word, keyword heading word, organism supplementary concept word, protocol supplementary concept word, rare disease supplementary concept word, unique identifier, synonyms] |
| 23 | **18 or 19 or 20 or 21 or 22** |
| 24 | (TI or TS or pannus or herbert*).mp. [mp=title, book title, abstract, original title, name of substance word, subject heading word, floating sub-heading word, keyword heading word, organism supplementary concept word, protocol supplementary concept word, rare disease supplementary concept word, unique identifier, synonyms] |
| 25 | **11 or 17 or 23 or 24** |
| 26 | **5 and 25** |

2. Scopus search: ( TITLE-ABS-KEY ( trachoma ) AND ( TITLE-ABS-KEY ( infect* OR pcr OR lcr ) OR TITLE-ABS-KEY ( sero* OR "lateral flow assay" OR antibod* ) OR TITLE-ABS-KEY ( photo* OR image OR images OR imaging ) OR TITLE-ABS-KEY ( ti OR ts OR pannus OR herbert* ) ) )

3. Global Index Medicus search: tw:((tw:(trachoma)) AND (tw:( infect* OR pcr OR lcr OR sero* OR "lateral flow assay" OR antibod* OR photo* OR image OR images OR imaging OR ti OR ts OR pannus OR herbert* )))

4. ETHoS Search: "trachoma"
