## Supplementary material for "Comparison of methods for assessing *Chlamydia trachomatis* transmission intensity: a systematic review": S4 - PRISMA diagram

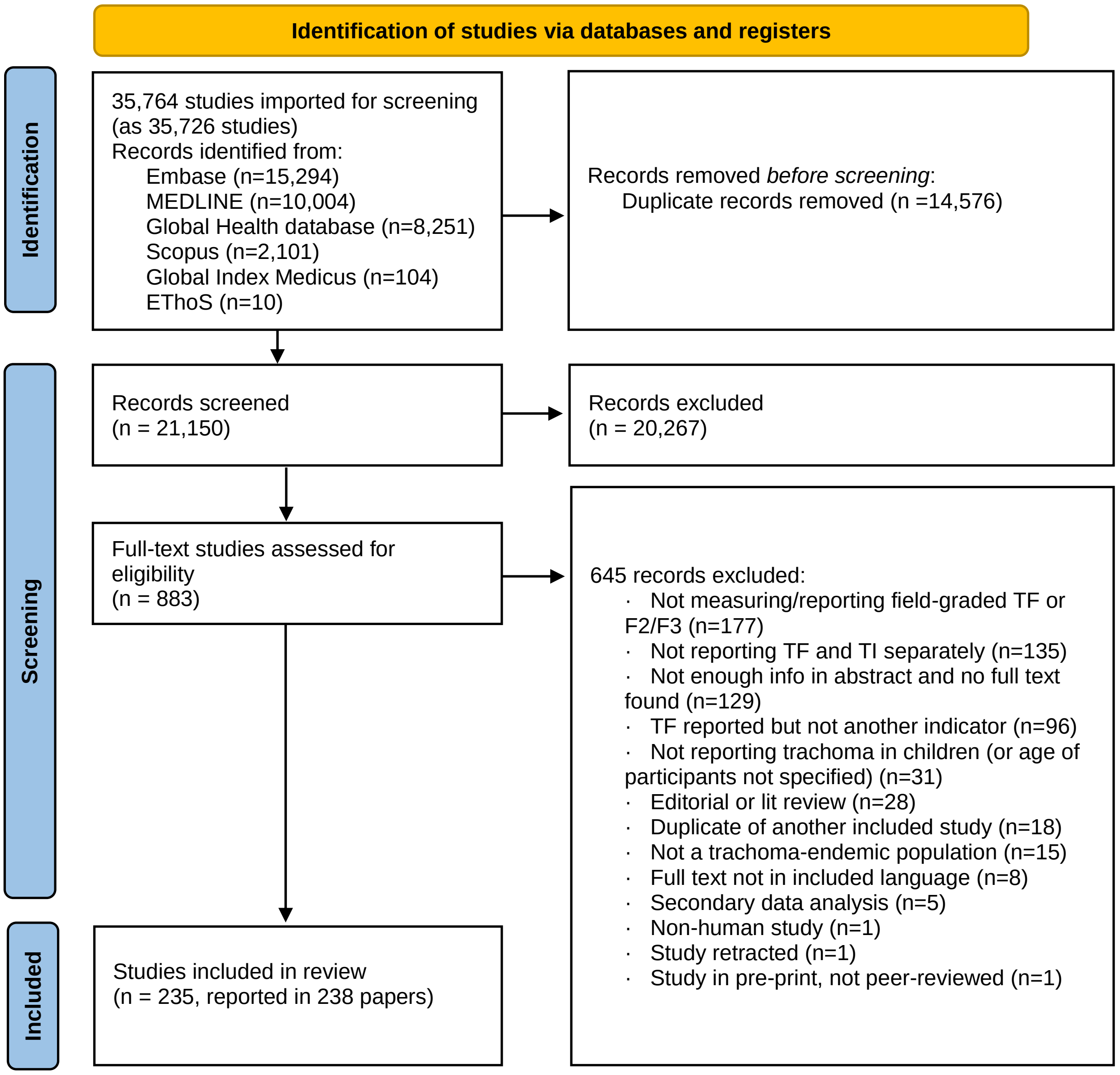


Source: Page MJ, et al. BMJ 2021;372:n71. doi: 10.1136/bmj.n71.

This work is licensed under CC BY 4.0. To view a copy of this license, visit https://creativecommons.org/licenses/by/4.0/
