## Supplementary material for "Comparison of methods for assessing *Chlamydia trachomatis* transmission intensity: a systematic review": S5 - Publication year of full texts

Analysis of publication year for full text articles that could not be found (n=93)

| Year of publication | Count of Published Year | running total | running percent |
|---|---|---|---|
| **1987** | 1 | 1 | 1% |
| **1988** | 3 | 4 | 4% |
| **1989** | 6 | 10 | 11% |
| **1990** | 6 | 16 | 17% |
| **1991** | 5 | 21 | 23% |
| **1992** | 12 | 33 | 35% |
| **1993** | 2 | 35 | 38% |
| **1994** | 3 | 38 | 41% |
| **1995** | 4 | 42 | 45% |
| **1996** | 3 | 45 | 48% |
| **1997** | 5 | 50 | 54% |
| **1999** | 3 | 53 | 57% |
| **2000** | 2 | 55 | 59% |
| **2001** | 4 | 59 | 63% |
| **2003** | 1 | 60 | 65% |
| **2004** | 2 | 62 | 67% |
| **2005** | 1 | 63 | 68% |
| **2007** | 4 | 67 | 72% |
| **2009** | 3 | 70 | 75% |
| **2010** | 1 | 71 | 76% |
| **2011** | 2 | 73 | 78% |
| **2012** | 1 | 74 | 80% |
| **2013** | 4 | 78 | 84% |
| **2014** | 3 | 81 | 87% |
| **2015** | 1 | 82 | 88% |
| **2016** | 4 | 86 | 92% |
| **2017** | 3 | 89 | 96% |
| **2018** | 1 | 90 | 97% |
| **2019** | 1 | 91 | 98% |
| **2020** | 2 | 93 | 100% |
