## Supplementary material for "Comparison of methods for assessing *Chlamydia trachomatis* transmission intensity: a systematic review": S6 - Table of included studies

| First author, year (reference number*) | Country or countries of data collection | Type of study | TF or FPC | other clinical indicators? | photo-graded TF or FPC? | infection? |
|---|---|---|---|---|---|---|
| Abdilwohab, 2020 (1) | Ethiopia | Cross-sectional | TF | TI | no | no |
| Abdou, 2007 (2) | Niger | Cross-sectional | TF | TI | no | yes |
| Abebo, 2017 (3) | Ethiopia | Cross-sectional | TF | TI | no | no |
| Adamu, 2018 (4) | Ethiopia | Cross-sectional | TF | TI, TF and/or TI | no | no |
| Adera, 2016 (5) | Ethiopia | Cross-sectional | TF | TI | no | no |
| Al-Arab, 2001 (6) | Egypt | Cross-sectional | TF | TI | no | no |
| Alemayehu, 2005 (7) | Ethiopia | Cross-sectional | TF | TI, TF and TI, TF and/or TI | no | no |
| Amer, 2018 (8) | Egypt | Cross-sectional | TF | TI | no | no |
| Amza, 2010 (9) | Cameroon | Cross-sectional | TF | TF and/or TI | no | no |
| Amza, 2012 (10) | Niger | Cross-sectional | TF | TF and/or TI | no | yes |
| Amza, 2013 (11) | Niger | Cross-sectional | TF | no | no | yes |
| Amza, 2017 (12) | Niger | Cross-sectional within RCT | TF | TI | no | yes |
| Amza, 2018 (13) | Niger | Cross-sectional within RCT | TF | no | no | yes |
| Amza, 2019 (14) | Niger | Cross-sectional within RCT | TF | TI | no | yes |
| Astle, 2006 (15) | Zambia | Cross-sectional | TF | TI | no | no |

| | | | | | | |
|---|---|---|---|---|---|---|
| Ayele, 2011 (16) | Ethiopia | Cross-sectional | TF | TI, TF and/or TI | no | yes |
| Ayelgn, 2021 (17) | Ethiopia | Cross-sectional | TF | TI | no | no |
| Ayena, 2010 (18) | Togo | Cross-sectional | TF | TI | no | no |
| Ayena, 2011 (19) | Togo | Cross-sectional | TF | TI | no | no |
| Babalola, 2005 (20) | Nigeria | Cross-sectional within RCT | TF | TI | no | no |
| Bailey, 1994 (21) | Gambia | Cross-sectional | any follicles | no | no | yes |
| Bamani, 2010 (22) | Mali | Cross-sectional | TF | TI | no | no |
| Bamani, 2010 (23) | Mali | Cross-sectional | TF | TI | no | no |
| Bejiga, 2001 (24) | Ethiopia | Cross-sectional | TF | TI | no | no |
| Bella, 2020 (25) | Cameroon | Cross-sectional | TF | TI | no | no |
| Berhane, 2007 (26) | Ethiopia | Cross-sectional | TF | TI, TF and/or TI | no | no |
| Bhosai, 2012 (27) | Niger | Cross-sectional | TF | TI | yes | no |
| Bid, 2013 (28) | Tanzania | Cross-sectional | TF | TI | no | no |
| Bio, 2017 (29) | Benin | Cross-sectional | TF | TI | no | no |
| Bobo, 1991 (30) | Tanzania | Cross-sectional | TF | TI | no | yes |
| Brady, 2021 (31) | Tanzania | Cross-sectional | TF | no | yes | no |
| Burr, 2013 (32) | Gambia | Cross-sectional | TF | no | no | yes |
| Burr, 2019 (33) | Malawi | Cross-sectional | TF | no | no | yes |
| Burton, 2010 (34) | Gambia | Cross-sectional within longitudinal | TF | no | no | yes |
| Burton, 2011 (35) | Tanzania | Cross-sectional | TF | TI | no | yes |
| Butcher, 2016 (36) | Solomon Islands | Cross-sectional | TF | TI | no | yes |

| | | | | | | |
|---|---|---|---|---|---|---|
| Butcher, 2017 (37) | Fiji | Cross-sectional | TF | TI | no | no |
| Butcher, 2018 (38) | Solomon Islands | Cross-sectional | TF | TI | no | yes |
| Butcher, 2020 (39) | Kiribati, Vanuatu | Cross-sectional | TF | TI | no | yes |
| Caligaris, 2006 (40) | Brazil | Cross-sectional | TF | TI | no | no |
| Cama, 2017 (41) | Kiribati | Cross-sectional | TF | TI | no | yes |
| Chen, 2022 (42) | Tanzania | Cross-sectional within longitudinal | TF | TI | no | yes |
| Cochereau, 2007 (43) | Guinea and Pakistan | Cross-sectional within RCT | TF | TF and/or TI | no | no |
| Cocks, 2016 (44)/Macleod, 2016 (45) | Fiji | Cross-sectional | TF | TI | no | yes |
| Cox, 2017 (46) | Tanzania | Cross-sectional | TF | no | no | yes |
| Cromwell, 2014 (47) | Niger | Cross-sectional | TF | TI | no | no |
| Cumberland, 2005 (48) | Ethiopia | Cross-sectional | TF | TI | no | no |
| Cumberland, 2008 (49) | Ethiopia | Cross-sectional | TF | no | no | yes |
| Damasceno, 2009 (50) | Brazil | Cross-sectional | TF | TI | no | no |
| Datta, 1994 (51) | Kenya | Cross-sectional | TF | TI | no | yes |
| Debrah, 2017 (52) | Ghana | Cross-sectional | TF | TI | no | no |
| Derrick, 2016 (53) | Guinea Bissau | Case-control | TF | Pannus | no | yes |
| Derrick, 2016 (54) | Guinea Bissau | Case-control | TF | P3 (FPC) | no | yes |
| Derrick, 2020 (55) | Tanzania | Cross-sectional | TF | P3 (FPC) | no | yes |
| de Sousa Meneghim, 2018 (56) | Brazil | Cross-sectional | TF | TI | no | no |
| Dorkenoo, 2012 (57) | Togo | Cross-sectional | TF | TI | no | no |

| | | | | | | |
|---|---|---|---|---|---|---|
| Duale, 2018 (58) | Ethiopia | Cross-sectional | TF | TI | no | no |
| Edwards, 2012 (59) | South Sudan | Cross-sectional | TF | TI | no | no |
| Ejigu, 2013 (60) | Ethiopia | Cross-sectional | TF | TI | no | no |
| Ervin, 2016 (61) | Tanzania | Cross-sectional | TF | TI, TF and TI | no | yes |
| Faal, 2005 (62) | Gambia | Cross-sectional | TF | TF and TI | no | yes |
| Faal, 2006 (63) | Gambia | Cross-sectional | TF | TI | no | yes |
| Favacho, 2018 (64) | Brazil | Cross-sectional | TF | TI | no | no |
| Faye, 2006 (65) | Senegal | Cross-sectional | TF | TI, TF and TI | no | no |
| Felton, 2003 (66) | Gambia | Case-control | TF | TI | no | no |
| Feng, 2017 (67) | China | Cross-sectional | TF | TI, <5 follicles | no | no |
| Ferede, 2017 (68) | Ethiopia | Cross-sectional | TF | TI | no | no |
| Freitas, 2016 (69) | Brazil | Cross-sectional | TF | TI | no | no |
| Gelaye, 2014 (70) | Ethiopia | Cross-sectional within longitudinal | TF | TI | no | no |
| Genet, 2022 (71) | Ethiopia | Cross-sectional | TF | TI | no | no |
| Ghasemian, 2018 (72) | Sudan | Case-control | TF | no | no | yes |
| Ghasemian, 2021 (73) | Morocco | Case-control | TF | no | no | yes |
| Goldschmidt, 2006 (74) | Pakistan | Cross-sectional | TF | TI | no | yes |
| Goldschmidt, 2012 (75) | Cameroon | Cross-sectional | TF | TI | no | yes |
| Goldschmidt, 2014 (76) | Cameroon | Cross-sectional | TF | TF and TI | no | no |
| Golovaty, 2009 (77) | Ethiopia | Cross-sectional | TF | TI, TF and/or TI | no | no |
| Gower, 2006 (78) | Gambia, Tanzania | Cross-sectional | TF | TI | no | yes |

| | | | | | | |
|---|---|---|---|---|---|---|
| Gwyn, 2018 (79) | Nepal | Cross-sectional | TF | no | no | no |
| Gwyn, 2019 (80) | Tanzania | Cross-sectional | TF | no | no | yes |
| Gwyn, 2021 (81) | Democratic Republic of the Congo, Togo | Cross-sectional | TF | no | no | no |
| Gwyn, 2021 (82)/Nash, 2021 (83) | Ethiopia | Cross-sectional | TF | TI | no | yes |
| Gyasi, 2010 (84) | Ghana | Cross-sectional | TF | TI | no | no |
| Hagan, 2009 (85) | Ghana | Cross-sectional | 2 or more follicles | TF and TI | no | no |
| Haileselassie, 2007 (86) | Ethiopia | Cross-sectional | TF | TI, TF and/or TI | no | no |
| Hammou, 2022 (87) | Morocco | Cross-sectional | TF | TI, TF and/or TI | no | no |
| Harding-Esch, 2008 (88) | Gambia | Cross-sectional | TF | TI, TF and/or TI | no | yes |
| Harding-Esch, 2009 (89) | Gambia | Cross-sectional | TF | TI | no | no |
| Harding-Esch, 2010 (90) | Gambia | Cross-sectional | TF | TI, TF and TI | no | yes |
| Harding-Esch, 2013 (91) | Gambia | Cross-sectional within longitudinal | TF | no | no | yes |
| Harding-Esch, 2017 (92) | Senegal | Cross-sectional | TF | TI | no | no |
| Harding-Esch, 2019 (93) | Senegal | Cross-sectional | TF | TI | no | yes |
| Hudson, 1992 (94) | unknown | Cross-sectional | TF | TI | no | yes |
| Huguet, 2010 (95) | Cameroon | Cross-sectional | TF | TF and/or TI, TF and TI | no | no |
| Jenson, 2013 (96) | Tanzania | Cross-sectional | TF | TI, TF and TI | no | yes |
| Jip, 2008 (97) | Nigeria | Cross-sectional | TF | TI, TF and/or TI | no | no |
| Kabona, 2013 (98) | Tanzania | Cross-sectional | TF | TI | no | no |

| | | | | | | |
|---|---|---|---|---|---|---|
| Kalua, 2010 (99) | Malawi | Cross-sectional | TF | TI | no | no |
| Kalua, 2014 (100) | Malawi | Cross-sectional | TF | TI | no | no |
| Kashaf, 2020 (101) | Tanzania | Cross-sectional within longitudinal | TF | no | no | yes |
| Kassim, 2019 (102) | Ethiopia | Cross-sectional | TF | TI, TF and/or TI, TF and TI | no | no |
| Kasubi, 2020 (103) | Tanzania | Cross-sectional | TF | no | no | yes |
| Katz, 1996 (104) | Nepal | Cross-sectional | TF | TI | no | no |
| Kedir, 2021 (105) | Ethiopia | Cross-sectional | TF | TI, TF and/or TI, TF and TI | no | no |
| Keenan, 2012 (106) | Ethiopia | Cross-sectional within RCT | TF | TI, TF and/or TI | no | yes |
| Keenan, 2018 (107) | Ethiopia | Cross-sectional within RCT | TF | no | no | yes |
| Ketema, 2012 (108) | Ethiopia | Cross-sectional | TF | TI, TF and/or TI | no | no |
| Khandekar, 2003 (109) | Oman | Case-control | TF | TI | no | no |
| Khandekar, 2005 (110) | Oman | Cross-sectional | TF | TI | no | no |
| Khanduja, 2009 (111) | India | Cross-sectional | TF | TI, TF and/or TI | no | yes |
| Kim, 2019 (112) | Niger | Cross-sectional | TF | TI | no | no |
| King, 2008 (113) | South Sudan | Cross-sectional | TF | TI, TF and/or TI | no | no |
| King, 2010 (114) | Nigeria | Cross-sectional | TF | TI, TF and/or TI | no | no |
| King, 2013 (115) | Ethiopia, Mali, Niger, Nigeria | Cross-sectional | TF | TI | no | no |
| King, 2014 (116) | Ethiopia | Cross-sectional | TF | TI | no | no |

| | | | | | | |
|---|---|---|---|---|---|---|
| Koroma, 2011 (117) | Sierra Leone | Cross-sectional | TF | TF and/or TI | no | no |
| Koukounari, 2011 (118) | Tanzania | Cross-sectional | TF | TI | no | yes |
| Kur, 2009 (119) | South Sudan | Cross-sectional | TF | TI | no | no |
| Laming, 1999 (120) | Australia | Cross-sectional | TF | no | no | yes |
| Last, 2014 (121)/Roberts, 2013 (122) | Guinea Bissau | Cross-sectional | TF | TI, TF and/or TI | no | yes |
| Last, 2015 (123) | Guinea Bissau | Cross-sectional within RCT | TF | no | no | yes |
| Last, 2017 (124) | Guinea Bissau | Cross-sectional | TF | TI | no | yes |
| Lee, 2013 (125) | Tanzania | Cross-sectional | TF | no | no | yes |
| Lee, 2014 (126) | Tanzania | Cross-sectional | TF | TI | no | yes |
| Liang, 2016 (127) | China | Cross-sectional | TF | TI | no | yes |
| Liu, 2015 (128) | Niger | Cross-sectional within longitudinal | TF | TI | no | yes |
| Luna, 2016 (129) | Brazil | Cross-sectional | TF | TI | no | no |
| Lynch, 2022 (130) | Australia | Cross-sectional | TF | <5 follicles | no | yes |
| Lynch, 2022 (131) | Nauru | Cross-sectional | TF | no | no | yes |
| Lynch, 2022 (132) | Australia | Cross-sectional | TF | <5 follicles | no | yes |
| Mabey, 1991 (133) | Gambia | Case-control | F2/F3 | P3, Pannus | no | no |
| Macleod, 2019 (134) | Sudan | Cross-sectional | TF | TI | no | no |
| Macleod, 2020 (135) | Papua New Guinea | Cross-sectional | TF | no | no | yes |
| Madani, 2003 (136) | Chad | Cross-sectional | TF | TI | no | no |
| Malhotra, 2016 (137) | India | Cross-sectional | TF | TI | no | no |
| Martin, 2015 (138) | Tanzania | Cross-sectional | TF | no | no | yes |

| | | | | | | |
|---|---|---|---|---|---|---|
| Martin, 2015 (139) | Tanzania | Cross-sectional | TF | TI | no | yes |
| Mathew, 2009 (140) | Fiji, Kiribati, Nauru, Solomon Islands, Vanuatu | Cross-sectional | TF | TI | no | no |
| Medina, 1998 (141) | Brazil | Cross-sectional | TF | TF and/or TI | no | no |
| Meng, 2016 (142) | Cambodia | Cross-sectional | TF | TI | no | no |
| Mesfin, 2006 (143) | Ethiopia | Cross-sectional | TF | TI | no | no |
| Michel, 2006 (144) | Tanzania | Cross-sectional | TF | no | no | yes |
| Michel, 2011 (145) | Australia | Cross-sectional | TF | no | no | yes |
| Migchelsen, 2017 (146) | Gambia, Laos, Uganda | Cross-sectional | TF | TI | no | no |
| Miller, 2020 (147) | Colombia | Cross-sectional | TF | TI | no | no |
| Miller, 2004 (148) | Ethiopia | Cross-sectional | TF | TI, TF and/or TI, TF and TI | no | yes |
| Miller, 2010 (149) | Colombia | Cross-sectional | TF | TI | no | no |
| Missamou, 2018 (150) | Republic of the Congo | Cross-sectional | TF | TI | no | no |
| Mkocha, 2009 (151) | Tanzania | Cross-sectional | TF | TI | no | yes |
| Mohammadi, 2017 (152) | Iran | Cross-sectional | TF | no | no | yes |
| Mpyet, 2008 (153) | Nigeria | Cross-sectional | TF | TI, TF and/or TI | no | no |
| Mpyet, 2010 (154) | Nigeria | Cross-sectional | TF | TI, TF and/or TI | no | no |
| Mpyet, 2012 (155) | Nigeria | Cross-sectional | TF | TI | no | no |
| Muhammad, 2018 (156) | Nigeria | Cross-sectional | TF | TI | no | no |
| Munoz, 2011 (157) | Tanzania | Cross-sectional | TF | no | yes | yes |

| | | | | | | |
|---|---|---|---|---|---|---|
| Nash, 2017 (158) | Ethiopia | Cross-sectional | TF | TI | no | yes |
| Nash, 2018 (159) | Ethiopia | Cross-sectional | TF | TI | no | yes |
| Nash, 2018 (160) | Ethiopia | Cross-sectional | TF | no | no | yes |
| Nash, 2020 (161) | Ethiopia | Cross-sectional | TF | TI | no | yes |
| Nash, 2022 (162) | Ethiopia | Cross-sectional | TF | TI | no | yes |
| Natividad, 2010 (163) | Gambia | Cross-sectional | F2/F3 | P3 | no | yes |
| Naufal, 2021 (164) | Tanzania | Cross-sectional | TF | no | yes | no |
| Ndisabiye, 2020 (165) | Burundi | Cross-sectional | TF | TI, TF and/or TI | no | no |
| Negrel, 1992 (166) | Morocco | Cross-sectional | TF | TF and/or TI | no | no |
| Nesemann, 2020 (167) | Ethiopia | Cross-sectional | TF | TI | yes | no |
| Nesemann, 2021 (168) | Peru | Cross-sectional | TF | no | no | yes |
| Ngondi, 2005 (169) | South Sudan | Cross-sectional | TF | TI, TF and/or TI | no | no |
| Ngondi, 2006 (170) | South Sudan | Cross-sectional | TF | TI, TF and/or TI | no | no |
| Ngondi, 2006 (171) | South Sudan | Cross-sectional | TF | TI, TF and/or TI | no | no |
| Ngondi, 2008 (172) | Ethiopia | Cross-sectional | TF | TI | no | no |
| Ngondi, 2009 (173) | Ethiopia | Cross-sectional | TF | TI, TF and/or TI | no | yes |
| Ngondi, 2010 (174) | Ethiopia | Cross-sectional | TF | TI | no | no |
| Nigusie, 2015 (175) | Ethiopia | Cross-sectional | TF | TI, TF and/or TI | no | no |

| | | | | | | |
|---|---|---|---|---|---|---|
| Noatina, 2013 (176) | Cameroon | Cross-sectional | TF | TF and/or TI | no | no |
| Noatina, 2014 (177) | Cameroon | Cross-sectional | TF | TF and/or TI | no | no |
| Odonkor, 2021 (178) | Tanzania | Cross-sectional | TF | no | yes | yes |
| Odonkor, 2022 (179) | Tanzania | Cross-sectional | TF | no | yes | no |
| Oldenburg, 2018 (180) | Niger | Cross-sectional within longitudinal | TF | no | no | yes |
| Oswald, 2017 (181) | Ethiopia | Cross-sectional | TF | TI, TF and/or TI | no | no |
| Phiri, 2018 (182) | Zimbabwe | Cross-sectional | TF | TI | no | no |
| Quicke, 2013 (183) | Gambia | Cross-sectional | TF | TI | no | no |
| Ramadhani, 2019 (184) | Tanzania | Cross-sectional within longitudinal | TF, F2/F3 | TI, P2/P3 | yes | yes |
| Ramyil, 2015 (185) | Nigeria | Cross-sectional | TF | TI | no | no |
| Reda, 2020 (186) | Ethiopia | Cross-sectional | TF | TI, TF and/or TI, TF and TI | no | no |
| Reilly, 2007 (187) | Brazil | Cross-sectional | TF | TI, TF and/or TI | no | no |
| Roba, 2013 (188) | Ethiopia | Cross-sectional | TF | TI, TF and/or TI | no | no |
| Rosen, DS, 1995 (189) | Yemen | Cross-sectional | TF | TI | no | no |
| Ruberanziza, 2009 (190) | Rwanda | Cross-sectional | TF | TI | no | no |
| Saboya-Diaz, 2022 (191) | Peru | Cross-sectional | TF | no | no | yes |
| Sanders, 2017 (192) | South Sudan | Cross-sectional | TF | TI, TF and/or TI | no | no |

| | | | | | | |
|---|---|---|---|---|---|---|
| Sanders, 2019 (193) | Sudan | Cross-sectional | TF | TI | no | no |
| Sanders, 2019 (194) | Sudan | Cross-sectional | TF | TI | no | no |
| Schellini, 2010 (195) | Brazil | Cross-sectional | TF | TI | no | no |
| Schemann, 1998 (196) | Mali | Cross-sectional | TF | TI | no | no |
| See, 2011 (197) | Ethiopia | Cross-sectional | TF | TI | no | yes |
| Senyonjo, 2018 (198) | Ghana | Cross-sectional | TF | no | no | yes |
| Shaffi, 2005 (199) | Ethiopia | Cross-sectional | TF | TI, TF and/or TI | no | no |
| Sheehan, 2018 (200) | Ethiopia | Cross-sectional | TF | no | no | yes |
| Shekhawat, 2014 (201) | Tanzania | Cross-sectional within longitudinal | TF | no | no | yes |
| Shimelash, 2022 (202) | Ethiopia | Cross-sectional | TF | TI, TF and/or TI | no | no |
| Silva, 2016 (203) | Brazil | Cross-sectional | TF | TI | no | no |
| Smith, 2007 (204) | Ethiopia | Cross-sectional | TF | TI, TF and/or TI | no | yes |
| Snyder, 2019 (205) | Burkina Faso | Cross-sectional | TF | TI | yes | no |
| Solomon, 2003 (206) | Tanzania | Cross-sectional within longitudinal | TF | TI, TF and/or TI | no | yes |
| Solomon, 2008 (207) | Tanzania | Cross-sectional within longitudinal | TF | no | no | yes |
| Stare, 2011 (208) | Gambia, Tanzania | Cross-sectional within longitudinal | TF | no | no | yes |
| Stewart, 2019 (209) | Ethiopia | Cross-sectional | TF | TI, TF and/or TI | no | no |
| Tadesse, 2017 (210) | Ethiopia | Cross-sectional within longitudinal | TF | TI, TF and/or TI | no | no |
| Tadesse, 2017 (211) | Ethiopia | Cross-sectional | TF | TI | no | no |

| | | | | | | |
|---|---|---|---|---|---|---|
| Vashist, 2013 (212) | India | Cross-sectional | TF | TI, TF and/or TI | no | no |
| Wang, 2016 (213) | China | Case-control | TF | no | no | yes |
| West, 1993 (214) | Tanzania | Cross-sectional | TF | TI | no | yes |
| West, 2001 (215) | Tanzania | Case-control | TF | TI | no | yes |
| West, 2007 (216) | Tanzania | Cross-sectional within longitudinal | TF | TI | no | yes |
| West, 2011 (217) | Tanzania | Cross-sectional within longitudinal | TF | TI | no | yes |
| West, 2013 (218) | Tanzania | Cross-sectional within longitudinal | TF | no | no | yes |
| West, 2015 (219) | Tanzania | Cross-sectional within longitudinal | TF | no | no | yes |
| West, 2016 (220) | Tanzania | Cross-sectional | TF | no | no | yes |
| West, 2017 (221) | Tanzania | Cross-sectional within longitudinal | TF | no | no | yes |
| West, 2017 (222) | Nepal | Cross-sectional | TF | no | no | yes |
| West, 2018 (223) | Tanzania | Cross-sectional | TF | no | no | yes |
| West, 2019 (224) | Tanzania | Cross-sectional | TF | no | no | yes |
| West, 2020 (225) | Tanzania | Cross-sectional | TF | no | no | yes |
| Wilson, 2019 (226) | Tanzania | Cross-sectional within longitudinal | TF | no | no | yes |
| Woldekidan, 2019 (227) | Ethiopia | Cross-sectional | TF | TI, TF and/or TI | no | no |
| Worku, 2002 (228) | Ethiopia | Cross-sectional | TF | TI | no | no |

| | | | | | | |
|---|---|---|---|---|---|---|
| Xue, 2016 (229) | China | Cross-sectional | TF | TI | no | no |
| Yang, 2007 (230) | Ethiopia | Cross-sectional | TF | TI, TF and/or TI | no | yes |
| Yang, 2009 (231) | Ethiopia | Cross-sectional | TF | TI, TF and/or TI | no | yes |
| Yaya, 2015 (232) | Central African Republic | Cross-sectional | TF | TI | no | no |
| Yayemain, 2009 (233) | Ghana | Cross-sectional | TF | TI, TF and/or TI | no | no |
| Yilkal, 2004 (234) | Ethiopia | Cross-sectional | TF | TI | no | no |
| Yohannan, 2013 (235) | Tanzania | Cross-sectional within longitudinal | TF | no | no | yes |
| Zambrano, 2016 (236) | Nepal | Cross-sectional | TF | no | no | yes |
| Zambrano, 2017 (237) | Tanzania | Cross-sectional within longitudinal | TF | TI, TF and/or TI | no | yes |
| Zerihun, 1997 (238) | Ethiopia | Cross-sectional | TF | TI, TF and/or TI | no | no |

TF: trachomatous inflammation--follicular, as measured by the World Health Organization (WHO) Simplified G
TI: trachomatous inflammation--intense, as measured by the WHO Simplified Grading System
RCT: randomised control trial
FPC: follicles, papillae, cicatrices system
SCR: seroconversion rate
DTA: Diagnostic Test Accuracy
*reference number refers to 'references' tab within this workbook, may not match reference number in man
**from the Joanna Briggs Institute (https://jbi.global/critical-appraisal-tools)

| serology ? | SCR? | Critical Appraisal Tool used** | Prevalence study: Was the sample frame appropriate to address the target pop? | Prevalence study: Were study participants sampled in an appropriate way? | Prevalence study: Was the sample size adequate? |
|---|---|---|---|---|---|
| no | no | Checklist for Prevalence Studies | Yes | Yes | Yes |
| no | no | Checklist for Prevalence Studies | Yes | Yes | Yes |
| no | no | Checklist for Prevalence Studies | Yes | Yes | Yes |
| no | no | Checklist for Prevalence Studies | Yes | Yes | Yes |
| no | no | Checklist for Prevalence Studies | Yes | Yes | No |
| no | no | Checklist for Prevalence Studies | Yes | Yes | Yes |
| no | no | Checklist for Prevalence Studies | Yes | Yes | Yes |
| no | no | Checklist for Prevalence Studies | Yes | Yes | Yes |
| no | no | Checklist for Prevalence Studies | Yes | Yes | Yes |
| no | no | Checklist for Prevalence Studies | Yes | Yes | Yes |
| no | no | Checklist for Prevalence Studies | Yes | Yes | Unclear |
| no | no | Checklist for Prevalence Studies | Yes | Yes | Unclear |
| no | no | Checklist for Prevalence Studies | Yes | Yes | Unclear |
| no | no | Checklist for Prevalence Studies | Yes | Yes | Unclear |
| no | no | Checklist for Prevalence Studies | Yes | Unclear, Not sure how villages were chosen for inclusion | Unclear |

| | | | | | |
|---|---|---|---|---|---|
| no | no | Checklist for Prevalence Studies | Yes | Yes | Yes |
| no | no | Checklist for Prevalence Studies | Yes | Yes | Yes |
| no | no | Checklist for Prevalence Studies | Yes | Unclear how villages/children chosen and if this was random. | Unclear |
| no | no | Checklist for Prevalence Studies | Yes | Unclear how villages/children chosen and if this was random. | Unclear |
| no | no | Checklist for Prevalence Studies | Yes | Unclear, villages chosen on basis of proximity to black fly breeding sites. | Unclear |
| no | no | Checklist for Prevalence Studies | Yes | Yes | Yes |
| no | no | Checklist for Prevalence Studies | Yes | Yes | Yes |
| no | no | Checklist for Prevalence Studies | Yes | Yes | Yes |
| no | no | Checklist for Prevalence Studies | Yes | Yes | Yes |
| no | no | Checklist for Prevalence Studies | Yes | Yes | Yes |
| no | no | Checklist for Prevalence Studies | Yes | Yes | Unclear |
| no | no | Both | Yes | Yes | Yes |
| no | no | Checklist for Prevalence Studies | Unclear | Unclear | Unclear |
| no | no | Checklist for Prevalence Studies | Yes | Yes | Yes |
| no | no | Checklist for Prevalence Studies | Yes | Yes | Unclear |
| no | no | Both | Unclear | Unclear | Unclear |
| no | no | Both | Yes | Yes | Yes |
| yes | yes | Both | Yes | Yes | Yes |
| no | no | Checklist for Prevalence Studies | Yes | Yes | Unclear |
| no | no | Both | Yes | Yes | Unclear |
| no | no | Checklist for Prevalence Studies | Yes | Yes | Yes |

| | | | | | |
|---|---|---|---|---|---|
| no | no | Checklist for Prevalence Studies | Yes | Yes | Yes |
| yes | no | Both | Unclear; only villages with previous TF rates >10% were chosen | Yes | Yes |
| yes | yes | Both | Yes | Yes | Yes |
| no | no | Checklist for Prevalence Studies | Yes | Yes | Yes |
| yes | no | Checklist for Prevalence Studies | Yes | Yes | Yes |
| yes | yes | Checklist for Prevalence Studies | Yes | Yes | Unclear |
| no | no | Checklist for Prevalence Studies | No | Unclear | Unclear |
| yes | no | Checklist for Prevalence Studies | Yes | Yes | Yes |
| no | no | Checklist for Prevalence Studies | Yes | Unclear | Unclear |
| no | no | Checklist for Prevalence Studies | Yes | Yes | Yes |
| no | no | Checklist for Prevalence Studies | Yes | Yes | Unclear |
| no | no | Checklist for Prevalence Studies | Yes | Yes | Unclear |
| no | no | Checklist for Prevalence Studies | Yes | Yes | Yes |
| no | no | Checklist for DTA studies | | | |
| no | no | Checklist for Prevalence Studies | Yes | Yes | Yes |
| no | no | Checklist for DTA studies | | | |
| no | no | Checklist for DTA studies | | | |
| no | no | Checklist for DTA studies | | | |
| no | no | Checklist for Prevalence Studies | No (what about private schools?) | Yes | Yes |
| no | no | Checklist for Prevalence Studies | Yes | Unclear | Unclear |

| | | | | | |
|---|---|---|---|---|---|
| no | no | Checklist for Prevalence Studies | Yes | Yes | Unclear |
| no | no | Checklist for Prevalence Studies | Yes | Yes | Unclear |
| no | no | Checklist for Prevalence Studies | Yes | Unclear - convenience sampling of sub-districts as per TRA methodology | Yes |
| no | no | Checklist for Prevalence Studies | Yes | Yes | Unclear |
| no | no | Checklist for DTA studies | | | |
| no | no | Checklist for DTA studies | | | |
| no | no | Checklist for Prevalence Studies | Yes | Yes | Unclear |
| no | no | Checklist for Prevalence Studies | Yes | Yes | Yes |
| no | no | Checklist for DTA studies | | | |
| no | no | Checklist for Prevalence Studies | Yes | Unclear | Unclear |
| no | no | Checklist for Prevalence Studies | Yes | Yes | Yes |
| no | no | Checklist for Prevalence Studies | Yes | Unclear | Unclear |
| no | no | Checklist for Prevalence Studies | Yes | Yes | Unclear |
| no | no | Checklist for Prevalence Studies | Yes | Yes | Yes |
| no | no | Checklist for DTA studies | | | |
| no | no | Checklist for DTA studies | | | |
| no | no | Checklist for Prevalence Studies | Yes | Unclear | Unclear |
| no | no | Checklist for Prevalence Studies | Yes | Yes | Yes |
| no | no | Checklist for Prevalence Studies | Yes | Yes | Yes |
| no | no | Checklist for Prevalence Studies | Yes | Yes | Yes |
| no | no | Both | Yes | Yes | Yes |

| | | | | | |
|---|---|---|---|---|---|
| yes | no | Checklist for Prevalence Studies | Yes | Yes | Yes |
| yes | no | Checklist for Prevalence Studies | Yes | Yes | Unclear |
| yes | yes | Checklist for Prevalence Studies | Yes | Yes | Unclear |
| yes | yes | Checklist for Prevalence Studies | Yes | Yes | Unclear |
| no | no | Checklist for Prevalence Studies | Yes | Yes | Yes |
| no | no | Checklist for Prevalence Studies | Yes | Yes | Unclear |
| no | no | Checklist for Prevalence Studies | Yes | Yes | Yes |
| yes | yes | Checklist for Prevalence Studies | Yes | Yes | Yes |
| no | no | Checklist for Prevalence Studies | Yes | Yes | Yes |
| no | no | Checklist for Prevalence Studies | Yes | Yes | Yes |
| no | no | Both | Yes | Yes | Unclear |
| no | no | Checklist for Prevalence Studies | Yes | Yes | Yes |
| no | no | Checklist for Prevalence Studies | Yes | Yes | Yes |
| no | no | Checklist for Prevalence Studies | Yes | Yes | Yes |
| no | no | Checklist for DTA studies | | | |
| no | no | Checklist for Prevalence Studies | Yes | Yes | Yes |
| no | no | Checklist for DTA studies | | | |
| no | no | Checklist for Prevalence Studies | Yes | Yes | Yes |
| no | no | Checklist for Prevalence Studies | Yes | Yes | Unclear |

| | | | | | |
|---|---|---|---|---|---|
| no | no | Checklist for Prevalence Studies | Yes | Yes | Yes |
| no | no | Checklist for Prevalence Studies | Yes | Yes | Yes |
| no | no | Checklist for Prevalence Studies | Yes | Yes | Yes |
| no | no | Checklist for Prevalence Studies | Yes | Yes | Yes |
| yes | no | Checklist for Prevalence Studies | Yes | Unclear | Unclear |
| no | no | Checklist for Prevalence Studies | Yes | Yes | Unclear |
| no | no | Checklist for Prevalence Studies | Yes | Yes | Yes |
| no | no | Both | Yes | Yes | Yes |
| no | no | Checklist for Prevalence Studies | Yes | Yes | Yes |
| no | no | Checklist for Prevalence Studies | Yes | Yes | Unclear |
| no | no | Checklist for Prevalence Studies | Yes | Yes | Yes |
| no | no | Checklist for Prevalence Studies | No | No | Unclear |
| no | no | Checklist for Prevalence Studies | No, using TRA methods, sampled most-likely to be positive areas | Yes | Yes |
| yes | no | Both | Yes | Yes | Unclear |
| no | no | Checklist for Prevalence Studies | Yes | Yes | Yes |
| no | no | Checklist for Prevalence Studies | Yes | Yes | Yes |
| no | no | Checklist for Prevalence Studies | Yes | Yes | Unclear |
| no | no | Checklist for Prevalence Studies | Yes | Yes | Yes |

| | | | | | |
|---|---|---|---|---|---|
| no | no | Checklist for Prevalence Studies | Yes | Yes | Yes |
| no | no | Checklist for Prevalence Studies | Unclear | Unclear | Unclear |
| no | no | Checklist for Prevalence Studies | Yes | Yes | Yes |
| no | no | Checklist for DTA studies | | | |
| no | no | Checklist for Prevalence Studies | Yes | Yes | Yes |
| no | no | Checklist for Prevalence Studies | Unclear | Unclear | Unclear |
| no | no | Checklist for Prevalence Studies | Yes | Yes | Yes |
| no | no | Checklist for Prevalence Studies | Unclear | Unclear | Unclear |
| no | no | Checklist for Prevalence Studies | Yes | Yes | Unclear |
| no | no | Checklist for Prevalence Studies | Unclear | Yes | Yes |
| no | no | Checklist for Prevalence Studies | Yes | Yes | Yes |
| no | no | Checklist for Prevalence Studies | Unclear, school-based | Yes | Yes |
| yes | no | Both | Yes | Unclear | Yes |
| yes | yes | Both | Yes | Yes | Yes |
| no | no | Checklist for Prevalence Studies | Yes | Yes | Yes |
| no | no | Checklist for Prevalence Studies | Unclear | Unclear | Unclear |
| no | no | Checklist for Prevalence Studies | Yes | Yes | Yes |
| yes | yes | Checklist for Prevalence Studies | Yes | Yes | Yes |
| no | no | Checklist for Prevalence Studies | Unclear | Unclear | Unclear |
| no | no | Checklist for Prevalence Studies | Yes | Yes | Yes |
| yes | no | Checklist for Prevalence Studies | Unclear | Unclear | Unclear |

| | | | | | |
|---|---|---|---|---|---|
| yes | yes | Checklist for Prevalence Studies | Yes | Yes | Yes |
| no | no | Checklist for Prevalence Studies | Yes | No, TRA | No |
| no | no | Checklist for Prevalence Studies | Yes | Yes | Yes |
| no | no | Checklist for Prevalence Studies | Yes | Yes | Yes |
| no | no | Checklist for Prevalence Studies | Yes | Yes | Unclear |
| no | no | Both | Yes | Unclear | Unclear |
| no | no | Checklist for Prevalence Studies | Yes | Yes | Yes |
| yes | yes | Both | Yes | Yes | Unclear |
| no | no | Checklist for Prevalence Studies | Yes | Yes | Unclear |
| no | no | Checklist for DTA studies | | | |
| no | no | Checklist for Prevalence Studies | Yes | Yes | No |
| no | no | Checklist for Prevalence Studies | Yes | Yes | Yes |
| no | no | Checklist for Prevalence Studies | Yes | Yes | Yes |
| no | no | Checklist for Prevalence Studies | Yes | No, only children w/ red eye screened for trachoma | Yes |
| no | no | Checklist for Prevalence Studies | Yes | Yes | Unclear |
| no | no | Checklist for Prevalence Studies | Yes | Yes | Yes |
| no | no | Checklist for Prevalence Studies | Yes | Yes | Yes |
| no | no | Checklist for Prevalence Studies | Yes | Yes | Yes |
| no | no | Both | Yes | Yes | Yes |

| | | | | | |
|---|---|---|---|---|---|
| no | no | Checklist for Prevalence Studies | Yes | Unclear | Unclear |
| no | no | Checklist for Prevalence Studies | Yes | Yes | Unclear |
| no | no | Checklist for Prevalence Studies | Yes | Yes | Unclear |
| no | no | Checklist for Prevalence Studies | Yes | Yes | Unclear |
| no | no | Checklist for Prevalence Studies | Yes | Yes | Unclear |
| no | no | Checklist for DTA studies | | | |
| no | no | Both | Unclear | Unclear | Unclear |
| no | no | Checklist for Prevalence Studies | Unclear, convenience sample from clinic | Unclear | Yes |
| no | no | Checklist for Prevalence Studies | Yes | Yes | Yes |
| no | no | Both | Yes | Yes | No |
| no | no | Checklist for Prevalence Studies | Yes | Yes | Unclear |
| no | no | Checklist for Prevalence Studies | Yes | Yes | Yes |
| no | no | Checklist for Prevalence Studies | Yes | Yes | Unclear |
| no | no | Checklist for Prevalence Studies | Yes | Yes | Yes |
| no | no | Checklist for Prevalence Studies | Yes | Yes | Yes |
| no | no | Checklist for Prevalence Studies | Yes | Yes | Unclear |
| no | no | Checklist for Prevalence Studies | Yes | Yes | Unclear |
| no | no | Checklist for Prevalence Studies | Yes | Yes | Yes |

| | | | | | |
|---|---|---|---|---|---|
| no | no | Checklist for Prevalence Studies | Yes | Yes | No |
| no | no | Checklist for Prevalence Studies | Unclear | Yes | Yes |
| yes | no | Both | Yes | Yes | Yes |
| no | no | Both | Unclear | Unclear | Unclear |
| no | no | Checklist for Prevalence Studies | Yes | Yes | Unclear |
| no | no | Checklist for Prevalence Studies | Yes | Yes | Unclear |
| no | no | Checklist for Prevalence Studies | Yes | Yes | Yes |
| no | no | Checklist for Prevalence Studies | Yes | Yes | Unclear |
| no | no | Checklist for Prevalence Studies | Yes | Unclear | Unclear |
| no | no | Checklist for Prevalence Studies | Yes | Yes | Yes |
| no | no | Checklist for Prevalence Studies | Yes | Yes | Yes |
| no | no | Checklist for Prevalence Studies | Yes | Unclear | Unclear |
| no | no | Checklist for Prevalence Studies | No, excluded communities receiving more frequent MDAs as part of other intervention studies | Yes | Yes |
| no | no | Checklist for Prevalence Studies | Unclear | Yes | Unclear |
| no | no | Checklist for Prevalence Studies | Yes | Yes | Yes |
| no | no | Checklist for Prevalence Studies | Yes | Yes | Yes |
| no | no | Checklist for Prevalence Studies | Yes | Yes | Yes |

| | | | | | |
|---|---|---|---|---|---|
| no | no | Checklist for Prevalence Studies | Yes | Yes | Yes |
| no | no | Checklist for Prevalence Studies | Yes | Yes | Yes |
| no | no | Checklist for Prevalence Studies | No, school-based | Yes | Yes |
| no | no | Checklist for Prevalence Studies | Yes | Yes | Unclear |
| no | no | Both | Yes | Yes | Unclear |
| yes | yes | Both | Yes | Yes | Unclear |
| no | no | Checklist for Prevalence Studies | Yes | Yes | Yes |
| no | no | Checklist for Prevalence Studies | Yes | Yes | Unclear |
| no | no | Checklist for Prevalence Studies | Yes | Yes | Unclear |
| no | no | Checklist for Prevalence Studies | Yes | Yes | Yes |
| no | no | Checklist for Prevalence Studies | No, school based | Unclear | Yes |
| no | no | Both | No, only includes children w/ a relative who had TT surgery | Not for a prevalence study | Unclear |
| no | no | Checklist for DTA studies | | | |
| no | no | Both | Yes | Yes | Yes |
| no | no | Checklist for Prevalence Studies | Yes | Yes | Yes |
| no | no | Checklist for Prevalence Studies | Yes | Yes | Unclear |
| no | no | Checklist for Prevalence Studies | Yes | Yes | Unclear |
| no | no | Checklist for Prevalence Studies | No, purposively sampled based on water status | Yes | Yes |
| no | no | Checklist for Prevalence Studies | Yes | Yes | Yes |

| | | | | | |
|---|---|---|---|---|---|
| no | no | Checklist for Prevalence Studies | No, purposive (TRA) | Yes | Unclear |
| no | no | Checklist for Prevalence Studies | No, school based | Unclear | Unclear |
| no | no | Checklist for Prevalence Studies | Yes | Unclear | Unclear |
| no | no | Checklist for Prevalence Studies | No, case control | Unclear | Unclear |
| no | no | Checklist for Prevalence Studies | Yes | Yes | Yes |
| no | no | Checklist for Prevalence Studies | Yes | Yes | Yes |
| no | no | Checklist for Prevalence Studies | Yes | Unclear | Yes |
| no | no | Checklist for Prevalence Studies | Yes | Yes | Unclear |
| yes | no | Checklist for Prevalence Studies | Yes | Yes | Yes |
| no | no | Checklist for Prevalence Studies | Yes | Yes | Unclear |
| yes | no | Checklist for Prevalence Studies | Yes | Yes | Yes |
| yes | yes | Checklist for Prevalence Studies | Yes | Yes | Yes |
| no | no | Checklist for Prevalence Studies | Yes | Yes | Yes |
| yes | yes | Checklist for Prevalence Studies | Yes | Yes | Yes |
| yes | no | Checklist for Prevalence Studies | Yes | Yes | Yes |
| no | no | Checklist for Prevalence Studies | Yes | Yes | Yes |
| no | no | Checklist for Prevalence Studies | No, school based | Yes | Unclear |

| | | | | | |
|---|---|---|---|---|---|
| no | no | Checklist for Prevalence Studies | Yes | Unclear | Unclear |
| no | no | Both | Yes | Unclear | Unclear |
| no | no | Both | Yes | Yes | Unclear |
| no | no | Checklist for Prevalence Studies | Yes | Yes | Yes |
| no | no | Checklist for Prevalence Studies | Unclear | Unclear | Unclear |
| no | no | Checklist for Prevalence Studies | Yes | Yes | Yes |
| no | no | Checklist for Prevalence Studies | Yes | Yes | Yes |
| yes | no | Checklist for Prevalence Studies | Yes | Yes | Unclear |
| no | no | Both | Yes | Yes | Yes |
| no | no | Checklist for Prevalence Studies | Yes | Yes | Yes |

rading System

uscript

| Prevalence study: Were the study subjects and setting described in detail? | Prevalence study: Was the data analysis conducted with sufficient coverage of the identified sample? | Prevalence study: Were valid methods used for the identification of the condition(s)? | Prevalence study: Was the condition measured in a standard, reliable way for all participants? | Prevalence study: Was there appropriate statistical analysis? |
|---|---|---|---|---|
| Yes | Yes | Yes | Yes | Yes |
| No | Yes | Yes | Yes | Yes |
| Yes | Yes | Yes | Yes | Yes |
| Yes | Unclear | Yes | Yes | Yes |
| Yes | Yes | Yes | Yes | Yes |
| No | Yes | Yes | Yes | Yes |
| Yes | Yes | Yes | Yes | Yes |
| No | Yes | Yes | Yes | Yes |
| Yes | Yes | Yes | Yes | Yes |
| Yes | Yes | Yes | Yes | Yes |
| No | Unclear | Yes | Yes | Yes |
| Yes | Unclear | Yes | Yes | Yes |
| Yes | Unclear | Yes | Yes | Yes |
| Yes | Unclear | Yes | Yes | Yes |
| Yes | Unclear | Yes | Yes | Yes |

| | | | | |
|---|---|---|---|---|
| Yes | Yes | Yes | Yes | Yes |
| No | Yes | Yes | Yes | Yes |
| Yes | Unclear | Yes | Yes | Yes |
| No | Unclear | Yes | Yes | Yes |
| No | Yes | Yes | Yes | Yes |
| No | Yes | Yes | Yes | Yes |
| Yes | Yes | Yes | Yes | Yes |
| Yes | Unclear | Yes | Yes | Yes |
| Yes | Yes | Yes | Yes | Yes |
| Yes | Yes | Yes | Yes | Yes |
| No | Unclear | Yes | Yes | Yes |
| Yes | Yes | Yes | Yes | Unclear |
| No | Unclear | Unclear | Unclear | Unclear |
| No | Yes | Yes | Yes | Yes |
| No | Unclear | Yes | Yes | Yes |
| Unclear | Unclear | Unclear | Unclear | Unclear |
| Yes | Yes | Yes | Yes | Yes |
| No | Unclear | Yes | Yes | Yes |
| No | Yes | Yes | Yes | Yes |
| No | Yes | Yes | Yes | Yes |
| Yes | Yes | Yes | Yes | Yes |

| | | | | |
|---|---|---|---|---|
| Yes | Unclear | Yes | Yes | Yes |
| Yes | Yes | Yes | Yes | Yes |
| Yes | Yes | Yes | Yes | Yes |
| No | Yes | Yes | Yes | Yes |
| No | Yes | Yes | Yes | Yes |
| Yes | Yes | Yes | Yes | Yes |
| Yes | Unclear | Yes | Yes | Yes |
| Yes | Yes | Yes | Yes | Yes |
| No | Unclear | Unclear | Unclear | Unclear |
| No | Yes | Yes | Yes | Yes |
| No | Unclear | Yes | Yes | Yes |
| Yes | Unclear | Yes | Yes | Yes |
| Yes | Unclear | Yes | Yes | Yes |
| Yes | Yes | Yes | Yes | Yes |
| No | Yes | Yes | Yes | Yes |
| No | Unclear | Yes | Yes | Yes |

| | | | | |
|---|---|---|---|---|
| Yes | Yes | Yes | Yes | Yes |
| Yes | Yes | Yes | Yes | Yes |
| Yes | Unclear | Yes | Yes | Yes |
| Yes | Unclear | Yes | Yes | Yes |
| Yes | Unclear | Yes | Yes | Yes |
| Unclear | Unclear | Yes | Yes | Yes |
| No | Unclear | Yes | Yes | Yes |
| No | Yes | Yes | Yes | Yes |
| Yes | Unclear | Yes | Unclear | Unclear |
| Yes | Unclear | Yes | Yes | Yes |
| Yes | Yes | Yes | Yes | Yes |
| Yes | Unclear | Yes | Yes | Yes |
| Yes | Yes | Yes | Yes | Yes |
| Yes | Unclear | Yes | Yes | Yes |
| Yes | Unclear | Yes | Yes | Yes |
| No | Yes | Yes | Yes | Yes |

| | | | | |
|---|---|---|---|---|
| Yes | Yes | Yes | Yes | Yes |
| Yes | Unclear | Yes | Yes | Yes |
| Yes | Yes | Yes | Yes | Yes |
| Yes | Unclear | Yes | Yes | Yes |
| No | Yes | Yes | Yes | Yes |
| No | Unclear | No, 2 follicles counted as TF | Yes | Yes |
| Yes | Unclear | Yes | Yes | Yes |
| Yes | Yes | Yes | Yes | Yes |
| Yes | Unclear | Yes | Yes | Yes |
| Yes | Unclear | Yes | Yes | Yes |
| Yes | Unclear | Yes | Yes | Yes |
| Yes | Yes | Yes | Yes | Yes |
| Yes | Yes | Yes | Yes | Yes |
| No | Yes | Yes | Yes | Yes |
| Yes | Yes | Yes | Yes | Yes |
| Yes | Unclear | Yes | Yes | Yes |
| No | Unclear | Unclear | Yes | Unclear |

| | | | | |
|---|---|---|---|---|
| Yes | Yes | Yes | Yes | Yes |
| Yes | Yes | Yes | Yes | Yes |
| No | Unclear | Yes | Yes | Yes |
| Yes | Unclear | Yes | Yes | Yes |
| Yes | Unclear | Yes | Yes | Unclear |
| No | Yes | Yes | Yes | Yes |
| Yes | Yes | Yes | Yes | Yes |
| No | Unclear | Yes | Yes | Yes |
| Yes | Unclear | Yes | Yes | Yes |
| Yes | Unclear | Yes | Yes | Yes |
| Yes | Unclear | Yes | Yes | Yes |
| Yes | Yes | Yes | Yes | Yes |
| Yes | Unclear | Yes | Yes | Yes |
| Yes | Unclear | Yes | Yes | Yes |
| Yes | Yes | Yes | Yes | Yes |
| Yes | Yes | Yes | Yes | Yes |
| Yes | Unclear | Yes | Yes | Yes |
| Yes | Yes | Yes | Yes | Yes |

| | | | | |
|---|---|---|---|---|
| Yes | Yes | Yes | Yes | Yes |
| No | Unclear | Unclear | Unclear | Unclear |
| Yes | Yes | Yes | Yes | Yes |
| Yes | Unclear | Yes | Yes | Yes |
| No | Unclear | Unclear | Yes | Yes |
| No | Yes | Yes | Yes | Yes |
| No | Unclear | Yes | Yes | Yes |
| No | Unclear | Yes | Yes | Yes |
| Yes | Yes | Yes | Yes | Yes |
| Yes | Yes | Yes | Yes | Yes |
| Yes | Yes | Yes | Yes | Yes |
| Yes | Yes | Yes | Yes | Yes |
| Yes | Yes | Yes | Yes | Yes |
| Yes | Yes | Yes | Yes | Yes |
| Yes | Unclear | Yes | Yes | Yes |
| Yes | Yes | Yes | Yes | Yes |
| Yes | Unclear | Yes | Yes | Yes |
| No | Yes | Yes | Yes | Unclear |
| Yes | Unclear | Yes | Yes | Yes |
| No | Unclear | Yes | Yes | Yes |

| | | | | |
|---|---|---|---|---|
| Yes | Low coverage of total pop (58%), coverage in 1-9 was higher (72%) | Yes | Yes | Yes |
| Yes | Low coverage (<80%) | Yes | Yes | Yes |
| No | Yes | Yes | Yes | Yes |
| No | Yes | Yes | Yes | Yes |
| Yes | Yes | Yes | Yes | Yes |
| No | Yes | Yes | Yes | Yes |
| Yes | Yes | Yes | Yes | Yes |
| Yes | Unclear | Yes | Yes | Yes |
| Yes | Yes | Yes | Yes | Yes |
| Yes | No | Yes | Yes | Yes |
| No | Yes | Yes | Yes | Yes |
| No | Unclear | Yes | Yes | Yes |
| No | No | Unclear | Yes | Yes |
| Yes | Unclear | Yes | Yes | Yes |
| Yes | Yes | Yes | Yes | Yes |
| Yes | Unclear | Yes | Yes | Yes |
| No | Yes | Yes | Yes | Yes |
| No | Unclear | Yes | Yes | Yes |

| | | | | |
|---|---|---|---|---|
| No | Unclear | Unclear | Yes | Unclear |
| No | Unclear | Yes | Yes | Yes |
| Yes | Unclear | Yes | Yes | Yes |
| Yes | Unclear | Yes | Yes | Yes |
| No | Unclear | Yes | Yes | Yes |
| No | Unclear | Yes | Yes | Yes |
| No | Yes | Unclear | Yes | Yes |
| Yes | Yes | Yes | Yes | Yes |
| No | Unclear | Yes | Yes | Yes |
| No | Yes | Yes | Yes | Yes |
| Yes | Yes | Yes | Yes | Yes |
| Yes | Unclear | Yes | Yes | Yes |
| Yes | Yes | Yes | Yes | Yes |
| Yes | Yes | Yes | Yes | Yes |
| Yes | Unclear | Yes | Yes | Yes |
| Yes | Unclear | Yes | Yes | Yes |
| Yes | Yes | Yes | Yes | Yes |

| | | | | |
|---|---|---|---|---|
| Yes | Yes | Yes | Yes | Yes |
| Yes | Yes | Yes | Yes | Yes |
| Yes | No, 78% coverage | Yes | Yes | Yes |
| No | Unclear | Yes | Yes | Yes |
| Yes | Unclear | Yes | Yes | Yes |
| No | Unclear | Yes | Yes | Yes |
| Yes | Yes | Yes | Yes | Yes |
| Yes | Yes | Yes | Yes | Yes |
| Yes | Unclear | Yes | Yes | Yes |
| Yes | Yes | Yes | Yes | Yes |
| Yes | Yes | Yes | Yes | Yes |
| No | Yes | Yes | Yes | Yes |
| Yes | Yes | Yes | Yes | Yes |
| No | Unclear | Unclear | Unclear | Unclear |
| Yes | Yes | Yes | Yes | Yes |
| Yes | Unclear | Yes | Yes | Yes |
| Yes | Yes | Yes | Yes | Yes |

| | | | | |
|---|---|---|---|---|
| Yes | No | Yes | Yes | Yes |
| Yes | Yes | Yes | Yes | Yes |
| Yes | Unclear | Yes | Yes | Yes |
| Yes | Unclear | Yes | Yes | Yes |
| No | Unclear | Yes | Yes | Yes |
| Yes | Yes | Yes | Yes | Yes |
| Yes | Unclear | Yes | Yes | Yes |
| No, No number sampled | Unclear | Yes | Yes | Yes |
| No | Unclear | Yes | Yes | Yes |
| Yes | Yes | Yes | Yes | Yes |
| Yes | Unclear | Unclear, TF definition Not given or cited | Yes | Yes |
| Yes | Unclear | Yes | Yes | Yes |
| Yes | Yes | Yes | Yes | Yes |
| No | Yes | Yes | Yes | Yes |
| No | Unclear | Yes | Yes | Yes |
| Yes | Unclear | Yes | Yes | Yes |
| Yes | Unclear | Yes | Yes | Yes |
| Yes | Unclear | Yes | Yes | Yes |

| | | | | |
|---|---|---|---|---|
| Yes | Unclear | Yes | Yes | Yes |
| Yes | Unclear | No, swabs included both upper tarsal and conj sac. In addition, cases were students with any number of follicles. | Yes | Yes |
| No, No year | Unclear | Yes | Yes | Yes |
| Yes | Unclear | Yes | Yes | Yes |
| Yes | Yes | Yes | Yes | Yes |
| No | Yes | Yes | Yes | Yes |
| No | Unclear | Yes | Yes | Yes |
| No | Unclear | Yes | Yes | Yes |
| No | Yes | Yes | Yes | Yes |
| No | Unclear | Yes | Yes | Yes |
| Yes | Unclear | Yes | Yes | Yes |
| Yes | Unclear | Yes | Yes | Yes |
| No, No year | Unclear | Yes | Yes | Yes |
| Yes | Unclear | Yes | Yes | Yes |
| Yes | Unclear | Yes | Yes | Yes |
| Yes | Yes | Yes | Yes | Yes |
| Yes | Unclear | Yes | Yes | Yes |

| | | | | |
|---|---|---|---|---|
| Yes | Yes | Yes | Yes | Yes |
| No | Unclear | Yes | Yes | Yes |
| Yes | Unclear | Yes | Yes | Yes |
| Yes | Yes | Yes | Yes | Yes |
| No | Unclear | Yes | Unclear | Unclear |
| Yes | Unclear | Yes | Yes | Yes |
| No | Unclear | Yes | Yes | Yes |
| No | Unclear | Yes | Yes | Yes |
| Yes | Unclear | Yes | Yes | Yes |
| Yes | Unclear | Yes | Yes | Yes |

| Risk of Bias checklist items** | | | | |
|---|---|---|---|---|
| **Prevalence study: Was the response rate adequate, and if not, was the low response rate managed appropriately?** | **DTA study: Was a consecutive or random sample of patients enrolled?** | **DTA study: Was a case-control design avoided?** | **DTA study: Did the study avoid inappropriate exclusions?** | **DTA: Were the index test results interpreted without knowledge of the results of the reference standard?** |
| Yes | | | | |
| Yes | | | | |
| Yes | | | | |
| Unclear | | | | |
| Yes | | | | |
| Yes | | | | |
| Yes | | | | |
| Yes | | | | |
| Yes | | | | |
| Yes | | | | |
| Unclear | | | | |
| Unclear | | | | |
| Unclear | | | | |
| Unclear | | | | |
| Unclear | | | | |

| | | | | |
|---|---|---|---|---|
| Yes | | | | |
| Yes | | | | |
| Unclear | | | | |
| Unclear | | | | |
| Yes | | | | |
| Yes | | | | |
| Yes | | | | |
| Unclear | | | | |
| Yes | | | | |
| Yes | | | | |
| Unclear | | | | |
| Unclear | Random | Yes | Yes | Yes |
| Unclear | | | | |
| Yes | | | | |
| Unclear | | | | |
| Unclear | Unclear | Unclear | Unclear | Unclear |
| Yes | Random | Yes | Yes | Yes |
| Unclear | Random | Yes | Yes | Yes |
| Yes | | | | |
| Yes | Unclear how village chosen | Yes | Yes | Unclear |
| Yes | | | | |

| | | | | |
|---|---|---|---|---|
| Unclear | | | | |
| Yes | Random | Yes | Yes | Yes |
| Yes | Random | Yes | Yes | Yes |
| Yes | | | | |
| Yes | | | | |
| Yes | | | | |
| Unclear | | | | |
| Yes | | | | |
| Unclear | | | | |
| Yes | | | | |
| Unclear | | | | |
| Unclear | | | | |
| Unclear | | | | |
| | Consecutive | Yes | No | Unclear |
| Yes | | | | |
| | Unclear | No | Yes | Unclear |
| | Unclear | No | Yes | Yes |
| | All children from selected villages | Yes | Yes | Yes |
| Yes | | | | |
| Unclear | | | | |

| | | | | |
|---|---|---|---|---|
| Yes | | | | |
| Yes | | | | |
| Unclear | | | | |
| Unclear | | | | |
| | Unclear | Yes | Yes | Unclear |
| | Unclear | Yes | Yes | Unclear |
| Unclear | | | | |
| Unclear | | | | |
| | Unclear | No | Yes | No |
| Unclear | | | | |
| Yes | | | | |
| Unclear | | | | |
| Unclear | | | | |
| Yes | | | | |
| | Unclear | No | Yes | Unclear |
| | Unclear | No | Yes | Unclear |
| Unclear | | | | |
| Yes | | | | |
| Unclear | | | | |
| Unclear | | | | |
| Yes | Unclear | Yes | Yes | Unclear |

| | | | | |
|---|---|---|---|---|
| Unclear | | | | |
| Unclear | | | | |
| Yes | | | | |
| Unclear | | | | |
| Yes | | | | |
| Unclear | | | | |
| Unclear | | | | |
| Yes | | | | |
| Unclear | | | | |
| Unclear | | | | |
| Unclear | Random | Yes | Yes | Unclear |
| Yes | | | | |
| Yes | | | | |
| Yes | | | | |
| | Unclear | Unclear | Unclear | Unclear |
| Yes | | | | |
| | Unclear | Yes | Yes | Unclear |
| Unclear | | | | |
| Unclear | | | | |

| | | | | |
|---|---|---|---|---|
| Yes | | | | |
| Yes | | | | |
| Unclear | | | | |
| Unclear | | | | |
| Unclear | | | | |
| Yes | | | | |
| Yes | | | | |
| Unclear | Random | Yes | Yes | Unclear |
| Unclear | | | | |
| Unclear | | | | |
| Unclear | | | | |
| Yes | | | | |
| Unclear | | | | |
| Unclear | random | Yes | Yes | Unclear |
| Yes | | | | |
| Yes | | | | |
| Unclear | | | | |
| Yes | | | | |

| | | | | |
|---|---|---|---|---|
| Yes | | | | |
| Unclear | | | | |
| Yes | | | | |
| | Unclear | Unclear | Unclear | Unclear |
| Unclear | | | | |
| Unclear | | | | |
| Yes | | | | |
| Unclear | | | | |
| Unclear | | | | |
| Yes | | | | |
| Yes | | | | |
| Yes | | | | |
| No | Unclear | Yes | Yes | Unclear |
| Yes | Random | Yes | Yes | Yes |
| Yes | | | | |
| Unclear | | | | |
| Yes | | | | |
| Unclear | | | | |
| Yes | | | | |
| Yes | | | | |
| Unclear | | | | |

| | | | | |
|---|---|---|---|---|
| No | | | | |
| Unclear | | | | |
| Yes | | | | |
| Yes | | | | |
| Yes | | | | |
| Yes | Unclear | Yes | Yes | Unclear |
| Yes | | | | |
| Unclear | Random | Yes | Yes | Yes |
| Yes | | | | |
| | Random | Yes | Yes | Yes |
| No | | | | |
| Yes | | | | |
| Unclear | | | | |
| Unclear | | | | |
| Unclear | | | | |
| Yes | | | | |
| Unclear | | | | |
| Yes | | | | |
| Unclear | Yes | Yes | Yes | Unclear |

| | | | | |
|---|---|---|---|---|
| Unclear | | | | |
| Unclear | | | | |
| Unclear | | | | |
| Unclear | | | | |
| Unclear | | | | |
| | Unclear | No | No, excluded on basis of ability to detect RNA | Unclear |
| Unclear | Random | Yes | Yes | Yes |
| Yes | | | | |
| Yes | | | | |
| Unclear | Random | Yes | Yes | Yes |
| Yes | | | | |
| Yes | | | | |
| Unclear | | | | |
| Yes | | | | |
| Yes | | | | |
| Unclear | | | | |
| Unclear | | | | |
| Yes | | | | |

| | | | | |
|---|---|---|---|---|
| Yes | | | | |
| Yes | | | | |
| Yes | Random | Yes | Yes | Yes |
| Unclear | Unclear | Yes | Yes | Yes |
| Unclear | | | | |
| Unclear | | | | |
| Yes | | | | |
| Yes | | | | |
| Unclear | | | | |
| Yes | | | | |
| Yes | | | | |
| Yes | | | | |
| Yes | | | | |
| Unclear | | | | |
| Yes | | | | |
| Unclear | | | | |
| Yes | | | | |

| | | | | |
|---|---|---|---|---|
| No | | | | |
| Yes | | | | |
| Unclear | | | | |
| Unclear | | | | |
| Unclear | Random | Yes | Yes | Yes |
| Yes | Random | Yes | Yes | Yes |
| Unclear | | | | |
| Unclear | | | | |
| Unclear | | | | |
| Yes | | | | |
| Unclear | | | | |
| Unclear | Unclear | Yes | Yes | Unclear |
| | Random | Yes | Yes | Unclear |
| Yes | NA, whole pop | Yes | Yes | Yes |
| Yes | | | | |
| Unclear | | | | |
| No | | | | |
| Unclear | | | | |
| Unclear | | | | |

| |
|---|
| Unclear |
| Unclear |
| Unclear |
| Unclear |
| Yes |
| Yes |
| Unclear |
| Unclear |
| Yes |
| Unclear |
| Unclear |
| Unclear |
| Unclear |
| Unclear |
| Unclear |
| Yes |
| Yes |

| | | | | |
|---|---|---|---|---|
| Yes | | | | |
| Unclear | Consecutive | Yes | Yes | Unclear |
| Unclear | Random | Yes | Yes | Unclear |
| Yes | | | | |
| Unclear | | | | |
| Unclear | | | | |
| Unclear | | | | |
| Unclear | | | | |
| Yes | Random | Yes | Yes | Yes |
| Yes | | | | |

| DTA: If a threshold was used, was it pre-specified? | DTA: Is the reference standard likely to correctly classify the target condition? | DTA: Were the reference standard results interpreted without knowledge of the results of the index test? | DTA: Was there an appropriate interval between index test and reference standard? | DTA: Did all patients receive the same reference standard? |
|---|---|---|---|---|

| | | | | |
|---|---|---|---|---|
| Yes | Unclear - LCA used | Yes | Yes | Yes |
| Unclear | Unclear | Unclear | Unclear | Unclear |
| Yes | Yes | Yes | Yes | Yes |
| Yes | Yes | Yes | Yes | Yes |
| Yes | Yes | Yes | Yes | Yes |

| | | | | |
|---|---|---|---|---|
| Yes | Yes | Yes | Yes | Yes |
| Yes | Yes | Yes | Yes | Yes |
| NA | Yes | Yes | Yes | Yes |
| Yes | Yes | Unclear | Yes | Yes |
| Yes | Yes | Unclear | Yes | Yes |
| Yes | Yes | Unclear | Yes | Yes |

| | | | | |
|---|---|---|---|---|
| Yes | Yes | Yes | Yes | Yes |
| Yes | Yes | Yes | Yes | Yes |
| Yes | Yes | No | Yes | Yes |
| Yes | Yes | Yes | Unclear | Yes |
| Yes | Yes | Yes | Unclear | Yes |
| Yes | Yes | Yes | Yes | Yes |

| | | | | |
|---|---|---|---|---|
| Yes | Yes | Yes | Yes | Yes |
| Yes | Yes | Unclear | Yes | Yes |
| Yes | Yes | Yes | Yes | Yes |

| | | | | |
|---|---|---|---|---|
| Yes | Yes | Yes | Yes | Yes |
| Yes | Yes | Yes | Yes | Yes |

| | | | | |
|---|---|---|---|---|
| Unclear | Unclear | Yes | Unclear | Unclear |
| Yes | Yes | Yes | Yes | Yes |
| Yes | Yes | Yes | Yes | Yes |

| | | | | |
|---|---|---|---|---|
| Yes | Yes | Yes | Yes | Yes |
| Yes | Yes | Yes | Yes | Yes |
| Yes | Yes | Yes | Yes | Yes |
| Yes | Yes | Yes | Yes | Yes |

| | | | | |
|---|---|---|---|---|
| Yes | Yes | Yes | Yes | Yes |
| Yes | Yes | Yes | Yes | Yes |
| Yes | Yes | Yes | Yes | Yes |

| | | | | |
|---|---|---|---|---|
| Yes | Yes | Yes | Yes | Yes |
| Yes | Yes | Yes | Yes | Yes |

| | | | | |
|---|---|---|---|---|
| Yes | Yes | Yes | Yes | Yes |
| Yes | Yes | Yes | Yes | Yes |
| Yes | Yes | Yes | Yes | Yes |
| Yes | Yes | Yes | Yes | Yes |
| Yes | Yes | Yes | Yes | Yes |

| | | | | |
|---|---|---|---|---|
| Yes | Yes | Yes | Yes | Yes |
| Yes | Yes | Yes | Yes | Yes |
| Yes | Yes | Yes | Yes | Yes |

| DTA: Were all patients included in the analysis? | Prevalence study score [meets criteria/total] (%) | DTA study score [meets criteria/total] (%) | Max score (if both checklists used, higher of 2 scores) |
|---|---|---|---|
| | [9/9] (100%) | NA | 100% |
| | [8/9] (89%) | NA | 89% |
| | [9/9] (100%) | NA | 100% |
| | [7/9] (78%) | NA | 78% |
| | [8/9] (89%) | NA | 89% |
| | [8/9] (89%) | NA | 89% |
| | [9/9] (100%) | NA | 100% |
| | [8/9] (89%) | NA | 89% |
| | [9/9] (100%) | NA | 100% |
| | [9/9] (100%) | NA | 100% |
| | [5/9] (56%) | NA | 56% |
| | [6/9] (67%) | NA | 67% |
| | [6/9] (67%) | NA | 67% |
| | [6/9] (67%) | NA | 67% |
| | [5/9] (56%) | NA | 56% |

| | | | |
|---|---|---|---|
| | [9/9] (100%) | NA | 100% |
| | [8/9] (89%) | NA | 89% |
| | [5/9] (56%) | NA | 56% |
| | [4/9] (44%) | NA | 44% |
| | [6/9] (67%) | NA | 67% |
| | [8/9] (89%) | NA | 89% |
| | [9/9] (100%) | NA | 100% |
| | [7/9] (78%) | NA | 78% |
| | [9/9] (100%) | NA | 100% |
| | [9/9] (100%) | NA | 100% |
| | [5/9] (56%) | NA | 56% |
| Yes | [7/9] (78%) | [8/10] (80%) | 80% |
| | [0/9] (0%) | NA | 0% |
| | [8/9] (89%) | NA | 89% |
| | [5/9] (56%) | NA | 56% |
| Unclear | [0/9] (0%) | [0/10] (0%) | 0% |
| Yes | [9/9] (100%) | [9/10] (90%) | 100% |
| Yes | [6/9] (67%) | [9/10] (90%) | 90% |
| | [7/9] (78%) | NA | 78% |
| Yes | [7/9] (78%) | [8/10] (80%) | 80% |
| | [9/9] (100%) | NA | 100% |

| | | | |
|---|---|---|---|
| | [7/9] (78%) | NA | 78% |
| Yes | [8/9] (89%) | [9/10] (90%) | 90% |
| Yes | [9/9] (100%) | [9/10] (90%) | 100% |
| | [8/9] (89%) | NA | 89% |
| | [8/9] (89%) | NA | 89% |
| | [8/9] (89%) | NA | 89% |
| | [4/9] (44%) | NA | 44% |
| | [9/9] (100%) | NA | 100% |
| | [1/9] (11%) | NA | 11% |
| | [8/9] (89%) | NA | 89% |
| | [5/9] (56%) | NA | 56% |
| | [6/9] (67%) | NA | 67% |
| | [7/9] (78%) | NA | 78% |
| No | NA | [5/10] (50%) | 50% |
| | [9/9] (100%) | NA | 100% |
| Yes | NA | [6/10] (60%) | 60% |
| Yes | NA | [7/10] (70%) | 70% |
| Yes | NA | [8/10] (80%) | 80% |
| | [7/9] (78%) | NA | 78% |
| | [4/9] (44%) | NA | 44% |

| | | | |
|---|---|---|---|
| | [8/9] (89%) | NA | 89% |
| | [8/9] (89%) | NA | 89% |
| | [6/9] (67%) | NA | 67% |
| | [6/9] (67%) | NA | 67% |
| Yes | NA | [8/10] (80%) | 80% |
| Unclear | NA | [7/10] (70%) | 70% |
| | [6/9] (67%) | NA | 67% |
| | [6/9] (67%) | NA | 67% |
| Yes | NA | [6/10] (60%) | 60% |
| | [4/9] (44%) | NA | 44% |
| | [8/9] (89%) | NA | 89% |
| | [3/9] (33%) | NA | 33% |
| | [6/9] (67%) | NA | 67% |
| | [9/9] (100%) | NA | 100% |
| Yes | NA | [6/10] (60%) | 60% |
| Yes | NA | [6/10] (60%) | 60% |
| | [5/9] (56%) | NA | 56% |
| | [9/9] (100%) | NA | 100% |
| | [7/9] (78%) | NA | 78% |
| | [7/9] (78%) | NA | 78% |
| Yes | [8/9] (89%) | [8/10] (80%) | 89% |

| | | | |
|---|---|---|---|
| | [8/9] (89%) | NA | 89% |
| | [6/9] (67%) | NA | 67% |
| | [8/9] (89%) | NA | 89% |
| | [6/9] (67%) | NA | 67% |
| | [8/9] (89%) | NA | 89% |
| | [4/9] (44%) | NA | 44% |
| | [7/9] (78%) | NA | 78% |
| | [9/9] (100%) | NA | 100% |
| | [7/9] (78%) | NA | 78% |
| | [7/9] (78%) | NA | 78% |
| Yes | [6/9] (67%) | [8/10] (80%) | 80% |
| | [9/9] (100%) | NA | 100% |
| | [9/9] (100%) | NA | 100% |
| | [8/9] (89%) | NA | 89% |
| Unclear | NA | [4/10] (40%) | 40% |
| | [9/9] (100%) | NA | 100% |
| Yes | NA | [8/10] (80%) | 80% |
| | [7/9] (78%) | NA | 78% |
| | [3/9] (33%) | NA | 33% |

| | | | |
|---|---|---|---|
| | [9/9] (100%) | NA | 100% |
| | [9/9] (100%) | NA | 100% |
| | [6/9] (67%) | NA | 67% |
| | [7/9] (78%) | NA | 78% |
| | [4/9] (44%) | NA | 44% |
| | [7/9] (78%) | NA | 78% |
| | [9/9] (100%) | NA | 100% |
| Yes | [6/9] (67%) | [8/10] (80%) | 80% |
| | [7/9] (78%) | NA | 78% |
| | [6/9] (67%) | NA | 67% |
| | [7/9] (78%) | NA | 78% |
| | [6/9] (67%) | NA | 67% |
| | [6/9] (67%) | NA | 67% |
| Yes | [6/9] (67%) | [8/10] (80%) | 80% |
| | [9/9] (100%) | NA | 100% |
| | [9/9] (100%) | NA | 100% |
| | [6/9] (67%) | NA | 67% |
| | [9/9] (100%) | NA | 100% |

| | | | |
|---|---|---|---|
| | [9/9] (100%) | NA | 100% |
| | [0/9] (0%) | NA | 0% |
| | [9/9] (100%) | NA | 100% |
| Unclear | NA | [1/10] (10%) | 10% |
| | [7/9] (78%) | NA | 78% |
| | [2/9] (22%) | NA | 22% |
| | [8/9] (89%) | NA | 89% |
| | [3/9] (33%) | NA | 33% |
| | [5/9] (56%) | NA | 56% |
| | [8/9] (89%) | NA | 89% |
| | [9/9] (100%) | NA | 100% |
| | [8/9] (89%) | NA | 89% |
| Yes | [7/9] (78%) | [8/10] (80%) | 80% |
| Yes | [9/9] (100%) | [9/10] (90%) | 100% |
| | [9/9] (100%) | NA | 100% |
| | [4/9] (44%) | NA | 44% |
| | [9/9] (100%) | NA | 100% |
| | [7/9] (78%) | NA | 78% |
| | [4/9] (44%) | NA | 44% |
| | [8/9] (89%) | NA | 89% |
| | [3/9] (33%) | NA | 33% |

| | | | |
|---|---|---|---|
| | [7/9] (78%) | NA | 78% |
| | [5/9] (56%) | NA | 56% |
| | [8/9] (89%) | NA | 89% |
| | [8/9] (89%) | NA | 89% |
| | [8/9] (89%) | NA | 89% |
| Yes | [6/9] (67%) | [8/10] (80%) | 80% |
| | [9/9] (100%) | NA | 100% |
| Yes | [6/9] (67%) | [9/10] (90%) | 90% |
| | [8/9] (89%) | NA | 89% |
| Yes | NA | [9/10] (90%) | 90% |
| | [6/9] (67%) | NA | 67% |
| | [8/9] (89%) | NA | 89% |
| | [6/9] (67%) | NA | 67% |
| | [4/9] (44%) | NA | 44% |
| | [6/9] (67%) | NA | 67% |
| | [9/9] (100%) | NA | 100% |
| | [7/9] (78%) | NA | 78% |
| | [8/9] (89%) | NA | 89% |
| Yes | [6/9] (67%) | [9/10] (90%) | 90% |

| | | | |
|---|---|---|---|
| | [2/9] (22%) | NA | 22% |
| | [5/9] (56%) | NA | 56% |
| | [6/9] (67%) | NA | 67% |
| | [6/9] (67%) | NA | 67% |
| | [5/9] (56%) | NA | 56% |
| No, omitted people without adequate quantity of RNA | NA | [5/10] (50%) | 50% |
| Yes, although ungradeable images excluded for some analyses | [3/9] (33%) | [8/10] (80%) | 80% |
| | [5/9] (56%) | NA | 56% |
| | [9/9] (100%) | NA | 100% |
| Yes | [5/9] (56%) | [9/10] (90%) | 90% |
| | [7/9] (78%) | NA | 78% |
| | [9/9] (100%) | NA | 100% |
| | [6/9] (67%) | NA | 67% |
| | [9/9] (100%) | NA | 100% |
| | [9/9] (100%) | NA | 100% |
| | [6/9] (67%) | NA | 67% |
| | [6/9] (67%) | NA | 67% |
| | [9/9] (100%) | NA | 100% |

| | | | |
|---|---|---|---|
| | [8/9] (89%) | NA | 89% |
| | [8/9] (89%) | NA | 89% |
| Depends on measure | [8/9] (89%) | [8/10] (80%) | 89% |
| Yes | [3/9] (33%) | [9/10] (90%) | 90% |
| | [6/9] (67%) | NA | 67% |
| | [5/9] (56%) | NA | 56% |
| | [9/9] (100%) | NA | 100% |
| | [8/9] (89%) | NA | 89% |
| | [5/9] (56%) | NA | 56% |
| | [9/9] (100%) | NA | 100% |
| | [9/9] (100%) | NA | 100% |
| | [6/9] (67%) | NA | 67% |
| | [8/9] (89%) | NA | 89% |
| | [1/9] (11%) | NA | 11% |
| | [9/9] (100%) | NA | 100% |
| | [7/9] (78%) | NA | 78% |
| | [9/9] (100%) | NA | 100% |

| | | | |
|---|---|---|---|
| | [7/9] (78%) | NA | 78% |
| | [9/9] (100%) | NA | 100% |
| | [6/9] (67%) | NA | 67% |
| | [6/9] (67%) | NA | 67% |
| Yes | [5/9] (56%) | [9/10] (90%) | 90% |
| Yes | [8/9] (89%) | [9/10] (90%) | 90% |
| | [7/9] (78%) | NA | 78% |
| | [5/9] (56%) | NA | 56% |
| | [5/9] (56%) | NA | 56% |
| | [9/9] (100%) | NA | 100% |
| | [4/9] (44%) | NA | 44% |
| Yes | [4/9] (44%) | [8/10] (80%) | 80% |
| Yes | NA | [8/10] (80%) | 80% |
| Yes | [9/9] (100%) | [9/10] (90%) | 100% |
| | [8/9] (89%) | NA | 89% |
| | [5/9] (56%) | NA | 56% |
| | [6/9] (67%) | NA | 67% |
| | [6/9] (67%) | NA | 67% |
| | [7/9] (78%) | NA | 78% |

| | | | |
|---|---|---|---|
| | [5/9] (56%) | NA | 56% |
| | [3/9] (33%) | NA | 33% |
| | [4/9] (44%) | NA | 44% |
| | [4/9] (44%) | NA | 44% |
| | [9/9] (100%) | NA | 100% |
| | [8/9] (89%) | NA | 89% |
| | [5/9] (56%) | NA | 56% |
| | [5/9] (56%) | NA | 56% |
| | [8/9] (89%) | NA | 89% |
| | [5/9] (56%) | NA | 56% |
| | [7/9] (78%) | NA | 78% |
| | [7/9] (78%) | NA | 78% |
| | [6/9] (67%) | NA | 67% |
| | [7/9] (78%) | NA | 78% |
| | [7/9] (78%) | NA | 78% |
| | [9/9] (100%) | NA | 100% |
| | [6/9] (67%) | NA | 67% |

| | | | |
|---|---|---|---|
| | [7/9] (78%) | NA | 78% |
| Yes | [4/9] (44%) | [8/10] (80%) | 80% |
| Yes | [6/9] (67%) | [8/10] (80%) | 80% |
| | [9/9] (100%) | NA | 100% |
| | [1/9] (11%) | NA | 11% |
| | [7/9] (78%) | NA | 78% |
| | [6/9] (67%) | NA | 67% |
| | [5/9] (56%) | NA | 56% |
| Yes | [8/9] (89%) | [9/10] (90%) | 90% |
| | [8/9] (89%) | NA | 89% |
| | | | 0% |
| | | | 0% |
| | | | 0% |
| | | | 0% |
| | | | 0% |
| | | | 0% |
| | | | 0% |
| | | | 0% |

| reference number | Title | Authors |
|---|---|---|
| 1 | High prevalence of clinically active trachoma and its associated | Abdilwohab, M. G.; Abebo, Z. H. |
| 2 | Prevalence and risk factors for trachoma and ocular Chlamydia | Abdou, A.; Nassirou, B.; Kadri, B.; Mou |
| 3 | Prevalence and distribution of active trachoma among children | Abebo, T. A.; Tesfaye, D. J. |
| 4 | Prevalence of active trachoma among children between 1-9 ye | Yilikal, Adamu; Semira, Fereji |
| 5 | Altitude - a risk factor for active trachoma in southern Ethiopia | Tesfay, Haileselassie; Samson, Bayu |
| 6 | The burden of trachoma in the rural Nile Delta of Egypt: a surv | Al-Arab, G. E.; Tawfik, N.; El-Gendy, R. |
| 7 | Active trachoma in children in central Ethiopia: association wit | Wondu, Alemayehu; Muluken, Melese |
| 8 | Prevalence of trachoma in four marakez of Elmenia and Bani S | Amer, K.; Muller, A.; Abdelhafiz, H. M. |
| 9 | Elimination of active trachoma after two topical mass treatme | Amza, A.; Goldschmidt, P.; Einterz, E.; |
| 10 | Community risk factors for ocular chlamydia infection in Niger: | Amza, A.; Kadri, B.; Nassirou, B.; Stolle |
| 11 | The Easiest Children to Reach Are Most Likely to Be Infected w | Amza, A.; Kadri, B.; Nassirou, B.; Yu, S. |
| 12 | A cluster-randomized trial to assess the efficacy of targeting tr | Amza, A.; Kadri, B.; Nassirou, B.; Cotte |
| 13 | Effectiveness of expanding annual mass azithromycin distributi | Amza, A.; Kadri, B.; Nassirou, B.; Cotte |
| 14 | Community-level association between clinical trachoma and o | Amza, A.; Kadri, B.; Nassirou, B.; Cotte |
| 15 | Trachoma control in Southern Zambia - an international team | Astle, W. F.; Wiafe, B.; Ingram, A. D.; M |
| 16 | Risk factors for ocular chlamydia after three mass azithromycin | Ayele, B.; Gebre, T.; Moncada, J.; Hous |
| 17 | Low prevalence of active trachoma and associated factors amo | Ayelgn, K.; Guadu, T.; Getachew, A. |
| 18 | [What do we know about trachoma in the economically weake | Ayena, K. D.; Amza, A.; Agbo, Y. M.; Do |
| 19 | [Trachoma rapid assessment in the infantile population of Tog | Ayena, K. D.; Dzidzinyo, K.; Koffi, K. S.; |
| 20 | The problem of trachoma within an onchocerciasis-endemic zo | Babalola, O. E.; Abiose, A.; Barrie, J. R. |
| 21 | Polymerase chain reaction for the detection of ocular chlamyd | Bailey, R. L.; Hampton, T. J.; Hayes, L. J |
| 22 | Evaluation of the prevalence of trachoma 12 years after baseli | Bamani, S.; Dembele, M.; Sankara, D.; |
| 23 | Where do we go from here? Prevalence of trachoma three yea | Bamani, S.; King, J. D.; Dembele, M.; C |
| 24 | Prevalence of trachoma and its determinants in Dalocha Distri | Bejiga, A.; Alemayehu, W. |
| 25 | Effectiveness and safety of azithromycin 1.5% eye drops for ma | Bella, A. L.; Einterz, E.; Huguet, P.; Ben |
| 26 | Prevalence of trachoma in Ethiopia | Yemane, Berhane; Alemayehu, Worku |
| 27 | Application of smartphone cameras for detecting clinically acti | Bhosai, S. J.; Amza, A.; Beido, N.; Baile |
| 28 | Piloting serological tools for trachoma postelimination surveilla | Bid, R.; Sandi, F.; Goodhew, B.; Massa |
| 29 | Prevalence of trachoma in northern Benin: results from 11 pop | Bio, A. A.; Boko, P. M.; Dossou, Y. A.; T |
| 30 | Diagnosis of Chlamydia trachomatis eye infection in Tanzania b | Bobo, L.; Munoz, B.; Viscidi, R.; Quinn, |
| 31 | Crowdsourcing can match field grading validity for follicular tra | Brady, C. J.; Wolle, M. A.; Naufal, F.; M |
| 32 | Association between ocular bacterial carriage and follicular tra | Burr, S. E.; Hart, J. D.; Edwards, T.; Bal |
| 33 | Pgp3 seroprevalence and associations with active trachoma an | Burr, S. E.; Hart, J.; Samikwa, L.; Chaim |
| 34 | Profound and sustained reduction in Chlamydia trachomatis in | Burton, M. J.; Holland, M. J.; Makalo, P |
| 35 | Active trachoma is associated with increased conjunctival expr | Burton, M. J.; Ramadhani, A.; Weiss, H |
| 36 | Low prevalence of conjunctival infection with Chlamydia trach | Butcher, R. M. R.; Sokana, O.; Jack, K.; |
| 37 | Using alternate indicators to define need for public health inte | Butcher, R. |
| 38 | Clinical signs of trachoma are prevalent among Solomon Island | Butcher, R.; Sokana, O.; Jack, K.; Sui, L |
| 39 | Ocular Chlamydia trachomatis infection, anti-Pgp3 antibodies a | Butcher, R.; Handley, B.; Garae, M.; Ta |
| 40 | Trachoma prevalence and risk factors among preschool childre | Abreu Caligaris, L. S.; Miyake Morimot |
| 41 | Prevalence of signs of trachoma, ocular Chlamydia trachomatis | Cama, A.; Muller, A.; Taoaba, R.; Butch |
| 42 | Risk of seroconversion and seroreversion of antibodies to Chla | Chen, X.; Munoz, B.; Mkocha, H.; Gayd |
| 43 | Efficacy and safety of short duration azithromycin eye drops ve | Cochereau, I.; Goldschmidt, P.; Goepo |
| 44 | Community seroprevalence survey for yaws and trachoma in t | Cocks, N.; Rainima-Qaniuci, M.; Yalen, |
| 45 | Low prevalence of ocular Chlamydia trachomatis infection and | Macleod, C. K.; Butcher, R.; Mudaliar, |

| | | |
|---|---|---|
| 46 | Prevalence of trachomatous scarring in children in a formerly e | Cox, J. T.; Mkocha, H.; Munoz, B.; Wes |
| 47 | Trachoma prevalence in Niger: results of 31 district-level surve | Cromwell, E. A.; Amza, A.; Kadri, B.; Be |
| 48 | Active trachoma in children aged three to nine years in rural co | Cumberland, P.; Girum, Hailu; Todd, J. |
| 49 | The impact of community level treatment and preventative int | Cumberland, P.; Edwards, T.; Hailu, G. |
| 50 | Trachoma: epidemiologic study of scholars from Alagoas state | Damasceno, R. W. F.; Santos, R. R.; Ca |
| 51 | Ophthalmia neonatorum in a trachoma endemic area | Datta, P.; Frost, E.; Peeling, R.; Masind |
| 52 | Elimination of trachoma as a public health problem in Ghana: p | Debrah, O.; Mensah, E. O.; Senyonjo, L |
| 53 | Can corneal pannus with trachomatous inflammation - follicula | Derrick, T.; Holland, M. J.; Cassama, E. |
| 54 | Inverse relationship between microRNA-155 and -184 expressi | Derrick, T.; Last, A. R.; Burr, S. E.; Robe |
| 55 | DjinniChip: evaluation of a novel molecular rapid diagnostic de | Derrick, T. R.; Sandetskaya, N.; Pickeri |
| 56 | Sequelae from Epidemic Viral Conjunctivitis Can Be Associated | de Sousa Meneghim, R. L. F.; Viveiros, |
| 57 | Nationwide integrated mapping of three neglected tropical dis | Dorkenoo, A. M.; Bronzan, R. N.; Ayen |
| 58 | Epidemiology of trachoma and its implications for implementin | Ahmed Badei, Duale; Nebiyu Negussu, |
| 59 | Prevalence of trachoma in Unity State, South Sudan: results fro | Edwards, T.; Smith, J.; Sturrock, H. J. W |
| 60 | Rapid trachoma assessment in Kersa District, Southwest Ethiop | Ejigu, M.; Kariuki, M. M.; Ilako, D. R.; G |
| 61 | Surveillance and azithromycin treatment for newcomers and tr | Ervin, A. M.; Mkocha, H.; Munoz, B.; D |
| 62 | Temporal cytokine gene expression patterns in subjects with tr | Faal, N.; Bailey, R. L.; Sarr, I.; Joof, H.; I |
| 63 | Conjunctival FOXP3 expression in trachoma: do regulatory T ce | Faal, N.; Bailey, R. L.; Jeffries, D.; Joof, |
| 64 | Prevalence of trachoma in school children in the Marajo Archip | Favacho, J.; Alves da Cunha, A. J. L.; Go |
| 65 | Rapid assessment for prioritisation of trachoma control at com | Faye, M.; Kuper, H.; Dineen, B.; Bailey, |
| 66 | Studies of human immune responses to various antigenic prote | Felton, JM |
| 67 | Survey, culture, and genome analysis of ocular Chlamydia trach | Feng, L.; Lu, X.; Yu, Y.; Wang, T.; Luo, S |
| 68 | Prevalence and determinants of active trachoma among prescl | Ferede, A. T.; Dadi, A. F.; Tariku, A.; Ad |
| 69 | Trachoma in Indigenous Settlements in Brazil, 2000-2008 | Freitas, H. S. A.; Medina, N. H.; Lopes, |
| 70 | School-based intervention: evaluating the role of water, latrine | Gelaye, B.; Kumie, A.; Aboset, N.; Berh |
| 71 | Prevalence of active trachoma and its associated factors amon | Genet, A.; Dagnew, Z.; Melkie, G.; Kele |
| 72 | Detection of Chlamydiaceae and Chlamydia-like organisms on t | Ghasemian, E.; Inic-Kanada, A.; Colling |
| 73 | Comparison of genovars and Chlamydia trachomatis infection l | Ghasemian, E.; Inic-Kanada, A.; Colling |
| 74 | Clinical and microbiological diagnosis of trachoma in children li | Goldschmidt, P.; Afghani, T.; Nadeem, |
| 75 | Clinical and microbiological assessment of trachoma in the Kol | Goldschmidt, P.; Benallaoua, D.; Amza |
| 76 | The limits of medical interventions for the elimination of preve | Goldschmidt, P.; Einterz, E. |
| 77 | Access to water source, latrine facilities and other risk factors o | Golovaty, I.; Jones, L.; Gelaye, B.; Melk |
| 78 | Chlamydial positivity of nasal discharge at baseline is associate | Gower, E. W.; Solomon, A. W.; Burton |
| 79 | Prevalence of Chlamydia trachomatis-Specific Antibodies befor | Gwyn, S. E.; Xiang, L.; Kandel, R. P.; De |
| 80 | Optimization of a rapid test for antibodies to the Chlamydia tra | Gwyn, S.; Mkocha, H.; Randall, J. M.; K |
| 81 | Comparison of platforms for testing antibodies to Chlamydia tr | Gwyn, S.; Awoussi, M. S.; Bakhtiari, A. |
| 82 | The performance of immunoassays to measure antibodies to t | Gwyn, S.; Nute, A. W.; Eshetu, Sata; Ze |
| 83 | Population-based prevalence of Chlamydia trachomatis infecti | Nash, S. D.; Astale, T.; Nute, A. W.; Bet |
| 84 | Trachoma in northern Ghana: a need for further studies | Gyasi, M. E.; Nsiire, A.; Yayemain, D.; D |
| 85 | Prevalence of active trachoma two years after control activitie | Hagan, M.; Yayemain, D.; Ahorsu, F.; A |
| 86 | Prevalence of and risk factors for trachoma in Southern Nation | Tesfaye Haileselassie, Adera; Macleod |
| 87 | Post-Validation Survey in Two Districts of Morocco after the El | Hammou, J.; Guagliardo, S. A. J.; Obtel |
| 88 | Risk factors for active trachoma in The Gambia | Harding-Esch, E. M.; Edwards, T.; Sillah |
| 89 | Active trachoma and ocular Chlamydia trachomatis infection in | Harding-Esch, E. M.; Edwards, T.; Sillah |
| 90 | Trachoma prevalence and associated risk factors in The Gambi | Harding-Esch, E. M.; Edwards, T.; Mko |
| 91 | Mass treatment with azithromycin for trachoma: when is one y | Harding-Esch, E. M.; Sillah, A.; Edward |
| 92 | Population-based prevalence survey of follicular trachoma and | Harding-Esch, E. M.; Kadimpeul, J.; Sar |

| | | |
|---|---|---|
| 93 | Impact of a single round of mass drug administration with azith | Harding-Esch, E. M.; Holland, M. J.; Sch |
| 94 | Inapparent ocular infection by Chlamydia trachomatis in exper | Hudson, A. P.; McEntee, C. M.; Reache |
| 95 | Mass treatment of trachoma with azithromycin 1.5% eye drops | Huguet, P.; Bella, L.; Einterz, E. M.; Go |
| 96 | Field evaluation of the cepheid GeneXpert Chlamydia trachoma | Jenson, A.; Dize, L.; Mkocha, H.; Munc |
| 97 | Blinding trachoma in Katsina State, Nigeria: population-based p | Jip, N. F.; King, J. D.; Diallo, M. O.; Miri |
| 98 | Evaluation of four years implementation of the safe strategy (s | Kabona, G.; Ngondi, J.; Kirumbi, E.; Kar |
| 99 | Prevalence and risk factors for trachoma in central and southe | Kalua, K.; Chirwa, T.; Kalilani, L.; Abber |
| 100 | Scaling up of trachoma mapping in Salima District, Central Mal | Kalua, K.; Singini, I.; Mukaka, M.; Msya |
| 101 | Incidence and progression of trachomatous scarring in a cohor | Kashaf, M. S.; Munoz, B. E.; Mkocha, H |
| 102 | Prevalence of active trachoma and associated risk factors amo | Kassim, K.; Kassim, J.; Aman, R.; Abdul |
| 103 | Alternative indicators to monitor trachoma elimination; does l | Kasubi, M.; Kabona, G.; Simon, A.; Kala |
| 104 | Prevalence and risk factors for trachoma in Sarlahi district, Nep | Katz, J.; West Jr, K. P.; Khatry, S. K.; Le |
| 105 | Prevalence and factors associated with active trachoma among | Kedir, S.; Lemnuro, K.; Yesse, M.; Abde |
| 106 | Diagnostic characteristics of tests for ocular Chlamydia after m | Keenan, J. D.; See, C. W.; Moncada, J.; |
| 107 | Mass azithromycin distribution for hyperendemic trachoma fol | Keenan, J. D.; Tadesse, Z.; Gebresillasi |
| 108 | Active trachoma and associated risk factors among children in | Ketema, K.; Tiruneh, M.; Woldeyohan |
| 109 | Outcome of azithromycin treatment of active trachoma in Oma | Khandekar, R.; Mohammed, A. J. |
| 110 | Active trachoma, face washing (F) and environmental improve | Khandekar, R.; Mabry, R.; Al-Hadrami, |
| 111 | Rapid assessment of trachoma among children living in rural N | Sumeet, Khanduja; Vishal, Jhanji; Nam |
| 112 | Community-level chlamydial serology for assessing trachoma e | Kim, J. S.; Oldenburg, C. E.; Cooley, G.; |
| 113 | The burden of trachoma in Ayod County of Southern Sudan | King, J. D.; Ngondi, J.; Gatpan, G.; Lopi |
| 114 | Mapping trachoma in Nasarawa and Plateau States, central Nig | King, J. D.; Jip, N.; Jugu, Y. S.; Othman, |
| 115 | Trachoma among children in community surveys from four Afr | King, J. D.; Odermatt, P.; Utzinger, J.; N |
| 116 | Prevalence of trachoma at sub-district level in Ethiopia: determ | King, J. D.; Teferi, T.; Cromwell, E. A.; Z |
| 117 | The epidemiology of trachoma in the five northern districts of | Koroma, J. B.; Heck, E.; Vandy, M.; Sor |
| 118 | Evaluation of diagnostics of chlamydia trachomatis infection in | Koukounari, A.; Moustaki, I.; Blake, I. M |
| 119 | Trachoma in Western Equatoria State, Southern Sudan: implica | Kur, L. W.; Picon, D.; Adibo, O.; Robins |
| 120 | Detection of chlamydial nucleic acid in follicular trachoma | Laming, A. C.; Hallsworth, P. J. |
| 121 | Risk factors for active trachoma and ocular Chlamydia trachom | Last, A. R.; Burr, S. E.; Weiss, H. A.; Ha |
| 122 | Development and evaluation of a next-generation digital PCR d | Roberts, C. H.; Last, A.; Molina-Gonzal |
| 123 | Trachoma elimination in an endemic island setting in West Afri | Last, A.; Cassama, E.; Bojang, E.; Nabic |
| 124 | The impact of a single round of community mass treatment wi | Last, A. R.; Burr, S. E.; Harding-Esch, E. |
| 125 | Decline of ocular Chlamydia trachomatis infection and follicula | Lee, J.; Munoz, B.; Mkocha, H.; Gaydos |
| 126 | The effect of multiple rounds of mass drug administration on t | Lee, J. S.; Munoz, B. E.; Mkocha, H.; Ga |
| 127 | Study of infectious conjunctivitis among children in rural areas | Liang, QingFeng; Lu, XinXin; Wang, Me |
| 128 | Short-term forecasting of the prevalence of clinical trachoma: | Liu, F. C.; Porco, T. C.; Amza, A.; Kadri, |
| 129 | Prevalence of trachoma in schoolchildren in Brazil | Luna, E. J. de A.; Lopes, M. de F. C.; M |
| 130 | Clinical signs of trachoma and laboratory evidence of ocular Ch | Lynch, K. D.; Morotti, W.; Brian, G.; Ke |
| 131 | A national survey integrating clinical, laboratory, and WASH da | Lynch, K. D.; Apadinuwe, S. C.; Lamber |
| 132 | Discord between presence of follicular conjunctivitis and Chlan | Lynch, K. D.; Brian, G.; Ahwang, T.; Ne |
| 133 | Expression of MHC class II antigens by conjunctival epithelial c | Mabey, D. C.; Bailey, R. L.; Dunn, D.; Jo |
| 134 | Unimproved water sources and open defecation are associated | Macleod, C. K.; Binnawi, K. H.; Elshafie |
| 135 | Trachoma, anti-Pgp3 serology, and ocular Chlamydia trachoma | Macleod, C. K.; Butcher, R.; Javati, S.; G |
| 136 | Trachoma in Northern Chad: Epidemiological survey. [French] | Madani, M. O.; Huguet, P.; Mariotti, S |
| 137 | Prevalence of trachoma in Car-Nicobar Island, India after three | Sumit, Malhotra; Praveen, Vashist; No |
| 138 | Serological Measures of Trachoma Transmission Intensity | Martin, D. L.; Wiegand, R.; Goodhew, |
| 139 | Serology for trachoma surveillance after cessation of mass dru | Martin, D. L.; Bid, R.; Sandi, F.; Goodhe |

| | | |
|---|---|---|
| 140 | Trachoma in the Pacific Islands: evidence from Trachoma Rapid | Mathew, A. A.; Keeffe, J. E.; Mesurier, |
| 141 | Epidemiological surveillance of trachoma in a school in the city | Medina, N. H.; Massaini, M. G.; Azeve |
| 142 | Assessment of trachoma in Cambodia: trachoma is not a public | Meng, N.; Seiha, D.; Thorn, P.; Willis, F |
| 143 | A community-based trachoma survey: prevalence and risk fact | Mesfin, M. M.; Camera, J. de la; Tarek |
| 144 | Field evaluation of a rapid point-of-care assay for targeting ant | Michel, C. E. C.; Solomon, A. W.; Magb |
| 145 | Correlation of clinical trachoma and infection in aboriginal com | Michel, C. E. C.; Roper, K. G.; Divena, M |
| 146 | Defining seropositivity thresholds for use in trachoma eliminat | Migchelsen, S. J.; Martin, D. L.; Southis |
| 147 | Prevalence of trachoma and associated factors in the rural area | Alfonso Miller, H.; Lopez Mesa, C. B. d |
| 148 | How reliable is the clinical exam in detecting ocular chlamydial | Miller, K.; Schmidt, G.; Melese, M.; Ale |
| 149 | Clinical evidence of trachoma in Colombian Amerindians of the | Miller, H.; Gallego, G.; Rodriguez, G. |
| 150 | A population-based trachoma prevalence survey covering seve | Missamou, F.; Marlhand, H.; Dzabatou |
| 151 | Trachoma and ocular Chlamydia trachomatis rates in children i | Mkocha, H.; Munoz, B.; West, S. |
| 152 | Follow-up and report on active trachoma in Zabol, Iran, promp | Mohammadi, S. F.; Bazi-Shad, A.; Akba |
| 153 | Prevalence of trachoma in Yobe State, North-Eastern Nigeria | Mpyet, C.; Ogoshi, C.; Goyol, M. |
| 154 | Personal and environmental risk factors for active trachoma in | Mpyet, C.; Goyol, M.; Ogoshi, C. |
| 155 | Prevalence of and risk factors for trachoma in Kano State, Nige | Mpyet, C.; Lass, B. D.; Yahaya, H. B.; So |
| 156 | Prevalence of trachoma in the Area Councils of the Federal Cap | Muhammad, N.; Mpyet, C.; Adamu, M |
| 157 | Can clinical signs of trachoma be used after multiple rounds of | Munoz, B.; Stare, D.; Mkocha, H.; Gay |
| 158 | Longitudinal trends in trachoma over eight years in a hyperend | Nash, S. D.; Sata, E.; Stewart, A. E. P.; A |
| 159 | Trachoma prevalence remains below threshold in five districts | Nash, S. D.; Stewart, A. E. P.; Astale, T. |
| 160 | Ocular Chlamydia trachomatis infection under the surgery, ant | Nash, S. D.; Stewart, A. E. P.; Zerihun, |
| 161 | Ocular chlamydia trachomatis infection and infectious load am | Nash, S. D.; Chernet, A.; Moncada, J.; S |
| 162 | Population-based prevalence of ocular Chlamydia trachomatis | Nash, S. D.; Chernet, A.; Astale, T.; Sat |
| 163 | Human conjunctival transcriptome analysis reveals the promin | Natividad, A.; Freeman, T. C.; Jeffries, |
| 164 | Evaluation of photography using head-mounted display techno | Naufal, F.; Brady, C. J.; Wolle, M. A.; Ka |
| 165 | Association of environmental risk factors and trachoma in Gash | Ndisabiye, D.; Gahungu, A.; Kayugi, D. |
| 166 | Trachoma in the province of Ouarzazate Morocco. [French] | Negrel, A. D.; Khazraji, Y. C.; Akalay, O |
| 167 | Comparison of smartphone photography, single-lens reflex pho | Nesemann, J. M.; Seider, M. I.; Snyder |
| 168 | Prevalence of trachoma in alto amazonas, loreto department, | Nesemann, J.; Munoz, M. B.; Morocho |
| 169 | The epidemiology of trachoma in Eastern Equatoria and Upper | Ngondi, J.; Onsarigo, A.; Adamu, L.; Ma |
| 170 | Effect of 3 years of SAFE (surgery, antibiotics, facial cleanliness | Ngondi, J.; Onsarigo, A.; Matthews, F.; |
| 171 | Blinding trachoma in postconflict southern Sudan | Ngondi, J.; Ole-Sempele, F.; Onsarigo, |
| 172 | Risk factors for active trachoma in children and trichiasis in adu | Ngondi, J.; Teshome, Gebre; Shargie, E |
| 173 | Evaluation of three years of the SAFE strategy (Surgery, Antibio | Ngondi, J.; Gebre, T.; Shargie, E. B.; Ad |
| 174 | Estimation of effects of community intervention with Antibioti | Ngondi, J.; Teshome, Gebre; Shargie, E |
| 175 | Prevalence and associated factors of active trachoma among c | Nigusie, A.; Berhe, R.; Gedefaw, M. |
| 176 | Prevalence of trachoma in the Far North region of Cameroon: | Noatina, B. N.; Kagmeni, G.; Mengouo |
| 177 | Prevalence of trachoma in the north region of Cameroon: resu | Noatina, B. N.; Kagmeni, G.; Souleyma |
| 178 | Serology, infection, and clinical trachoma as tools in prevalenc | Odonkor, M.; Naufal, F.; Munoz, B.; M |
| 179 | The Impact of Image Quality and Trachomatous Inflammation | Odonkor, M.; Naufal, F.; Mkocha, H.; F |
| 180 | Comparison of mass azithromycin coverage targets of children | Oldenburg, C. E.; Amza, A.; Kadri, B.; N |
| 181 | Active trachoma and community use of sanitation, Ethiopia | Oswald, W. E.; Stewart, A. E. P.; Krame |
| 182 | The burden of and risk factors for trachoma in selected district | Phiri, I.; Manangazira, P.; Macleod, C. |
| 183 | Follicular trachoma and trichiasis prevalence in an urban comm | Quicke, E.; Sillah, A.; Harding-Esch, E. |
| 184 | Ocular immune responses, Chlamydia trachomatis infection an | Ramadhani, A. M.; Derrick, T.; Macleo |
| 185 | Prevalence of trachoma in Jigawa State, Northwestern Nigeria | Ramyil, A.; Wade, P.; Ogoshi, C.; Goyo |
| 186 | Prevalence and associated factors of active trachoma among 1 | Reda, G.; Yemane, D.; Gebreyesus, A. |

| | | |
|---|---|---|
| 187 | Preliminary evidence that synanthropic flies contribute to the t | Reilly, L. A.; Favacho, J.; Garcez, L. M.; |
| 188 | Risk of trachoma in a SAFE intervention area | Roba, A. A.; Patel, D.; Zondervan, M. |
| 189 | Trachoma among children in western Yemen | D.S, Rosen |
| 190 | Prevalence and risk factors for trachoma in Rwanda | Ruberanziza, E.; Mupfasoni, D.; Nizeyi |
| 191 | Associated factors of the co-occurrence of trachoma and soil-t | Saboya-Diaz, M. I.; Angeles, C. A. C.; Sc |
| 192 | Burden of trachoma in five counties of Eastern Equatoria state | Sanders, A. M.; Stewart, A. E. P.; Makc |
| 193 | Progress toward elimination of trachoma as a public health pro | Sanders, A. M.; Abdalla, Z.; Elshafie, B. |
| 194 | Prevalence of trachoma within refugee camps serving South Su | Sanders, A. M.; Abdalla, Z.; Elshafie, B. |
| 195 | Prevalence and spatial distribution of trachoma among schoolc | Schellini, S. A.; Lavezzo, M. M.; Ferraz, |
| 196 | Mapping trachoma in Mali: Results of a national survey. [Frenc | Schemann, J. F.; Sacko, D.; Banou, A.; |
| 197 | How reliable are tests for trachoma?--a latent class approach | See, C. W.; Alemayehu, W.; Melese, M |
| 198 | Serological and PCR-based markers of ocular Chlamydia tracho | Senyonjo, L. G.; Debrah, O.; Martin, D. |
| 199 | Common eye diseases in children of rural community in Goro c | Mohammed, Shaffi; Abebe, Bejiga |
| 200 | School-Based versus Community-Based Sampling for Trachoma | Sheehan, J. P.; Gebresillasie, S.; Shifera |
| 201 | Cohort and age effects of mass drug administration on prevale | Shekhawat, N.; Mkocha, H.; Munoz, B. |
| 202 | Prevalence of active trachoma and associated factors among s | Shimelash, A.; Alemayehu, M.; Dagne, |
| 203 | PrevalÃªncia de Tracoma em crianÃ§as em idade escolar no m | Silva, Evanildo JosÃ© da; Oliveira, Lay |
| 204 | Relationship between trachoma and chronic and acute malnut | Smith, A. G.; Broman, A. T.; Alemayeh |
| 205 | Smartphone photography as a possible method of post-validat | Snyder, B. M.; Sie, A.; Tapsoba, C.; Dah |
| 206 | Rational use of azithromycin in the control of trachoma : using | Solomon, A. |
| 207 | Two doses of azithromycin to eliminate trachoma in a Tanzania | Solomon, A. W.; Harding-Esch, E.; Alex |
| 208 | Design and baseline data of a randomized trial to evaluate cov | Stare, D.; Harding-Esch, E.; Munoz, B.; |
| 209 | Progress to eliminate trachoma as a public health problem in A | Stewart, A. E. P.; Mulat, Zerihun; Dem |
| 210 | Effect of water, sanitation and hygiene interventions on active | Tadesse, B.; Worku, A.; Kumie, A.; Yim |
| 211 | The burden of and risk factors for active trachoma in the North | Beselam, Tadesse; Alemayehu, Worku |
| 212 | Rapid assessment of trachoma in underserved population of Ca | Praveen, Vashist; Noopur, Gupta; Rath |
| 213 | Etiological characteristics of chlamydia trachoma conjunctivitis | Wang, M.; Lu, X.; Hu, A.; Zhang, M.; Li, |
| 214 | Nonocular chlamydia infection and risk of ocular reinfection af | West, S.; Munoz, B.; Bobo, L.; Quinn, T |
| 215 | Progression of active trachoma to scarring in a cohort of Tanza | West, S. K.; Munoz, B.; Mkocha, H.; Hs |
| 216 | Trachoma and ocular Chlamydia trachomatis were not elimina | West, S. K.; Munoz, B.; Mkocha, H.; Ga |
| 217 | Do infants increase the risk of re-emergent infection in househ | West, S. K.; Stare, D.; Mkocha, H.; Mu |
| 218 | A randomized trial of two coverage targets for mass treatment | West, S. K.; Bailey, R.; Munoz, B.; Edw |
| 219 | Risk of infection with Chlamydia trachomatis from migrants to | West, S. K.; Munoz, B. E.; Mkocha, H.; |
| 220 | Can we use antibodies to Chlamydia trachomatis as a surveillar | West, S. K.; Munoz, B.; Weaver, J.; Mr |
| 221 | Treating village newcomers and travelers for trachoma: results | West, S. K.; Munoz, B.; Mkocha, H.; Di |
| 222 | Surveillance surveys for reemergent trachoma in formerly end | West, S. K.; Zambrano, A. I.; Sharma, S |
| 223 | Longitudinal change in the serology of antibodies to Chlamydia | West, S. K.; Munoz, B.; Kaur, H.; Dize, |
| 224 | Evidence for contamination with C. trachomatis in the househo | West, S. K.; Nanji, A. A.; Mkocha, H.; M |
| 225 | The effect of mass drug administration for trachoma on antibo | West, S. K.; Munoz, B.; Mkocha, H.; Ga |
| 226 | Evaluation of a single dose of azithromycin for trachoma in low | Wilson, N.; Goodhew, B.; Mkocha, H.; |
| 227 | Prevalence of active trachoma and associated factors among c | Woldekidan, E.; Daka, D.; Legesse, D.; |
| 228 | Screening for ocular abnormalities and subnormal vision in sch | Yoseph, Worku; Samson, Bayu |
| 229 | A cross-sectional population-based survey of trachoma among | Xue, WenWen; Lu, LiNa; Zhu, JianFeng |
| 230 | Comparison of an rRNA-based and DNA-based nucleic acid am | Yang, J. L.; Schachter, J.; Moncada, J.; |
| 231 | Detection of Chlamydia trachomatis ocular infection in trachor | Yang, J. L.; Hong, K. C.; Schachter, J.; M |
| 232 | Blinding trachoma: results of a prevalence survey in 8 health d | Yaya, G.; Kemata, B.; Baanam, M. Y.; B |
| 233 | Achieving trachoma control in Ghana after implementing the S | Yayemain, D.; King, J. D.; Debrah, O.; E |

| | | |
|---|---|---|
| 234 | The impact of water supply on trachoma prevalence | Yilkal, Alemu; Abebe, Bejiga |
| 235 | Can we stop mass drug administration prior to 3 annual rounds | Yohannan, J.; Munoz, B.; Mkocha, H.; |
| 236 | The World Health Organization Recommendations for Trachom | Zambrano, A. I.; Sharma, S.; Crowley, |
| 237 | Measuring trachomatous inflammation-intense (TI) when prev | Zambrano, A. I.; Munoz, B. E.; Mkocha |
| 238 | Trachoma in Jimma zone, South Western Ethiopia | Zerihun, N. |

| Published | Journal | Volume | Issue | Pages |
|---|---|---|---|---|
| 2020 | Clinical Ophthalmology | 14 | | 3709-3718 |
| 2007 | British Journal of Ophthalmology | 91 | 1 | 13-17 |
| 2017 | Current Pediatric Research | 21(3) | | 507-513 |
| 2018 | Ethiopian Journal of Health Development | 32 | 2 | unpaginate |
| 2007 | Ethiopian Medical Journal | 45 | 2 | 181-186 |
| 2001 | British Journal of Ophthalmology | 85 | 12 | 1406-1410 |
| 2005 | Transactions of the Royal Society of Tropical Medicine and Hygiene | 99 | 11 | 840-843 |
| 2018 | Special Issue: 2017 Global Trachoma Mapping Project. | 25 | Suppl. 1 | 70-78 |
| 2010 | PLoS Neglected Tropical Diseases | 4 | 11 | e895 |
| 2012 | PLoS Neglected Tropical Diseases | 6(4) (no pagination) | | |
| 2013 | PLoS Neglected Tropical Diseases | 7(1) (no pagination) | | |
| 2017 | Clinical Infectious Diseases | 64 | 6 | 743-750 |
| 2018 | British Journal of Ophthalmology | 102 | 5 | 680-686 |
| 2019 | Ophthalmic Epidemiology | 26 | 4 | 231-237 |
| 2006 | Ophthalmic Epidemiology | 13 | 4 | 227-236 |
| 2011 | PLoS Neglected Tropical Diseases | 5(12) (no pagination) | | |
| 2021 | Italian Journal of Pediatrics | 47(1) (no pagination) | | |
| 2010 | Bulletin de la Societe belge d'ophtalmologie | | 316 | 37-42 |
| 2011 | Medecine tropicale : revue du Corps de sante colonial | 71(5) | | 515-516 |
| 2005 | Ophthalmic Epidemiology | 12 | 5 | 311-9 |
| 1994 | Journal of Infectious Diseases | 170 | 3 | 709-712 |
| 2010 | Tropical Medicine and International Health | 15 | 3 | 306-311 |
| 2010 | PLoS Neglected Tropical Diseases | 4 | 7 | e734 |
| 2001 | Ophthalmic Epidemiology | 8 | 44960 | 119-25 |
| 2020 | BMJ Open Ophthalmology | 5 | 1 | |
| 2007 | Ethiopian Journal of Health Development | 21 | 3 | 211-215 |
| 2012 | British Journal of Ophthalmology | 96 | 10 | 1350-1351 |
| 2013 | American Journal of Tropical Medicine and Hygiene | 1) | | 12 |
| 2017 | Ophthalmic Epidemiology | 24 | 4 | 265-273 |
| 1991 | Lancet | 338 | Oct. 5 | 847-850 |
| 2021 | Investigative Ophthalmology and Visual Science. Conference: Annua | 62 | 8 | |
| 2013 | PLoS Neglected Tropical Diseases | 7 | 7 | e2347 |
| 2019 | PLoS Neglected Tropical Diseases | 13 | 10 | e0007749 |
| 2010 | PLoS Neglected Tropical Diseases | 4 | 10 | e835 |
| 2011 | Infection and Immunity | 79 | 12 | 4977-4983 |
| 2016 | PLoS Neglected Tropical Diseases | 10 | 9 | e0004863 |
| 2017 | | | | |
| 2018 | Wellcome Open Research | 3 | | |
| 2020 | Journal of Infection | 80 | 4 | 454-461 |
| 2006 | Ophthalmic Epidemiology | 13(6) | | 365-370 |
| 2017 | PLoS Neglected Tropical Diseases | 11 | 9 | e0005863 |
| 2022 | PLoS Neglected Tropical Diseases | 16(7) (no pagination) | | |
| 2007 | British Journal of Ophthalmology | 91 | 5 | 667-672 |
| 2016 | Transactions of the Royal Society of Tropical Medicine and Hygiene | 110 | 10 | 582-587 |
| 2016 | PLoS Neglected Tropical Diseases | 10 | 7 | e0004798 |

| | | | | |
|---|---|---|---|---|
| 2017 | Investigative Ophthalmology and Visual Science. Conference | 58 | 8 | |
| 2014 | Transactions of the Royal Society of Tropical Medicine and Hygiene | 108 | 1 | 42-48 |
| 2005 | Transactions of the Royal Society of Tropical Medicine and Hygiene | 99 | 2 | 120-127 |
| 2008 | International Journal of Epidemiology | 37 | 3 | 549-558 |
| 2009 | Arquivos Brasileiros de Oftalmologia | 72 | 3 | 355-359 |
| 1994 | Sexually Transmitted Diseases | 21 | 1 | 44930 |
| 2017 | PLoS Neglected Tropical Diseases | 11 | 12 | e0006099 |
| 2016 | Parasites and Vectors | 9 | 30 | |
| 2016 | BMC Infectious Diseases | 16 | 60 | |
| 2020 | Parasites and Vectors | 13 | 533 | |
| 2018 | Seminars in Ophthalmology | 33 | 2 | 219-222 |
| 2012 | Tropical Medicine and International Health | 17 | 7 | 896-903 |
| 2018 | Special Issue: 2017 Global Trachoma Mapping Project. | 25 | Suppl. 1 | 25-32 |
| 2012 | PLoS Neglected Tropical Diseases | 6 | 4 | e1585 |
| 2013 | Ethiopian Journal of Health Sciences | 23 | 1 | 44935 |
| 2016 | Ophthalmic Epidemiology | 23 | 6 | 347-353 |
| 2005 | Clinical and Experimental Immunology | 142 | 2 | 347-353 |
| 2006 | PLoS Medicine / Public Library of Science | 3 | 8 | e266 |
| 2018 | PLoS Neglected Tropical Diseases | 12(2) (no pagination) | | |
| 2006 | Transactions of the Royal Society of Tropical Medicine and Hygiene | 100 | 2 | 149-157 |
| 2003 | | | | |
| 2017 | Frontiers in Cellular and Infection Microbiology | 6 | JAN | |
| 2017 | Infectious Diseases of Poverty | 6(1) (no pagination) | | |
| 2016 | Ophthalmic Epidemiology | 23(6) | | 354-359 |
| 2014 | Journal of Water, Sanitation and Hygiene for Development | 4 | 1 | 120-130 |
| 2022 | PLoS ONE [Electronic Resource] | 17(6 June) (no pagination) | | |
| 2018 | Scientific Reports | 8 | 1 | 7432 |
| 2021 | PLoS Neglected Tropical Diseases | 15 | 8 | |
| 2006 | Ophthalmic Epidemiology | 13(5) | | 335-342 |
| 2012 | Tropical Medicine and Health | 40 | 1 | 45121 |
| 2014 | Tropical Medicine and Health | 42 | 1 | 43-52 |
| 2009 | PLoS ONE [Electronic Resource] | | August | e6702 |
| 2006 | Investigative Ophthalmology & Visual Science | 47 | 11 | 4767-4771 |
| 2018 | American Journal of Tropical Medicine & Hygiene | 98 | 1 | 216-220 |
| 2019 | Diagnostic Microbiology and Infectious Disease | 93 | 4 | 293-298 |
| 2021 | Scientific Reports | 11 | 3 | |
| 2021 | American Journal of Tropical Medicine and Hygiene | 105 | 5 | 1362-1367 |
| 2021 | American Journal of Tropical Medicine and Hygiene | 104(1) | | 207-215 |
| 2010 | Ophthalmic Epidemiology | 17 | 6 | 343-348 |
| 2009 | Ghana Medical Journal | 43 | 2 | 54-60 |
| 2016 | Special Issue: Trachoma mapping. | 23 | Suppl. 1 | 84-93 |
| 2022 | American Journal of Tropical Medicine & Hygiene | 28 | | 28 |
| 2008 | Transactions of the Royal Society of Tropical Medicine and Hygiene | 102 | 12 | 1255-1262 |
| 2009 | PLoS Neglected Tropical Diseases | 3 | 12 | e573 |
| 2010 | PLoS Neglected Tropical Diseases | 4 | 11 | e861 |
| 2013 | PLoS Neglected Tropical Diseases | 7 | 6 | e2115 |
| 2017 | BMC Public Health | 18 | 1 | 62 |

| | | | | |
|---|---|---|---|---|
| 2019 | Parasites and Vectors | 12 | 497 | |
| 1992 | Current Eye Research | 11 | 3 | 279-83 |
| 2010 | British Journal of Ophthalmology | 94 | 2 | 157-160 |
| 2013 | PLoS Neglected Tropical Diseases | 7 | 7 | e2265 |
| 2008 | Ophthalmic Epidemiology | 15 | 5 | 294-302 |
| 2013 | American Journal of Tropical Medicine and Hygiene | 1) | | 45210 |
| 2010 | PLoS ONE [Electronic Resource] | | February | e9067 |
| 2014 | Health | 6 | 1 | 57-63 |
| 2020 | PLoS Neglected Tropical Diseases | 14 | 10 | |
| 2019 | BMC Infectious Diseases | 19(1) (no pagination) | | |
| 2020 | American Journal of Tropical Medicine and Hygiene | 103(5 SUPPL) | | 365 |
| 1996 | British Journal of Ophthalmology | 80(12) | | 1037-1041 |
| 2021 | Open Ophthalmology Journal | 15 | 1 | 108-116 |
| 2012 | Investigative Ophthalmology & Visual Science | 53 | 1 | 235-40 |
| 2018 | PLoS Medicine / Public Library of Science | 15 | 8 | e1002633 |
| 2012 | BMC Public Health | 12 | | 1105 |
| 2003 | Eastern Mediterranean Health Journal | 9 | 45052 | 1026-1033 |
| 2005 | Eastern Mediterranean Health Journal | 11 | 3 | 402-409 |
| 2009 | Ophthalmic Epidemiology | 16 | 4 | 206-211 |
| 2019 | PLoS Neglected Tropical Diseases | 13 | 1 | e0007127 |
| 2008 | PLoS Neglected Tropical Diseases | 2 | 9 | e299 |
| 2010 | British Journal of Ophthalmology | 94 | 1 | 14-19 |
| 2013 | International Health (RSTMH) | 5 | 4 | 280-287 |
| 2014 | PLoS Neglected Tropical Diseases | 8 | 3 | e2732 |
| 2011 | Ophthalmic Epidemiology | 18 | 4 | 150-157 |
| 2011 | American Journal of Tropical Medicine and Hygiene | 1) | | 168 |
| 2009 | PLoS Neglected Tropical Diseases | 3 | 7 | e492 |
| 1999 | Medical Journal of Australia | 170 | 4 | 190 |
| 2014 | PLoS Neglected Tropical Diseases | 8 | 6 | e2900 |
| 2013 | Journal of Clinical Microbiology | 51 | 7 | 2195-2203 |
| 2015 | American Journal of Tropical Medicine and Hygiene | 93(4 Supplement) | | 403-404 |
| 2017 | Parasites and Vectors | 10 | 624 | |
| 2013 | Investigative Ophthalmology and Visual Science. Conference | 54 | 15 | |
| 2014 | PLoS Neglected Tropical Diseases | 8 | 4 | e2761 |
| 2016 | Special Issue: Etiology and prevention of trachoma. | 59 | 6 | 548-554 |
| 2015 | Parasites and Vectors | 8 | 535 | |
| 2016 | Ophthalmic Epidemiology | 23 | 6 | 360-365 |
| 2022 | Medical Journal of Australia. | | | |
| 2022 | PLoS Neglected Tropical Diseases | 16 | 4 | |
| 2022 | Australian and New Zealand journal of public health | 46 | 2 | 155-160 |
| 1991 | Journal of Clinical Pathology | 44 | 4 | 285-9 |
| 2019 | Transactions of the Royal Society of Tropical Medicine and Hygiene | 113 | 10 | 599-609 |
| 2020 | Clinical Infectious Diseases | 72 | 3 | 423-430 |
| 2003 | Cahiers Sante | 13(1) | | 45184 |
| 2016 | PLoS ONE [Electronic Resource] | 11 | 7 | e0158625 |
| 2015 | Scientific Reports | 5 | | 18532 |
| 2015 | PLoS Neglected Tropical Diseases | 9 | 2 | e0003555 |

| | | | | |
|---|---|---|---|---|
| 2009 | British Journal of Ophthalmology | 93 | 7 | 866-870 |
| 1998 | Vigilancia epidemiologica do tracoma em instituicao de ensino na ci | 32 | 1 | 59-63 |
| 2016 | Special Issue: Trachoma mapping. | 23 | Suppl. 1 | 44992 |
| 2006 | Ophthalmic Epidemiology | 13 | 3 | 173-181 |
| 2006 | Lancet | 367(9522) | | 1585-1590 |
| 2011 | PLoS Neglected Tropical Diseases | 5 | 3 | e986 |
| 2017 | PLoS Neglected Tropical Diseases | 11 | 1 | e0005230 |
| 2020 | PLoS ONE [Electronic Resource] | 15 | 5 | |
| 2004 | Ophthalmic Epidemiology | 11 | 3 | 255-62 |
| 2010 | Biomedica | 30(3) | | 432-439 |
| 2018 | Special Issue: 2017 Global Trachoma Mapping Project. | 25 | Suppl. 1 | 155-161 |
| 2009 | Tanzania journal of health research | 11 | 3 | 103-110 |
| 2017 | New Microbes and New Infections | 20 | | 14-15 |
| 2008 | Ophthalmic Epidemiology | 15 | 5 | 303-307 |
| 2010 | Tropical Medicine and International Health | 15 | 2 | 168-172 |
| 2012 | PLoS ONE [Electronic Resource] | 7 | 7 | e40421 |
| 2018 | Special Issue: 2017 Global Trachoma Mapping Project. | 25 | Suppl. 1 | 45247 |
| 2011 | Investigative Ophthalmology & Visual Science | 52 | 12 | 8806-10 |
| 2017 | American Journal of Tropical Medicine and Hygiene | 97(5 Supplement 1) | | 228 |
| 2018 | Transactions of the Royal Society of Tropical Medicine and Hygiene | 112(12) | | 538-545 |
| 2018 | Clinical Infectious Diseases | 67 | 12 | 1840-1846 |
| 2020 | PLoS Neglected Tropical Diseases | 14(5) | | 44942 |
| 2022 | American Journal of Tropical Medicine and Hygiene | 106 | 1 | 62-65 |
| 2010 | Infection and Immunity | 78 | 11 | 4895-4911 |
| 2021 | PLoS Neglected Tropical Diseases | 15 | 11 | |
| 2020 | African Health Sciences | 20 | 1 | 182-189 |
| 1992 | Bulletin of the World Health Organization | 70(4) | | 451-456 |
| 2020 | American Journal of Tropical Medicine and Hygiene | 103(6) | | 2488-2491 |
| 2021 | Investigative Ophthalmology and Visual Science. Conference: Annua | 62 | 8 | |
| 2005 | Bulletin of the World Health Organization | 83 | 12 | 904-912 |
| 2006 | Lancet (British edition) | 368 | 9535 | 589-595 |
| 2006 | PLoS Medicine | 3 | 12 | e478 |
| 2008 | Transactions of the Royal Society of Tropical Medicine and Hygiene | 102 | 5 | 432-438 |
| 2009 | Transactions of the Royal Society of Tropical Medicine and Hygiene | 103(10) | | 1001-1010 |
| 2010 | British Journal of Ophthalmology | 94 | 3 | 278-281 |
| 2015 | BMC Research Notes | 8 | | 641 |
| 2013 | PLoS Neglected Tropical Diseases | 7 | 5 | e2240 |
| 2014 | PLoS Neglected Tropical Diseases | 8 | 6 | e2932 |
| 2021 | PLoS Neglected Tropical Diseases | 15 | 4 | |
| 2022 | Translational Vision Science & Technology | 11 | 3 | 11 |
| 2018 | American Journal of Tropical Medicine and Hygiene | 98 | 2 | 389-395 |
| 2017 | Bulletin of the World Health Organization | 95 | 4 | 250-260 |
| 2018 | Special Issue: 2017 Global Trachoma Mapping Project. | 25 | Suppl. 1 | 181-191 |
| 2013 | Tropical Medicine and International Health | 18 | 11 | 1344-1352 |
| 2019 | PLoS Neglected Tropical Diseases | 13 | 7 | e0007559 |
| 2015 | Special Issue: Trachoma. | 22 | 3 | 184-189 |
| 2020 | BMC Ophthalmology | 20(1) | | 144 |

| | | | | |
|---|---|---|---|---|
| 2007 | Cadernos de Saude Publica | 23 | 7 | 1682-1688 |
| 2013 | International Ophthalmology | 33 | 1 | 53-59 |
| 1995 | | 3 | | 45245 |
| 2009 | East African Journal of Public Health | 6 | 3 | 287-91 |
| 2022 | PLoS Neglected Tropical Diseases | 16 | 7 | |
| 2017 | PLoS Neglected Tropical Diseases | 11 | 6 | e0005658 |
| 2019 | American Journal of Tropical Medicine and Hygiene | 101 | 6 | 1296-1302 |
| 2019 | PLoS Neglected Tropical Diseases | 13 | 6 | e0007491 |
| 2010 | Prevalencia e localizacao espacial dos casos de tracoma detectados | 73 | 4 | 358-362 |
| 1998 | Bulletin of the World Health Organization | 76(6) | | 599-606 |
| 2011 | Investigative Ophthalmology & Visual Science | 52 | 9 | 6133-7 |
| 2018 | PLoS Neglected Tropical Diseases | 12 | 12 | e0007027 |
| 2005 | Ethiopian Journal of Health Development | 19 | 2 | 148-152 |
| 2018 | American Journal of Tropical Medicine and Hygiene | 99(1) | | 150-154 |
| 2014 | Investigative Ophthalmology & Visual Science | 55 | 4 | 2307-2314 |
| 2022 | Italian Journal of Pediatrics | 48 | 1 | 61 |
| 2016 | Rev. bras. oftalmol | 75 | 3 | 181-184 |
| 2007 | Journal of Tropical Pediatrics | 53(5) | | 308-312 |
| 2019 | International Health | 11(6) | | 613-615 |
| 2003 | | | | |
| 2008 | New England Journal of Medicine | 358 | 17 | 1870-1871 |
| 2011 | Ophthalmic Epidemiology | 18 | 1 | 20-29 |
| 2019 | American Journal of Tropical Medicine and Hygiene | 101 | 6 | 1286-1295 |
| 2017 | PLoS Neglected Tropical Diseases | 11(11) (no pagination) | | |
| 2017 | Infectious Diseases of Poverty | 6 | 143 | |
| 2013 | PLoS ONE [Electronic Resource] | 8 | 6 | e65918 |
| 2016 | Science China | Life sciences. 59(6) | | 555-560 |
| 1993 | Investigative Ophthalmology and Visual Science | 34(11) | | 3194-3198 |
| 2001 | Ophthalmic Epidemiology | 8 | 44960 | 137-44 |
| 2007 | Investigative Ophthalmology & Visual Science | 48 | 4 | 1492-1497 |
| 2011 | Investigative Ophthalmology & Visual Science | 52 | 8 | 6040-2 |
| 2013 | PLoS Neglected Tropical Diseases | 7 | 8 | e2415 |
| 2015 | Special Issue: Trachoma. | 22 | 3 | 170-175 |
| 2016 | PLoS Neglected Tropical Diseases | 10 | 1 | e0004352 |
| 2017 | PLoS ONE [Electronic Resource] | 12 | 6 | e0178595 |
| 2017 | JAMA Ophthalmology | 135 | 11 | 1141-1146 |
| 2018 | Scientific Reports | 8 | 1 | 3520 |
| 2019 | PLoS Neglected Tropical Diseases | 13 | 12 | |
| 2020 | Scientific Reports | 10 | 9 | |
| 2019 | Ophthalmic Epidemiology | 26 | 1 | 44932 |
| 2019 | BMC Infectious Diseases | 19(1) (no pagination) | | |
| 2002 | Ethiopian Journal of Health Development | 16 | 2 | 165-171 |
| 2016 | BioMed Research International | 2016 | | Article-869 |
| 2007 | British Journal of Ophthalmology | 91(3) | | 293-295 |
| 2009 | Investigative Ophthalmology and Visual Science | 50(1) | | 90-94 |
| 2015 | Le trachome evolutif: resultats d'une enquete de prevalence dans h | 108 | 4 | 299-304 |
| 2009 | Transactions of the Royal Society of Tropical Medicine and Hygiene | 103 | 10 | 993-1000 |

| | | | | |
|---|---|---|---|---|
| 2004 | Ethiopian Medical Journal | 42 | 3 | 179-184 |
| 2013 | JAMA Ophthalmology | 131 | 4 | 431-6 |
| 2016 | PLoS Neglected Tropical Diseases | 10(9) (no pagination) | | |
| 2017 | Investigative Ophthalmology & Visual Science | 58 | 2 | 997-1000 |
| 1997 | Tropical Medicine and International Health | 2 | 12 | 1115-1121 |

| Digital Object Indentifier (DOI) (where available) |
|---|
| https://dx.doi.org/10.2147/OPTH.S282567 |
| http://dx.doi.org/10.1136/bjo.2006.099507 |
| ed |
| http://dx.doi.org/10.1136/bjo.85.12.1406 |
| http://dx.doi.org/10.1016/j.trstmh.2005.06.013 |
| http://dx.doi.org/10.1080/09286586.2018.1446536 |
| http://dx.doi.org/10.1371/journal.pntd.0000895 |
| https://dx.doi.org/10.1371/journal.pntd.0001586 |
| https://dx.doi.org/10.1371/journal.pntd.0001983 |
| http://dx.doi.org/10.1136/bjophthalmol-2017-310916 |
| http://dx.doi.org/10.1080/09286586.2019.1597129 |
| http://dx.doi.org/10.1080/09286580600718974 |
| https://dx.doi.org/10.1371/journal.pntd.0001441 |
| https://dx.doi.org/10.1186/s13052-021-01064-x |
| http://dx.doi.org/10.1093/infdis/170.3.709 |
| http://dx.doi.org/10.1111/j.1365-3156.2009.02459.x |
| http://dx.doi.org/10.1371/journal.pntd.0000734 |
| 10.1136/bmjophth-2020-000531 |
| 10.1136/bjophthalmol-2012-302050 |
| http://dx.doi.org/10.1080/09286586.2017.1279337 |
| http://dx.doi.org/10.1371/journal.pntd.0002347 |
| http://dx.doi.org/10.1371/journal.pntd.0007749 |
| http://dx.doi.org/10.1371/journal.pntd.0000835 |
| http://dx.doi.org/10.1128/IAI.05718-11 |
| 10.12688/wellcomeopenres.13423.1 |
| http://dx.doi.org/10.1016/j.jinf.2020.01.015 |
| https://dx.doi.org/10.1080/09286580601013078 |
| http://dx.doi.org/10.1371/journal.pntd.0005863 |
| https://dx.doi.org/10.1371/journal.pntd.0010629 |
| http://dx.doi.org/10.1136/bjo.2006.099275 |
| http://dx.doi.org/10.1093/trstmh/trw069 |

| |
|---|
| http://dx.doi.org/10.1093/trstmh/trt101 |
| http://dx.doi.org/10.1016/j.trstmh.2004.03.011 |
| http://dx.doi.org/10.1093/ije/dyn045 |
| http://dx.doi.org/10.1590/S0004-27492009000300014 |
| http://dx.doi.org/10.1371/journal.pntd.0006099 |
| http://dx.doi.org/10.1186/s13071-020-04414-6 |
| 10.1080/08820538.2016.1208762 |
| http://dx.doi.org/10.1111/j.1365-3156.2012.03004.x |
| http://dx.doi.org/10.1080/09286586.2017.1409358 |
| http://dx.doi.org/10.1371/journal.pntd.0001585 |
| http://dx.doi.org/10.1080/09286586.2016.1238947 |
| http://dx.doi.org/10.1111/j.1365-2249.2005.02917.x |
| https://dx.doi.org/10.1371/journal.pntd.0006282 |
| http://dx.doi.org/10.1016/j.trstmh.2005.06.029 |
| 10.3389/fcimb.2016.00207 |
| https://dx.doi.org/10.1186/s40249-017-0345-8 |
| https://dx.doi.org/10.3109/09286586.2015.1131305 |
| http://dx.doi.org/10.2166/washdev.2013.060 |
| https://dx.doi.org/10.1371/journal.pone.0268441 |
| http://dx.doi.org/10.1038/s41598-018-23887-1 |
| http://dx.doi.org/10.1371/journal.pntd.0009655 |
| https://dx.doi.org/10.1080/09286580600943796 |
| http://dx.doi.org/10.2149/tmh.2011-26 |
| http://dx.doi.org/10.2149/tmh.2013-26 |
| http://dx.doi.org/10.1371/journal.pone.0006702 |
| https://dx.doi.org/10.4269/ajtmh.17-0102 |
| http://dx.doi.org/10.1016/j.diagmicrobio.2018.11.001 |
| http://dx.doi.org/10.1038/s41598-021-86639-8 |
| http://dx.doi.org/10.4269/ajtmh.21-0541 |
| https://dx.doi.org/10.4269/AJTMH.20-0777 |
| http://dx.doi.org/10.3109/09286586.2010.528132 |
| https://dx.doi.org/10.4269/ajtmh.21-1140 |
| http://dx.doi.org/10.1016/j.trstmh.2008.04.022 |
| http://dx.doi.org/10.1371/journal.pntd.0000573 |
| http://dx.doi.org/10.1371/journal.pntd.0000861 |
| http://dx.doi.org/10.1371/journal.pntd.0002115 |
| https://dx.doi.org/10.1186/s12889-017-4605-0 |

| |
|---|
| http://dx.doi.org/10.1136/bjo.2009.161513 |
| http://dx.doi.org/10.1371/journal.pntd.0002265 |
| http://dx.doi.org/10.1080/09286580802256542 |
| http://dx.doi.org/10.1371/journal.pone.0009067 |
| http://dx.doi.org/10.4236/health.2014.61009 |
| http://dx.doi.org/10.1371/journal.pntd.0008708 |
| https://dx.doi.org/10.1186/s12879-019-3992-5 |
| http://dx.doi.org/10.1136/bjo.80.12.1037 |
| 10.2174/1874364102115010108 |
| https://dx.doi.org/10.1167/iovs.11-8493 |
| https://dx.doi.org/10.1371/journal.pmed.1002633 |
| http://dx.doi.org/10.1080/09286580902999389 |
| http://dx.doi.org/10.1371/journal.pntd.0007127 |
| http://dx.doi.org/10.1371/journal.pntd.0000299 |
| http://dx.doi.org/10.1136/bjo.2009.165282 |
| http://dx.doi.org/10.1093/inthealth/iht027 |
| http://dx.doi.org/10.3109/09286586.2011.594204 |
| http://dx.doi.org/10.1371/journal.pntd.0000492 |
| http://dx.doi.org/10.1371/journal.pntd.0002900 |
| http://dx.doi.org/10.1128/JCM.00622-13 |
| http://dx.doi.org/10.1186/s13071-017-2566-x |
| http://dx.doi.org/10.1371/journal.pntd.0002761 |
| http://dx.doi.org/10.1007/s11427-016-5058-x |
| http://dx.doi.org/10.1080/09286586.2016.1244274 |
| https://dx.doi.org/10.5694/mja2.51735 |
| http://dx.doi.org/10.1371/journal.pntd.0010275 |
| http://dx.doi.org/10.1111/1753-6405.13179 |
| http://dx.doi.org/10.1093/trstmh/trz042 |
| http://dx.doi.org/10.1093/cid/ciaa042 |
| http://dx.doi.org/10.1371/journal.pone.0158625 |
| https://dx.doi.org/10.1038/srep18532 |
| http://dx.doi.org/10.1371/journal.pntd.0003555 |

| |
|---|
| http://dx.doi.org/10.1136/bjo.2008.151720 |
| http://dx.doi.org/10.1590/S0034-89101998000100008 |
| http://dx.doi.org/10.1080/09286580600611427 |
| https://dx.doi.org/10.1016/S0140-6736%2806%2968695-9 |
| http://dx.doi.org/10.1371/journal.pntd.0000986 |
| http://dx.doi.org/10.1371/journal.pntd.0005230 |
| http://dx.doi.org/10.1371/journal.pone.0229297 |
| http://dx.doi.org/10.1080/09286586.2018.1546878 |
| https://dx.doi.org/10.1016/j.nmni.2017.08.001 |
| http://dx.doi.org/10.1080/09286580802237633 |
| http://dx.doi.org/10.1111/j.1365-3156.2009.02436.x |
| http://dx.doi.org/10.1371/journal.pone.0040421 |
| http://dx.doi.org/10.1080/09286586.2017.1367409 |
| https://dx.doi.org/10.1167/iovs.11-8074 |
| https://dx.doi.org/10.1093/trstmh/try096 |
| http://dx.doi.org/10.1093/cid/ciy377 |
| https://dx.doi.org/10.1371/journal.pntd.0008226 |
| http://dx.doi.org/10.4269/ajtmh.21-0873 |
| http://dx.doi.org/10.1128/IAI.00844-10 |
| http://dx.doi.org/10.1371/journal.pntd.0009928 |
| http://dx.doi.org/10.4314/ahs.v20i1.23 |
| https://dx.doi.org/10.4269/ajtmh.20-0386 |
| http://dx.doi.org/10.1016/S0140-6736(06)69202-7 |
| http://dx.doi.org/10.1371/journal.pmed.0030478 |
| http://dx.doi.org/10.1016/j.trstmh.2008.02.014 |
| https://dx.doi.org/10.1016/j.trstmh.2008.11.023 |
| http://dx.doi.org/10.1136/bjo.2009.168260 |
| https://dx.doi.org/10.1186/s13104-015-1529-6 |
| http://dx.doi.org/10.1371/journal.pntd.0002240 |
| http://dx.doi.org/10.1371/journal.pntd.0002932 |
| http://dx.doi.org/10.1371/journal.pntd.0009343 |
| https://dx.doi.org/10.1167/tvst.11.3.11 |
| http://dx.doi.org/10.4269/ajtmh.17-0501 |
| http://dx.doi.org/10.2471/blt.16.177758 |
| http://dx.doi.org/10.1080/09286586.2017.1298823 |
| http://dx.doi.org/10.1111/tmi.12182 |
| http://dx.doi.org/10.1371/journal.pntd.0007559 |
| https://dx.doi.org/10.1186/s12886-020-01394-0 |

| |
|---|
| http://dx.doi.org/10.1590/S0102-311X2007000700020 |
| 10.1007/s10792-012-9632-3 |
| http://dx.doi.org/10.1371/journal.pntd.0010532 |
| http://dx.doi.org/10.1371/journal.pntd.0007491 |
| http://dx.doi.org/10.1590/S0004-27492010000400012 |
| https://dx.doi.org/10.1167/iovs.11-7419 |
| https://dx.doi.org/10.4269/ajtmh.17-0904 |
| http://dx.doi.org/10.1167/iovs.13-12701 |
| https://dx.doi.org/10.1186/s13052-022-01258-x |
| 10.5935/0034-7280.20160038 |
| https://dx.doi.org/10.1093/tropej/fmm039 |
| https://dx.doi.org/10.1093/inthealth/ihz035 |
| http://dx.doi.org/10.1056/NEJMc0706263 |
| http://dx.doi.org/10.3109/09286586.2010.545500 |
| https://dx.doi.org/10.1371/journal.pntd.0006080 |
| http://dx.doi.org/10.1186/s40249-017-0358-3 |
| http://dx.doi.org/10.1371/journal.pone.0065918 |
| https://dx.doi.org/10.1007/s11427-016-5060-3 |
| https://dx.doi.org/10.1167/iovs.11-7372 |
| http://dx.doi.org/10.1371/journal.pntd.0002415 |
| http://dx.doi.org/10.1371/journal.pntd.0004352 |
| http://dx.doi.org/10.1371/journal.pone.0178595 |
| http://dx.doi.org/10.1001/jamaophthalmol.2017.3062 |
| http://dx.doi.org/10.1038/s41598-018-21127-0 |
| http://dx.doi.org/10.1371/journal.pntd.0007834 |
| http://dx.doi.org/10.1038/s41598-020-71833-x |
| http://dx.doi.org/10.1080/09286586.2017.1293693 |
| https://dx.doi.org/10.1186/s12879-019-4495-0 |
| 2685 |
| https://dx.doi.org/10.1136/bjo.2006.099150 |
| https://dx.doi.org/10.1167/iovs.08-2247 |
| http://dx.doi.org/10.1007/s13149-015-0446-1 |
| http://dx.doi.org/10.1016/j.trstmh.2009.02.007 |

| |
|---|
| https://dx.doi.org/10.1001/jamaophthalmol.2013.2356 |
| https://dx.doi.org/10.1371/journal.pntd.0005003 |
