## Supplementary material for "Comparison of methods for assessing *Chlamydia trachomatis* transmission intensity: a systematic review": S7 - Analysis by region

Studies by region

| Region (World Health Organization) | Number of studies included in review | Percent of total number of studies* |
|---|---|---|
| African Region | 177 | 75% |
| Region of the Americas | 14 | 6% |
| Eastern Mediterranean Region | 14 | 6% |
| South-East Asia Region | 7 | 3% |
| Western Pacific Region | 19 | 8% |
| Country not specified | 2 | 1% |

* Total number of studies is 235. Studies may have included data from one or more regions.
