## Supplementary material for "Comparison of methods for assessing *Chlamydia trachomatis* transmission intensity: a systematic review": S8 - Bland Altman plot

Bland-Altman plot: Field grade vs. photo grade, follicular trachoma*

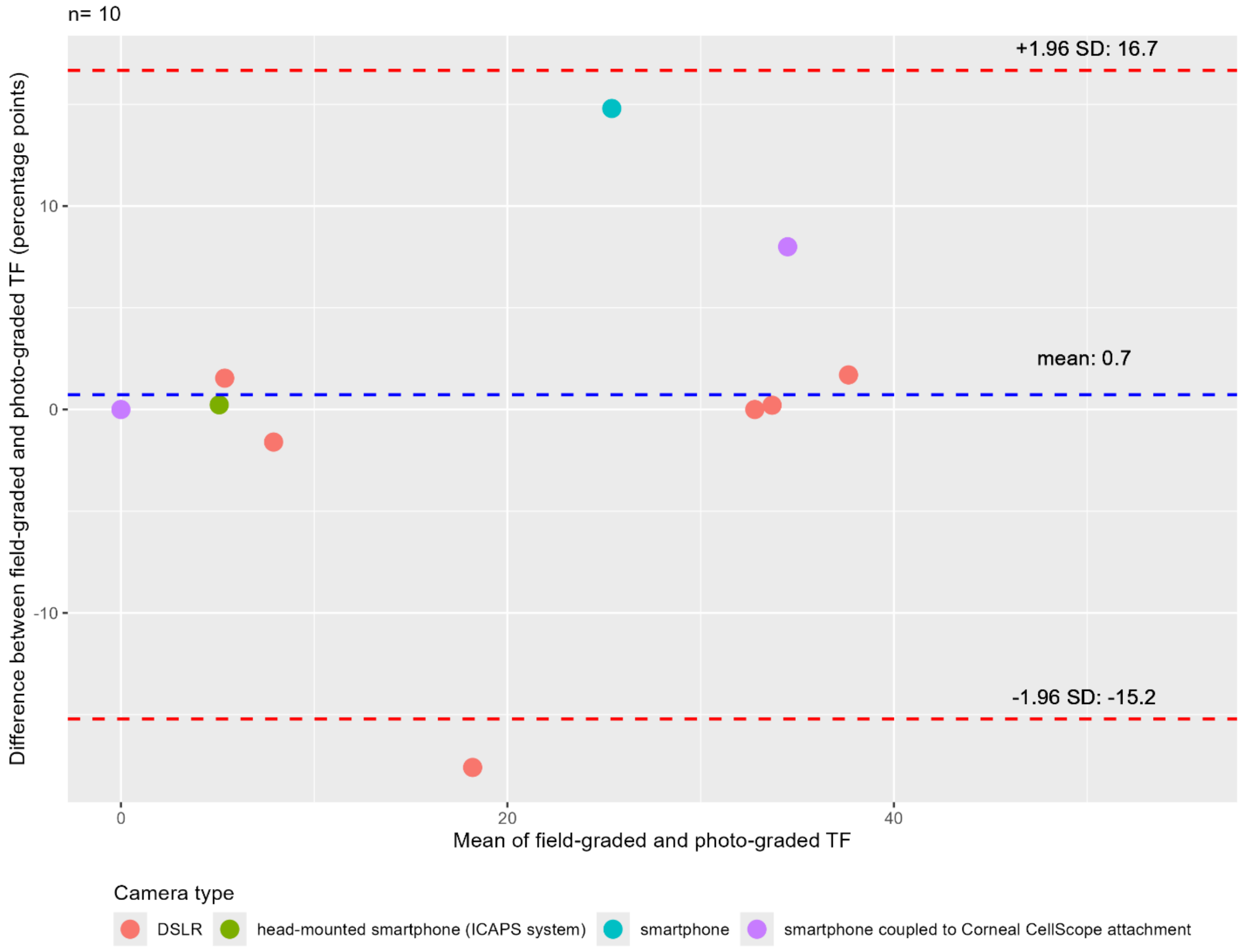


*measured as trachomatous inflammation—follicular in the simplified World Health Organization system or F2/F3 in the Follicles, Papillae, Cicatrices system

TF = follicular trachoma, SD = standard deviation, DSLR = digital single-lens reflex
