## Supplementary figures and images for "Comparison of methods for assessing *Chlamydia trachomatis* transmission intensity: a systematic review"

### S9 - TI vs. infection plot

Supplemental File S9

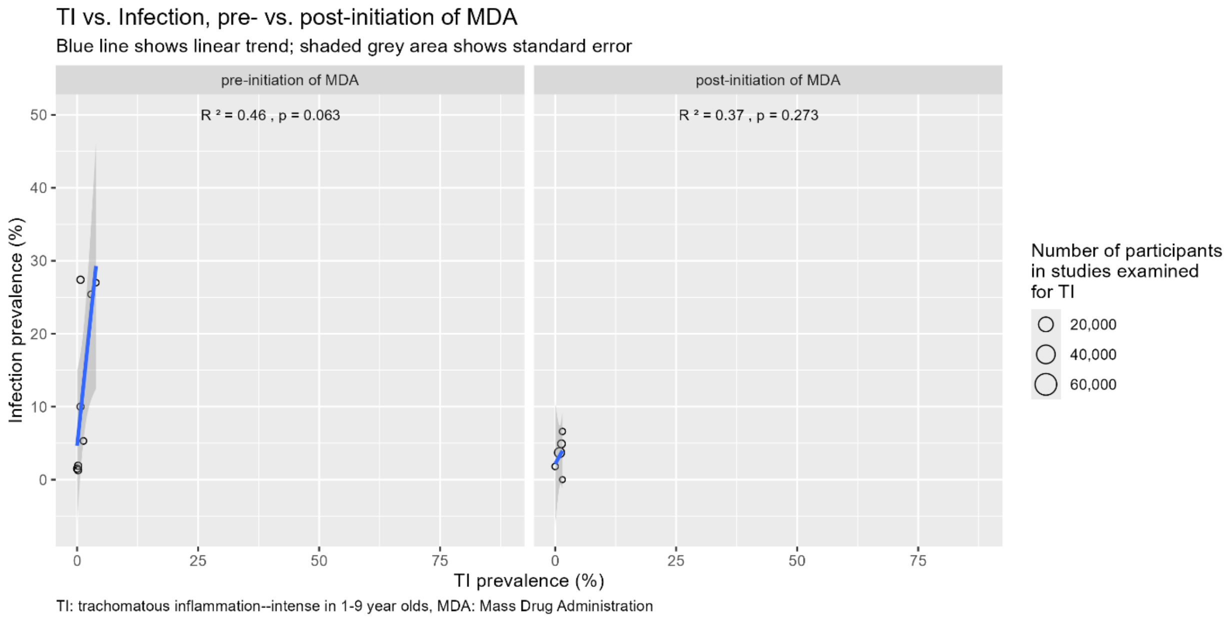
