## Supplementary material for "Comparison of methods for assessing *Chlamydia trachomatis* transmission intensity: a systematic review": S11 - TF vs. infection, TF vs. SCR

TF vs. Infection, TF vs. SCR in subset of populations with all three indicators

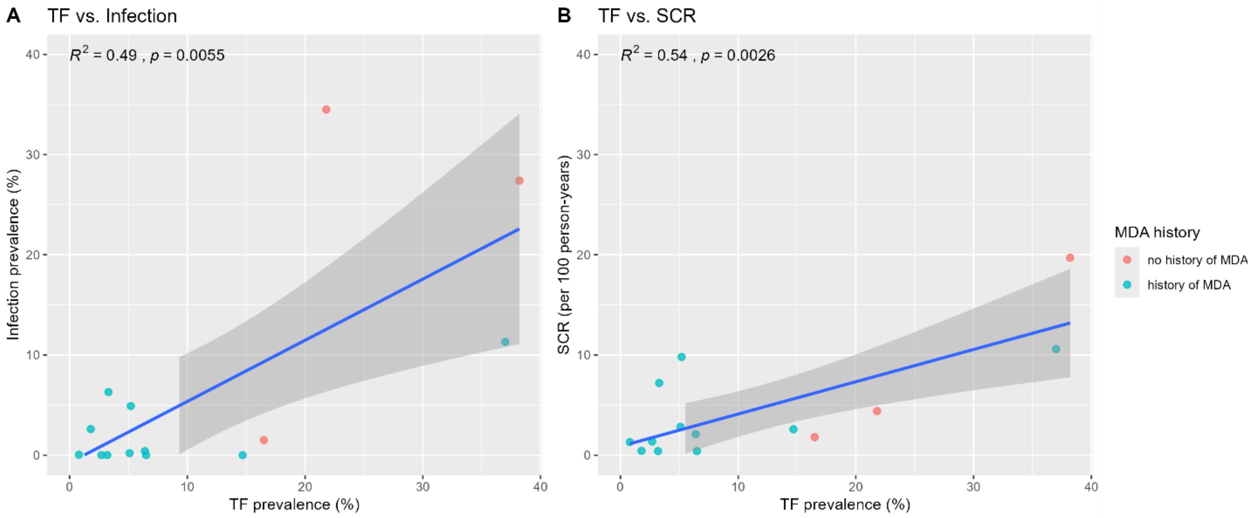


TF: trachomatous inflammation—follicular in children 1–9, SCR: seroconversion rate, MDA: mass drug administration
